# Genetic insights into immunothrombosis: from shared loci to repurposed drugs for autoimmune and thrombotic diseases

**DOI:** 10.64898/2026.02.28.26346627

**Authors:** Yichen Long, Yang Ou, Guangxiang Huang, Xin Tan, Shengyan Zhao, Lang Min, Chunyan Sun, Zeyu Luo, Hui Pan

**Author notes:** These authors contributed equally (Yichen Long, Yang Ou, Guangxiang Huang).

## Abstract

**Objective:** Autoimmune diseases (ADs) markedly elevate venous thromboembolism (VTE) risk, yet the shared genetic architecture and tissue-specific regulatory mechanisms of this “Autoimmune-Thrombotic Axis” remain poorly defined. We aimed to characterize the genomic landscape of immunothrombosis to identify causal links and therapeutic targets.

**Approach and Results:** We integrated large-scale GWAS data for VTE and 16 ADs using a multi-omics framework, including pleiotropy scanning, local genetic correlation, and summary-based Mendelian randomization (SMR). We identified 21 Immunothrombotic Shared Loci (ISLs) and 274 pleiotropic genes enriched in complement and coagulation cascades. Mendelian randomization (MR) analysis revealed a robust causal effect of genetically predicted systemic lupus erythematosus (SLE) on VTE risk (OR = 1.018, 95% CI: 1.008-1.029, P = 0.0003). Mechanistically, IL6R and PLCL1 emerged as central mediators with distinct tissue-specific regulatory partitioning. Colocalization confirmed that shared genetic susceptibility is primarily driven by expression in arterial tissues (aorta and coronary) rather than exclusively in immune cells. Furthermore, the lead SNP rs4129267 was identified as a potential predictor for VTE in rheumatoid arthritis patients, and drug prioritization nominated TNF inhibitors as promising candidates for mitigating thrombotic burden.

**Conclusion:** This study establishes the first genomic atlas of the autoimmune-thrombotic axis, demonstrating that vasculature-specific gene regulation drives immunothrombosis. These findings provide a biological basis for VTE risk stratification and suggest that genotype-guided therapy may optimize vascular outcomes in AD patients.

## INTRODUCTION

Venous thromboembolism (VTE), comprising deep vein thrombosis and pulmonary embolism, is a major vascular disorder and a leading cause of global morbidity and mortality(1, 2). While classical risk factors—such as surgery, malignancy, and immobility—are well established, growing evidence highlights the importance of systemic inflammation and immune dysregulation in thrombogenesis(3). Autoimmune diseases (ADs), defined by chronic immune activation and loss of immunological tolerance, are increasingly recognized as significant contributors to VTE risk(4, 5).

Epidemiological studies consistently show that patients with autoimmune disorders—including systemic lupus erythematosus (SLE), rheumatoid arthritis (RA), inflammatory bowel disease (IBD), and multiple sclerosis (MS)—have a markedly higher risk of VTE than the general population(5–7). A large UK record-linkage study showed that hospitalization for immune-mediated diseases was associated with a significant increase in VTE incidence(5). Moreover, systematic reviews suggest that certain ADs not only elevate first-event VTE risk but may also increase recurrence risk, although estimates vary across studies(8). Proposed biological mechanisms include immune-mediated endothelial injury, platelet activation, and dysregulation of coagulation and fibrinolytic pathways(3, 4).

Clinically, managing thrombosis in patients with ADs is challenging. Early manifestations of VTE may be subtle or nonspecific, resulting in missed diagnoses in routine practice(9). Decisions about long-term anticoagulation are further complicated by heterogeneous patient profiles: while some individuals benefit from extended therapy, others may be exposed to unnecessary bleeding risks. Data from the RIETE registry show that, in AD patients, the risk of major bleeding during anticoagulation is similar to the risk of VTE recurrence after therapy withdrawal(10). Importantly, VTE events in ADs often result from acute “second-hit” triggers—such as surgery, trauma, infection, myocardial infarction, or disease flares—underscoring the multifactorial nature of thrombosis in these populations(11).

Emerging genomic evidence indicates that ADs and VTE may share common genetic underpinnings. Multiple genome-wide association studies (GWAS) have identified loci implicated in both immune regulation and thrombotic susceptibility. For example, *SH2B3*, a key regulator of hematopoietic and immune signaling, has been associated with several ADs and VTE(12, 13). Variants in *IL6R*, *STAT4*, and *TNFAIP3*, which are central to cytokine signaling and inflammatory control, further suggest biological links between immune dysfunction and vascular activation(14, 15). A recent Mendelian randomization (MR) study also reported a potential causal effect of genetically predicted ulcerative colitis on VTE(15), reinforcing the hypothesis of shared mechanisms.

However, the extent and structure of the shared genetic architecture between ADs and VTE remain incompletely defined. Most previous work has focused on single diseases or isolated loci, limiting the ability to uncover genome-wide pleiotropy, tissue-specific effects, and convergent biological pathways. A systematic cross-trait approach is needed to characterize genetic correlation, identify pleiotropic loci, and dissect regulatory mechanisms that link immune dysregulation with thrombosis.

Using large-scale GWAS datasets, this study aims to comprehensively map the shared genetic architecture and causal relationships between VTE and multiple ADs. Through an integrated framework—including genetic correlation analysis, pleiotropy scanning, functional annotation, colocalization, MR, and drug-target prioritization—we seek to elucidate the mechanisms underlying immunothrombosis and identify therapeutic opportunities for drug repurposing.

## METHODS

### Data sources

GWAS summary statistics for VTE(16) and sixteen ADs—including Crohn’s disease (CD)(17), dermatomyositis/polymyositis (DMPM)(18), Guillain-Barré syndrome (GBS)(18), Graves’ disease (GD)(19), Hashimoto’s thyroiditis (HT)(19), myasthenia gravis (MG)(18), MS(20), neuromyelitis optica (NMO)(21), psoriasis(19), RA(22), SLE(23), systemic sclerosis (SSC)(18), Sjögren’s syndrome (SS)(18), type 1 diabetes (T1D)(24), UC(17), and ankylosing spondylitis (AS)(18)—were obtained from large-scale biobanks and publicly available resources (FinnGen, BioBank Japan, deCODE Genetics, GWAS Catalog, and published meta-analyses; Table S1).

These datasets comprised predominantly European (Finland, Iceland, U.S.), with a smaller subset of traits derived from studies including both European and East Asian ancestry samples (Japan, Republic of Korea, China) and typically included hundreds of thousands of participants per trait (NMO being an exception with ∼1,500 cases). For traits available in multiple ancestral groups, we preferentially used European-ancestry GWAS summary statistics when available. For traits with robust evidence across both ancestries, we used cross-ancestry meta-analyzed summary statistics, restricted to loci that were replicated or reached genome-wide significance in both European and East Asian populations.

All summary statistics were harmonized using standard allele-alignment and strand-flip corrections.

### Genetic correlation

We estimated genome-wide genetic correlations between VTE and each AD using linkage disequilibrium score regression (LDSC)(25). Because the majority of GWAS summary statistics were derived from European-ancestry populations, precomputed LD scores derived from European-ancestry samples in the 1000 Genomes Project (1KG) were used. For traits analyzed using cross-ancestry meta-analyzed summary statistics, European LD scores were also applied, which is a commonly adopted and validated approach when the meta-analysis is dominated by European samples and focuses on loci showing consistent effects across ancestries. The extended human leukocyte antigen (HLA) region excluded due to complex linkage disequilibrium (LD)(26).

As a complementary approach, we applied high-definition likelihood (HDL)(27), which yields more efficient, lower-variance correlation estimates. Only trait pairs showing consistent direction and magnitude of genetic correlation across LDSC and HDL were prioritized, reducing potential bias arising from population heterogeneity. Trait pairs with concordant signals across methods were prioritized for subsequent analysis of shared immunothrombotic pathways.

### Cross-trait pleiotropy analysis at the SNP-level

We identified shared pleiotropic single-nucleotide polymorphisms (SNPs) using two complementary approaches: Pleiotropic Analysis under Composite Null Hypothesis (PLACO)(28) and Cross-Phenotype Association (CPASSOC)(29). PLACO is optimized to detect true cross-trait pleiotropy, whereas CPASSOC accommodates heterogeneity of effects; SNPs reaching genome-wide significance (P < 5×10⁻⁸) in both tests were considered high-confidence pleiotropic variants.

### Functional annotation

Significant pleiotropic SNPs were annotated using Functional Mapping and Annotation of Genetic Associations (FUMA, v1.5.0) (30), integrating positional mapping, eQTL mapping, and chromatin interaction data. SNPs were annotated to gene-level with Multi-marker Analysis of GenoMic Annotation (MAGMA v1.10)(31), which aggregates SNP-level statistics to gene-level Z-scores while accounting for LD(32).

Pleiotropic genes identified by MAGMA were conceptualized as candidate Immunothrombosis Gene Modules underwent pathway enrichment against curated gene sets from the Molecular Signatures Database (MSigDB)(33), including canonical pathways (C2) and Gene Ontology (GO) terms (C5). Enrichment P-values were adjusted by the Benjamini-Hochberg false discovery rate (FDR).

### Local genetic correlations

To detect locus-specific sharing, we used Local Analysis of [co]Variant Association (LAVA) (34). LAVA estimates local genetic correlations across predefined genomic segments, enabling detection of regions showing concordant or opposing effects between traits.

Genomic regions annotated as risk loci by FUMA and passing LAVA significance criteria (FDR < 0.05) were defined as immunothrombotic shared loci (ISLs). Analyses used LD matrices derived from European samples in the 1KG reference panel.

### Summary-based Mendelian randomization (SMR)

To prioritize genes whose expression may mediate trait associations, we applied the SMR framework(35), which integrates GWAS with eQTL data and uses the Heterogeneity in Dependent Instruments (HEIDI) test to distinguish pleiotropy from linkage. The cis-expression quantitative trait loci (cis-eQTL) summary data was retrieved from the eQTLGen consortium (whole blood)(36) and the Genotype-Tissue Expression project (GTEx) v8 (multiple tissues including whole blood, EBV-transformed lymphocytes, cultured fibroblasts, skeletal muscle, thyroid, spleen, arterial tissues, and skins)(37).

Significance criteria: SMR associations with Benjamini-Hochberg FDR < 0.05 and HEIDI P > 0.01 with at least three SNPs were retained as candidate mediators.

### Bayesian colocalization analysis

We performed Bayesian colocalization using the coloc R package (v5.2.1). Posterior probabilities were computed for five hypotheses describing shared versus distinct causal variants. Regions with posterior probability for a shared causal variant (PP.H4) > 0.90 were considered colocalized. For trait-trait colocalization we examined SNPs within LAVA-defined ISLs; for trait-eQTL colocalization we used SNPs within ±500 kb of the gene.

### Causal inference analysis

To evaluate possible causal effects of 5 ADs on VTE risk, we performed a one-directional two-sample Mendelian randomization (MR) analysis. Genotype data from 1000 Genomes Project Phase 3 panels(26) of European descent and PLINK 1.9 software was used to undertake LD clumping, where *r*^2^ was set to 0.001 and window size was set to a physical distance of 10,000 KB(26). The number of independent genetic instruments selected for each autoimmune disease is reported in Table 2 (No. of SNPs). This allowed us to identify independent significance SNPs (*P* < 5 × 10^-8^) for MR analyses, which was necessary to ensure MR estimates were not biased by using correlated instruments. To reduce bias from horizontal pleiotropy, SNPs located in loci with known shared effects on autoimmune disease and VTE (e.g., IL6, IL6R) were carefully evaluated, and sensitivity analyses using pleiotropy-robust MR methods were performed. Next, the exposure and outcome data were harmonized to ensure the concordance of effect alleles between the datasets, and SNPs with non-concordant alleles as well as palindromic SNPs were excluded from the instrumental variables. MR results were further corrected for multiple testing across autoimmune diseases using FDR adjustment. Given the extensive genetic pleiotropy observed between autoimmune diseases and VTE, MR analyses were considered exploratory and intended to assess consistency of evidence rather than to establish definitive causal relationships.

For univariable MR, the inverse variance weighted (IVW)(38) method was first employed as the initial analysis. The weighted median(39) and MR-Egger methods(40) were further applied to enhance the robustness and replicability of the findings. Sensitivity was assessed using both the IVW and MR-Egger approaches. All genetic instruments for the exposures were harmonized with the outcome data using the “TwoSampleMR” R package(41). In addition, the MR-PRESSO test was implemented to detect horizontal pleiotropic outliers among the instrumental variables, and effect estimates were recalculated after outlier removal using the “MR-PRESSO” R package(42).

### Drug prioritization analysis

We mapped pleiotropic genes from MAGMA to drug-gene interactions using DrugCentral(43), DGIdb(44), and PharmGKB(45). Enrichment and prioritization analyses were performed with the clusterProfiler R package, ranking candidate drugs by target significance and cross-disease relevance to define a therapeutic repositioning landscape.

## RESULTS

### Shared genetic architecture between ADs and VTE

The LDSC and HDL methods yielded largely consistent genetic correlation profiles (Table 1). Specifically, LDSC analysis identified eight ADs with significant positive genetic correlations with VTE: HT (rg = 0.32, P = 1.91×10⁻⁹), psoriasis (rg = 0.23, P = 1.30×10⁻⁵), RA (rg = 0.11, P = 4.00×10⁻⁴), SLE (rg = 0.16, P = 8.46×10⁻⁵), SS (rg = 0.14, P = 3.03×10⁻²), T1D (rg = 0.08, P = 8.20×10⁻³), DMPM (rg = 0.17, P = 6.78×10⁻²), and GBS (rg = 0.09, P = 3.63×10⁻¹). In contrast, MS showed a significant negative genetic correlation with VTE (rg = −0.14, P = 1.13×10⁻²). Using the HDL approach, significant positive genetic correlations were confirmed between VTE and five ADs: HT (rg = 0.63, P = 7.50×10⁻³), psoriasis (rg = 0.29, P = 2.35×10⁻³), RA (rg = 0.17, P = 4.74×10⁻⁴), SLE (rg = 0.16, P = 2.98×10⁻³), and SS (rg = 0.22, P = 2.76×10⁻²). Based on the concordance between methods, these five trait pairs—showing significant, stable, and positive genetic correlations—were selected for further analysis of the Autoimmune-Thrombotic axis.

**Table 1.** Genetic correlation between ADs and VTE.

| Trait pairs | LDSC |  | HDL |  |
| --- | --- | --- | --- | --- |
| | $r_g(S \times 10)$ | P | $r_g(S \times 10)$ | P |
| VTE & CD | -0.10 (0.17) | $5.79 \times 10^{-01}$ | -0.004 (0.01) | $6.06 \times 10^{-01}$ |
| VTE & DMPM | 0.17 (0.09) | $6.78 \times 10^{-02}$ | NA | NA |
| VTE & GBS | 0.09 (0.10) | $3.63 \times 10^{-01}$ | NA | NA |
| VTE & GD | 0.02 (0.35) | $7.24 \times 10^{-01}$ | NA | NA |
| VTE & HT | 0.32 (0.05) | $1.91 \times 10^{-09}$ | 0.63 (0.24) | $7.50 \times 10^{-03}$ |
| VTE & MG | 0.15 (0.06) | $1.71 \times 10^{-02}$ | NA | NA |
| VTE & MS | -0.14 (0.05) | $1.13 \times 10^{-02}$ | -0.10 (0.04) | $1.89 \times 10^{-02}$ |
| VTE & NMO | 0.13 (0.13) | $3.46 \times 10^{-01}$ | NA | NA |
| VTE & psoriasis | 0.23 (0.05) | $1.30 \times 10^{-05}$ | 0.29 (0.10) | $2.35 \times 10^{-03}$ |
| VTE & RA | 0.11 (0.03) | $4.00 \times 10^{-04}$ | 0.17 (0.05) | $4.74 \times 10^{-04}$ |
| VTE & SLE | 0.16 (0.04) | $8.46 \times 10^{-05}$ | 0.16 (0.05) | $2.98 \times 10^{-03}$ |
| VTE & SSC | 0.23 (0.27) | $3.84 \times 10^{-01}$ | NA | NA |
| VTE & SS | 0.14 (0.06) | $3.03 \times 10^{-02}$ | 0.22 (0.10) | $2.76 \times 10^{-02}$ |
| VTE & T1D | 0.08 (0.03) | $8.20 \times 10^{-03}$ | NA | NA |
| VTE & UC | 0.13 (0.15) | $3.76 \times 10^{-01}$ | -0.01 (0.001) | $1.15 \times 10^{-25}$ |
| VTE & AS | 0.05 (0.06) | $3.65 \times 10^{-01}$ | 0.10 (0.08) | $1.80 \times 10^{-01}$ |

Abbreviations: LDSC, linkage disequilibrium score regression; HDL, high-definition likelihood; $r_g$ , genetic correlation; SE, standard error; AD, autoimmune disease; VTE, venous thromboembolism; CD, Crohn's disease; DMPM, Dermatomyositis and polymyositis; GBS, Guillain-Barré syndrome; GD, Graves' disease; HT, Hashimoto's thyroiditis; MG, myasthenia gravis; MS, multiple sclerosis; NMO, neuromyelitis optica; RA, rheumatoid arthritis; SLE, systemic lupus erythematosus; SSC, systemic sclerosis; SS, Sjögren's syndrome; T1D, type 1 diabetes; UC, ulcerative colitis; AS, ankylosing spondylitis.

### Pleiotropic SNPs, loci and genes for ADs and VTE

Based on the significant genetic correlations observed between VTE and multiple ADs—including RA, SLE, SS, psoriasis, and HT—we further performed pleiotropy analysis to identify shared pleiotropic SNPs. We applied two complementary statistical approaches, PLACO and CPASSOC, and considered the overlapping SNPs identified by both methods as robust pleiotropic SNPs (Supplementary Figure 1 and 2).

A total of 21 ISLs. were defined as genomic regions that were functionally annotated as risk loci by FUMA and exhibited significant local genetic correlation with VTE in LAVA analysis (Figure 1). Regional Manhattan plots highlighted six colocalized loci (PP.H4 > 0.9) associated with RA, which displayed the largest number of shared loci with VTE. These regions are located at cytobands 7p21.2, 7p15.1, 4p14, 2q11.2, and 1q21.3 (Supplementary Figure 3A-E). HT also showed a prominent shared genomic locus at chr12q24.12-q24.13 (Supplementary Figure 3F). More importantly, rs4129267 emerged as a common lead SNP within the 1q21.3 ISL, showing a shared genetic association with VTE and RA and suggesting a potential contribution to VTE susceptibility in RA patients. MAGMA identified 274 shared pleiotropic genes between VTE and ADs (Table S2), which collectively represent core Immunothrombosis Gene Modules significantly enriched in multiple immune- and coagulation-related pathways (Figure 2). Key enriched pathways included “Formation of Fibrin Clot (Clotting Cascade)” from Reactome (FDR = 3.34×10⁻⁷, 9 genes), “Complement and coagulation cascades” from Kyoto Encyclopedia of Genes and Genomes (KEGG) (FDR = 4.48×10⁻⁹, 12 genes), and “Antigen processing and presentation” from both KEGG (FDR = 1.20×10⁻⁶, 9 genes) and GO Process (FDR = 6.23×10⁻⁵, 10 genes). Additional significantly enriched pathways were “Immune system process” from GO Process (FDR = 6.23×10⁻⁵, 44 genes) and “MHC protein complex” from GO Component (FDR = 0.0066, 5 genes).

**Figure 1.**
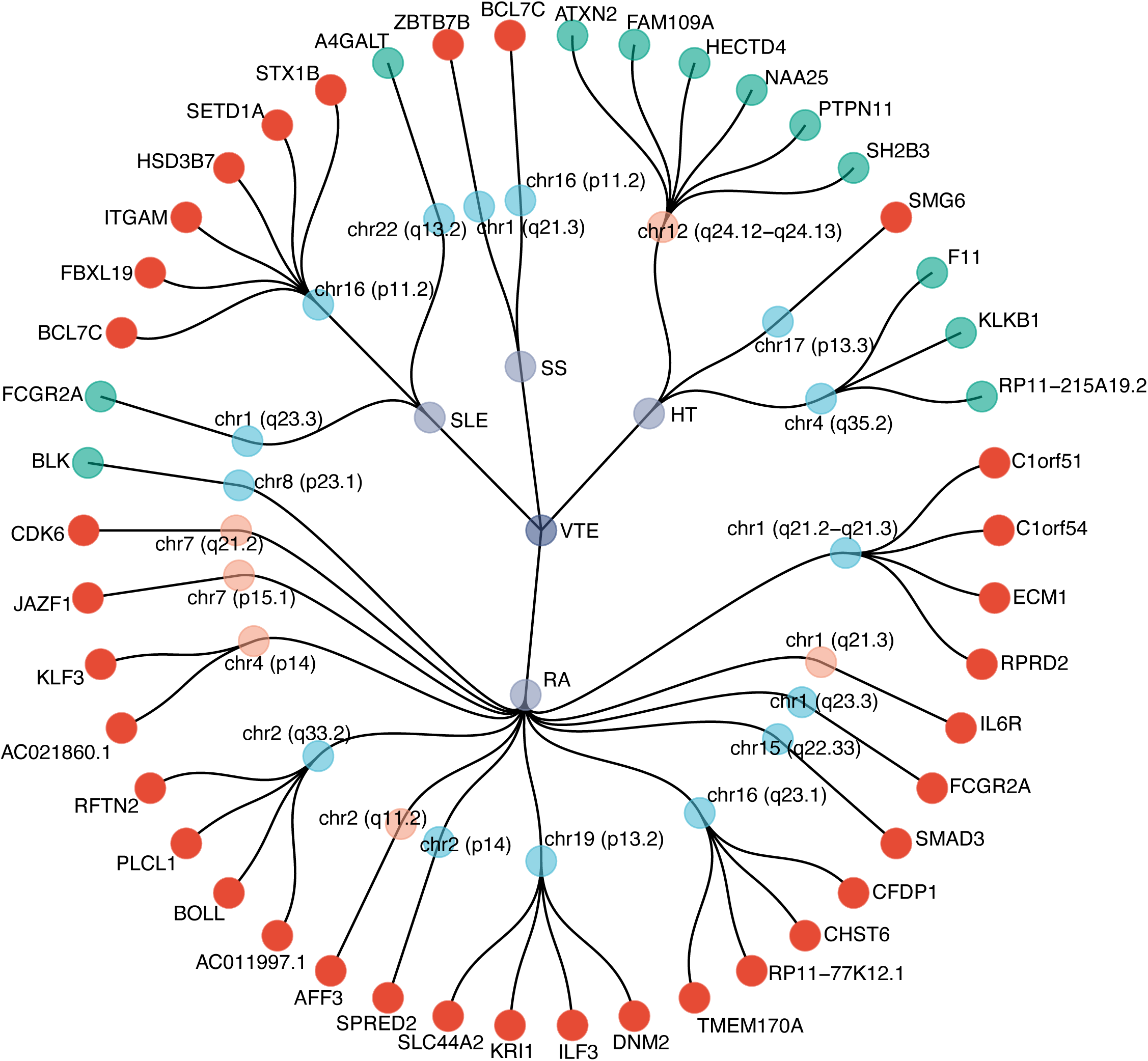
Dendrogram of ISLs and pleiotropic genes across disease pairs. ISLs with evidence from two-trait colocalization are highlighted in orange; shared pleiotropic genes supported by eQTL mapping are highlighted in red. Abbreviations: ISLs, Immunothrombotic Shared Loci; RA, rheumatoid arthritis; SLE, systemic lupus erythematosus; SS, Sjögren’s syndrome; VTE, venous thromboembolism; eQTL, expression quantitative trait locus; HT, Hashimoto’s thyroiditis.

**Figure 2.**
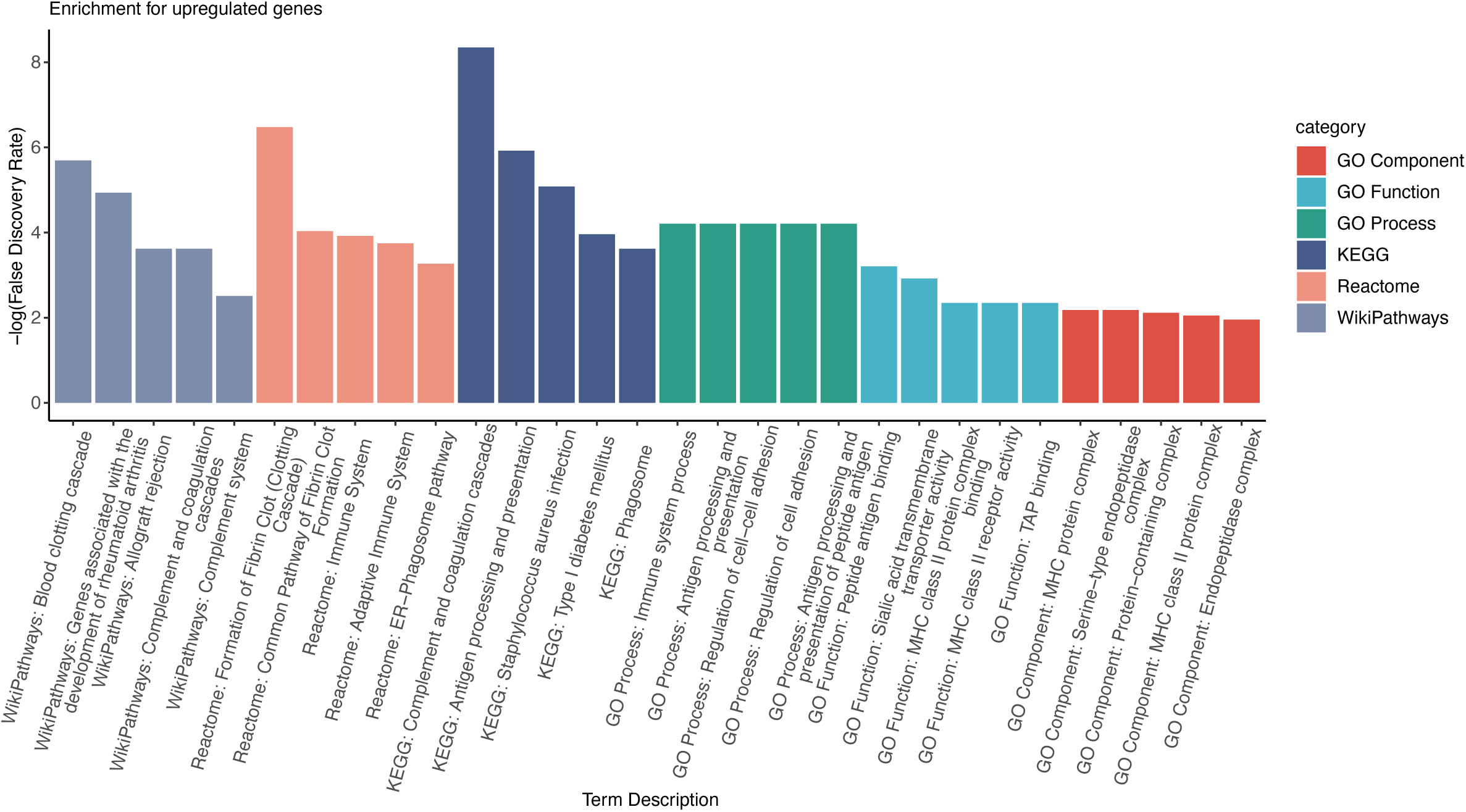
Pathway Enrichment Analysis of Immunothrombosis Gene Modules between VTE and ADs. Bar chart displaying the significantly enriched biological pathways from the analysis of 274 pleiotropic genes constituting core Immunothrombosis Gene Modules. Abbreviations: AD, autoimmune disease; VTE, venous thromboembolism.

### The landscape of pleiotropic genes within pleiotropic loci

To further investigate the functional relevance of the identified pleiotropic genes, we performed comprehensive colocalization and SMR analyses to elucidate potential gene-level associations and tissue-specific regulatory mechanisms across the Autoimmune-Thrombotic Axis (RA, SLE, HT, psoriasis, and SS). Both analyses were conducted under stringent criteria: SMR associations were considered significant at FDR < 0.05 (Benjamini-Hochberg corrected) with HEIDI p-value > 0.01, while colocalization required a posterior probability of PP.H4 > 0.9.

Colocalization analysis revealed strong evidence across multiple gene-trait-tissue combinations (Supplementary Figures 4-15), with SMR further identifying numerous significant gene-tissue-disease associations (Supplementary Figures 19-24). Notably, two genes—*PLCL1* and *IL6R*—were consistently supported by both SMR and colocalization across multiple tissues in VTE and ADs, emerging as key nodes within the Immunothrombosis Gene Modules(Figure 3).

**Figure 3.**
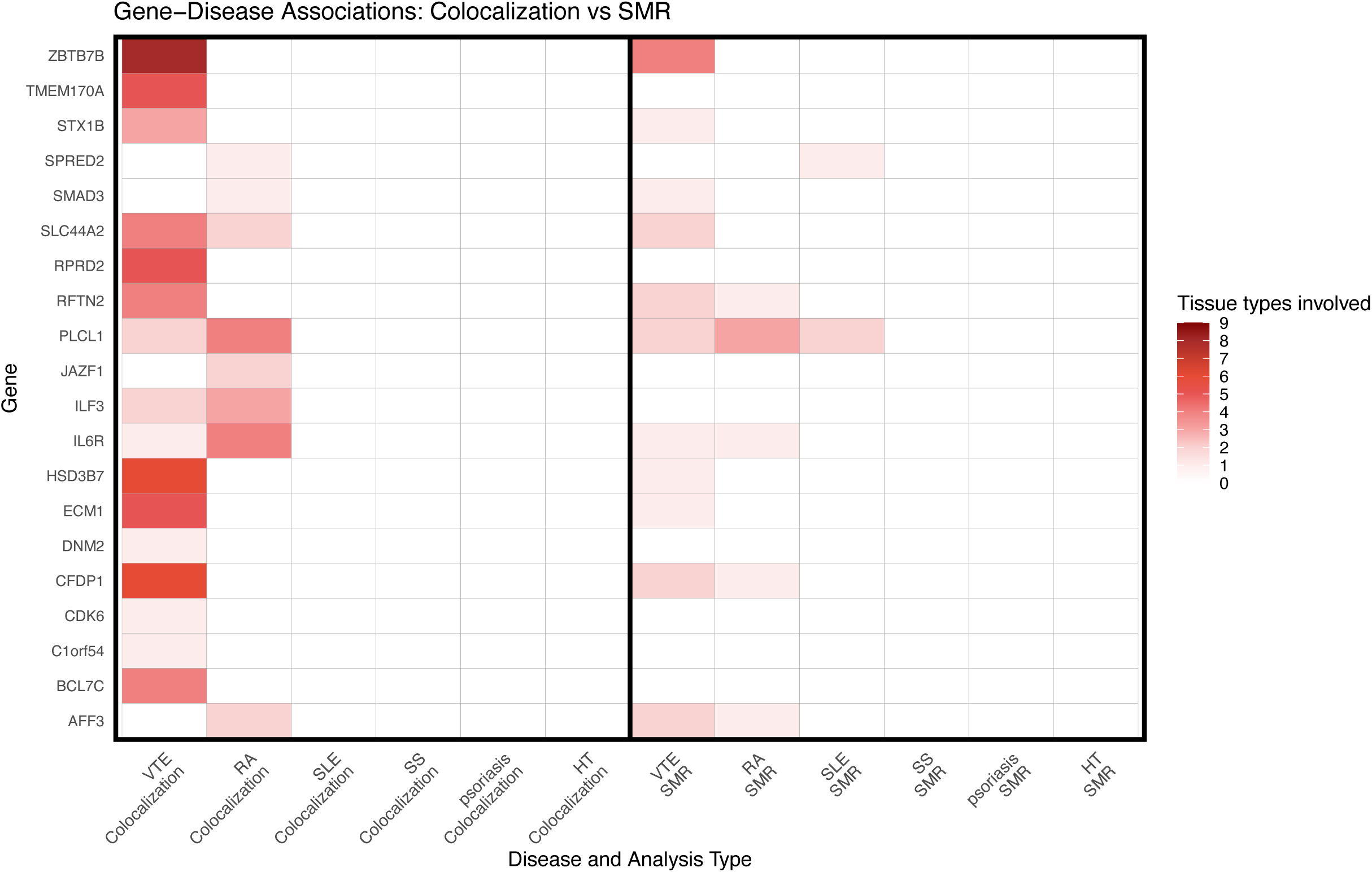
Tissue involvement in colocalization and SMR analyses across the Autoimmune-Thrombotic Axis. Colocalization analysis assesses whether gene expression and disease share a common causal variant in a specific genomic region; SMR evaluates the potential causal influence of gene expression on disease risk. Color intensity indicates the frequency of tissue involvement. Abbreviations: RA, rheumatoid arthritis; SLE, systemic lupus erythematosus; SMR, summary-based Mendelian randomization; SS, Sjögren’s syndrome; VTE, venous thromboembolism; HT, Hashimoto’s thyroiditis.

Regional colocalization plots for *IL6R* and *PLCL1* (Supplementary Figures 15 and 16) revealed distinct patterns of trait-tissue-gene regulatory architecture. For *IL6R*, colocalization with RA in EBV-transformed lymphocytes was driven by a single lead SNP, rs11265608 (Supplementary Figure 15A), which did not serve as the lead variant in any other trait-tissue combination. In contrast, RA- or VTE-*IL6R* colocalizations in arterial tissues (coronary, aorta, tibial) were driven by a different set of lead SNPs—rs7518199, rs6664201, rs7529229, and rs4129267—all of which reside in strong LD with each other (Supplementary Figure 16B-E, Supplementary Figure 18A). Thus, *IL6R* colocalizations are underpinned by two distinct LD-defined SNP clusters: one specific to RA in lymphocytes, and another shared across RA/VTE in arterial tissues (Supplementary Figure 18A), supporting a shared genetic mechanism rather than individual-level risk prediction.

For *PLCL1*, a similar partitioning of trait-tissue lead SNPs was observed. Colocalizations of RA with *PLCL1* in spleen or whole blood were driven by rs1579695, rs10497813, rs700649, and rs700679—a tightly linked set of SNPs indicative of an immune-related regulatory module (Supplementary Figure 17A-C). In contrast, RA- or VTE-*PLCL1* colocalizations in arterial tissues and thyroid were associated with a separate group of lead SNPs, including rs2293255 and rs16826873, which formed a distinct high-LD cluster (Supplementary Figure 17D). These results indicate that *PLCL1* exhibits locus-specific regulatory partitioning within the ISLs, with one LD block associated with immune tissues and another with arterial contexts (Supplementary Figure 18B).

### The causal relationship between ADs and VTE estimated by MR

Based on the established genetic correlations between ADs and VTE, we performed MR analysis to assess potential causal relationships (Table 2). Using the IVW method, a statistically significant association between genetically predicted SLE and VTE risk was observed (OR = 1.018, 95% CI: 1.008-1.029, P = 0.0003). Although RA showed nominal significance in the IVW analysis (OR = 1.016, 95% CI: 1.001-1.032, P = 0.042), this association did not remain significant after correction for multiple testing and was not consistently supported by complementary MR methods.

**Table 2.** MR Analysis of the Causal Effects of ADs on VTE Risk.

| Exposure | Outcome (No.of<br>SNPs) | IVW |  | MR-Egger |  | Weighted median |  |
| --- | --- | --- | --- | --- | --- | --- | --- |
|  |  | OR (95% CI) | P-value | OR (95% CI) | P-value | OR (95% CI) | P-value |
| SLE | VTE (107) | 1.02 (1.01-1.03) | 0 | 1.03 (1.01-1.05) | 0.02 | 1.01 (1.01-1.03) | 0.01 |
| RA | VTE (202) | 1.02 (1.00-1.03) | 0.04 | 0.97 (0.95-1.00) | 0.03 | 1.00 (0.99-1.02) | 0.80 |
| HT | VTE (70) | 1.01 (1.00-1.03) | 0.12 | 1.01 (0.99-1.04) | 0.35 | 1.01 (0.99-1.03) | 0.59 |
| SS | VTE (23) | 1.01 (0.99-1.02) | 0.34 | 0.99 (0.97-1.01) | 0.33 | 1.01 (0.99-1.03) | 0.23 |
| psoriasis | VTE (57) | 1.00 (0.98-1.02) | 0.86 | 0.97 (0.93-1.01) | 0.12 | 0.99 (0.97-1.01) | 0.30 |

Abbreviations: IVW, inverse-variance weighted; MR, Mendelian randomization; VTE, venous thromboembolism; SLE, systemic lupus erythematosus; RA, rheumatoid arthritis; HT, Hashimoto's thyroiditis; SS, Sjögren's syndrome; OR, odds ratio; CI, confidence interval.

For SLE, the direction of effect was broadly consistent across complementary MR methods, with both MR-Egger (OR = 1.028, 95% CI: 1.005-1.052, P = 0.017) and weighted median (OR = 1.018, 95% CI: 1.005-1.031, P = 0.007) analyses yielding concordant results (Figure 4B). The scatter plot revealed a consistent positive slope across all MR methods, visually confirming the significant causal association between SLE and VTE (Figure 4C). Sensitivity analyses further reinforced the robustness of the SLE-VTE association. While Cochran’s Q test indicated moderate heterogeneity (IVW Q = 195, I² = 45.5%, P = 3.43×10⁻⁷), the MR-Egger intercept test showed no evidence of horizontal pleiotropy (intercept = −0.0018, P = 0.346) (Table S3). MR-PRESSO analysis confirmed the stability of the causal estimate, with the outlier-corrected result (beta = 0.0196, SD = 0.0044, P = 2.16×10⁻⁵) remaining significant and directionally consistent with the primary IVW estimate. The symmetrical funnel plot provided additional evidence against directional pleiotropy (Figure 4D).

**Figure 4.**
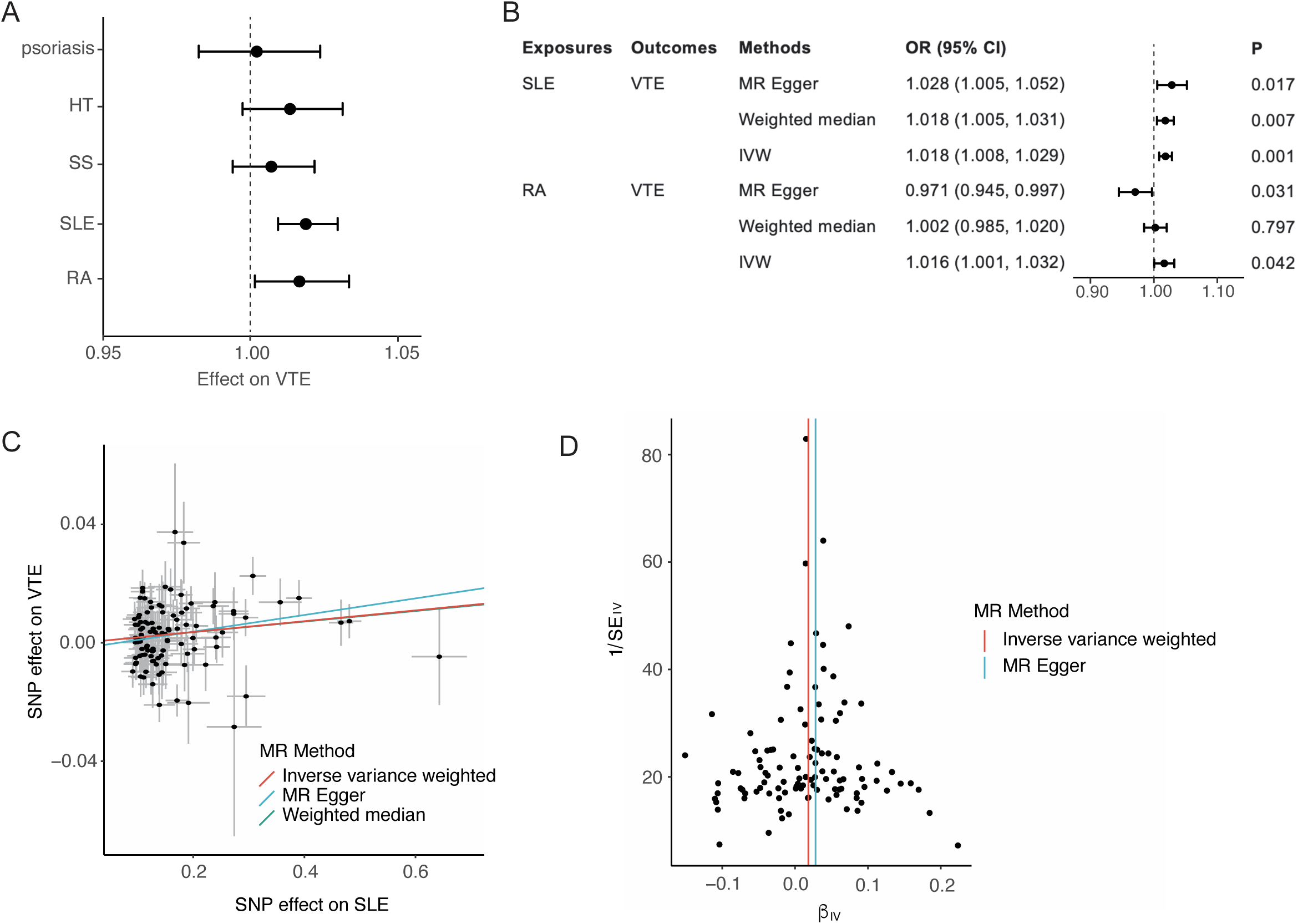
Causal inference across the Autoimmune-Thrombotic Axis. (A) Forest plot of causal association estimates for selected ADs on VTE using the inverse variance weighted (IVW) method in MR analysis. (B) Causal effect estimates of SLE and RA on VTE assessed by multiple MR methods. (C) Scatter plot demonstrating the significant causal association between SLE and VTE. (D) Funnel plot showing symmetry of the causal estimates, indicating no directional pleiotropy for the SLE-VTE association. Abbreviations: AD, autoimmune disease; IVW, inverse variance weighted; MR, Mendelian randomization; RA, rheumatoid arthritis; SLE, systemic lupus erythematosus; VTE, venous thromboembolism.

Collectively, these results support a putative causal effect of SLE on VTE risk, with consistent findings across multiple MR methods and no significant pleiotropic bias. However, the magnitude of the observed effect was modest (OR ≈ 1.02), indicating that SLE explains only a small proportion of VTE risk and does not imply strong individual-level risk prediction.

### Potential drugs identification for AD and VTE

To identify potential therapeutic agents targeting the shared pathophysiology between ADs and VTE, we performed a drug enrichment analysis based on pleiotropic genes identified by MAGMA, using drug-gene interaction data from DrugCentral, DGIdb, and PharmGKB.

An UpSet plot displays the overlap of candidate drugs across different AD-VTE pairs (Figure 5A). The top 10 most significant drug candidates for each pair—VTE with SS, SLE, RA, psoriasis, or HT—are shown in Figure 5B-F. Among these, drugs labeled in red represent those shared across systemic ADs (SS, SLE, and RA), highlighting consistent repurposing opportunities for immunothrombotic disorders. Although few in number, these shared agents—including ALLOPURINOL, ADALIMUMAB, INFLIXIMAB, ETANERCEPT, and other TNF-ALPHA INHIBITORS—are likely to target core inflammatory and thrombotic pathways common to systemic autoimmunity and VTE. Notably, WARFARIN and ANTITHROMBIN ALFA were recurrently identified across all five AD-VTE contexts, underscoring their broad antithrombotic and immunomodulatory potential.

**Figure 5.**
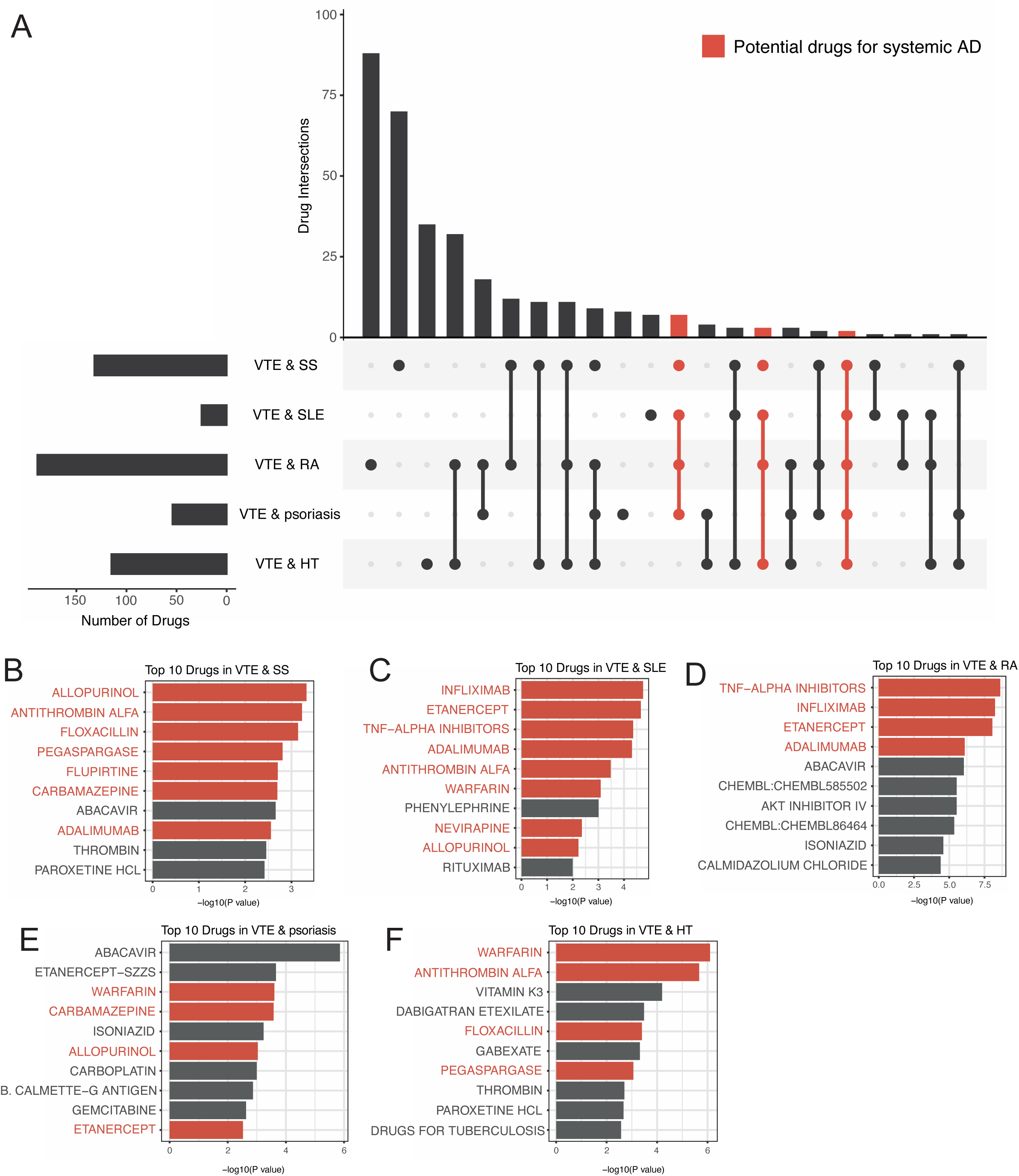
Identification of potential drugs targeting the Autoimmune-Thrombotic Axis. (A) Upset plot displaying the intersections of candidate drugs identified across different disease pairs. (B-F) The top 10 most significant drug candidates for VTE & SS, VTE & SLE, VTE & RA, VTE & psoriasis, and VTE & HT, respectively. Drugs highlighted in red represent those identified across all systemic AD (SS, SLE, and RA) & VTE pairs. These findings define a therapeutic repositioning landscape for AD-VTE comorbidity. Abbreviations: AD, autoimmune disease; RA, rheumatoid arthritis; SLE, systemic lupus erythematosus; SS, Sjögren’s syndrome; VTE, venous thromboembolism; HT, Hashimoto’s thyroiditis.

Together, these results define a therapeutic repositioning landscape for co-occurring autoimmune and thrombotic conditions.

## DISCUSSION

Our study elucidates the shared genetic architecture and causal relationships between ADs and VTE through an integrative multi-omics framework. We first established significant positive genetic correlations between VTE and five ADs (RA, SLE, SS, psoriasis, and HT), collectively defining a broad Autoimmune-Thrombotic Axis. MR analysis further delineated a statistically supported but modest causal effect of genetically predicted SLE on VTE risk, contrasting with less consistent evidence for RA. Mechanistically, we identified 21 ISLs and 274 pleiotropic genes constituting core Immunothrombosis Gene Modules, which were significantly enriched in key immune and coagulation pathways, directly linking autoimmune dysregulation to thrombotic mechanisms. Colocalization and SMR analyses pinpointed *IL6R* and *PLCL1* as key mediators underpinning this link, revealing their distinct, tissue-specific regulatory architectures. More importantly, rs4129267 within the 1q21.3 ISL is a potential predictor for VTE in RA patients. Finally, drug enrichment analysis nominated promising repurposable candidates, including warfarin and TNF inhibitors, which emerged as recurrent hits across multiple AD-VTE pairs. These findings collectively define a therapeutic repositioning landscape with tangible translational potential.

The causal association between genetically predicted SLE and VTE observed in our Mendelian randomization analysis aligns with well-established clinical observations that SLE patients are prone to thrombotic events(46–48). Importantly, the small effect size observed in MR reflects an average population-level genetic contribution rather than a clinically deterministic risk factor, and therefore should not be interpreted as explaining the full thrombotic burden observed in SLE patients. The pathophysiology of SLE predisposes patients to VTE through an imbalance between coagulation and fibrinolysis, with coagulation abnormalities encompassing hypercoagulability, endothelial injury, and platelet activation—the classical Virchow triad. First, chronic immune dysregulation induces a hypercoagulable state: impaired control of the tissue factor pathway, reduced TFPI activity(49), and the presence of antiphospholipid antibodies (aPL), often in patients with overlapping APS, form β2-glycoprotein I-dependent immune complexes that activate coagulation, while neutrophil extracellular traps (NETs) provide a scaffold for thrombin generation(50, 51). In addition, alterations in anticoagulant pathways have been reported in subsets of SLE patients, although evidence for consistent deficiencies in natural anticoagulants such as protein C, protein S, or antithrombin remains limited and heterogeneous across studies(52). Second, endothelial injury amplifies thrombotic risk: SLE patients exhibit increased circulating apoptotic endothelial cells, impaired microvascular function, and immune complex-mediated vascular inflammation, reflecting both immune-mediated endothelial apoptosis and SLE-associated vasculitis(53–55). This endothelial dysfunction exposes procoagulant surfaces and facilitates platelet adhesion, linking immune activation and vascular inflammation to thrombosis. Third, platelet activation and hemodynamic factors further contribute: aPL directly activate platelets(56), while neutrophil extracellular traps (NETs) and vascular abnormalities promote platelet aggregation and adhesion(49). Elevated fibrinogen and dysregulated tissue factor pathway signaling sustain a prothrombotic environment independent of immune cell activation(57). Moreover, fibrinolytic impairment in SLE, including elevated fibrinogen and reduced plasmin-mediated clot breakdown, further amplifies thrombus formation(52). Collectively, these mechanisms create a self-perpetuating immunothrombotic milieu in SLE, providing a mechanistic explanation for the causal association between genetically predicted SLE and VTE observed in our MR analysis.

*PLCL1* encodes a catalytically inactive phospholipase C-like protein that nonetheless participates in intracellular signalling(58), and regulates receptor turnover and integrin activity(59), in the control of immune-cell function. The *PLCL1* locus harbors genetic variants associated with susceptibility to multiple immune-mediated diseases, such as CD(60), SLE(61), and allergic disorders(62). Integrative genomics work has demonstrated colocalization between a PU.1 transcription factor QTL (tfQTL) at this locus and CD GWAS; this tfQTL physically contacts the *PLCL1* promoter in neutrophils and monocytes, and shows allelic imbalance and strong eQTL effects on *PLCL1* expression in both cell types(63). Moreover, in RA, *PLCL1* promotes inflammation in fibroblast-like synoviocytes (FLS) by activating the *NLRP3* inflammasome, which drives production of pro-inflammatory cytokines (IL-6, IL-1β, CXCL8) and synovial cell activation, linking *PLCL1* to autoimmunity and immune cell trafficking(64). Although there is no direct evidence tying *PLCL1* to coagulation or thrombosis, its role in intracellular signaling, integrin regulation in myeloid cells(65), and amplification of systemic inflammation (e.g., via IL-6)(66) provide a biologically plausible bridge to pro-thrombotic risk within the Immunothrombosis Gene Modules.

In RA, elevated IL-6 is a major driver of systemic inflammation, contributing not only to articular damage but also to a distinctly pro-thrombotic milieu(67). IL-6 promotes hypercoagulability by inducing key procoagulant mediators such as tissue factor (TF), factor VIII (FVIII), and von Willebrand factor (VWF)(68), while simultaneously impairing endogenous anticoagulant pathways through downregulation of protein S(68). In parallel, IL-6 enhances platelet biogenesis, increasing circulating platelet mass, and further augments platelet thrombogenicity by activating platelet 12-lipoxygenase, which exposes pro-coagulant phospholipids and facilitates thrombin generation(69). Collectively, these IL-6-driven alterations disrupt vascular homeostasis and substantially heighten thrombotic risk in RA.

Rs11265608, the lead SNP identified in *IL6R* eQTL colocalization with RA in EBV-transformed lymphocytes, is located within an exon of *IL6R-AS*1 and may regulate the expression of associated genes(70). Meanwhile, rs4129267, included in the cluster of lead SNPs for RA-VTE-*IL6R* colocalizations across arterial tissues (coronary, aorta, tibial), has been reported to disrupt interactions between myocardial infarction- and coronary artery disease-associated open chromatin regions in cardiac fibroblasts(71). While IL6R represents a potential therapeutic target, it is important to note that modulating IL6R alone may not fully address the underlying source of inflammation, unless specific polymorphisms heighten IL6R sensitivity. More importantly, rs4129267 is a common lead SNP within the 1q21.3 ISL, potentially predicting VTE in RA patients. However, the clinical utility of rs4129267 requires validation using individual-level data and independent cohorts.

Although IL-6 clearly contributes to the pro-thrombotic milieu in RA, the impact of therapeutically targeting the IL-6 pathway on thrombotic risk remains controversial. Clinical studies of IL-6 receptor blockade have shown improvements in coagulation biomarkers such as D-dimer, and fibrinogen generation, suggesting a potential reduction in inflammation-driven hypercoagulability(72). Mechanistically, IL-6 signals through the JAK-STAT pathway, meaning that JAK inhibitors can indirectly modulate IL-6-mediated effects. Unlike IL-6 receptor blockade, which has shown beneficial effects on coagulation biomarkers, JAK inhibitors have raised safety concerns. In the ORAL Surveillance trial, tofacitinib was associated with higher rates of major adverse cardiovascular events compared with TNF inhibitors in older RA patients or those with cardiovascular risk factors(73). While meta-analyses of RCTs suggest that, in the short term, JAK inhibitors do not significantly increase the risk of cardiovascular events, MACE, or VTE, postmarketing and long-term data indicate that higher doses of tofacitinib and baricitinib may increase thromboembolic risk(74). Thus, despite well-defined IL-6-mediated mechanisms of thrombosis, the net thrombotic effect of IL-6-pathway-directed therapies—especially JAK inhibitors, which also modulate IL-6 signaling via JAK-STAT-remains unsettled and warrants further long-term, real-world evaluation.

While our study provides comprehensive insights into the shared genetic architecture of ADs and VTE, several limitations should be acknowledged. First, although most GWAS summary statistics used were derived from European-ancestry populations, a subset of traits relied on cross-ancestry meta-analyzed data. For these traits, we focused on loci with evidence of replication or genome-wide significance in both European and East Asian populations; nevertheless, the use of European LD reference panels may not fully capture ancestry-specific LD patterns. Second, despite employing multiple MR methods and sensitivity analyses to mitigate pleiotropy, residual horizontal pleiotropy could still potentially bias the causal estimates. Third, the sample size for some ADs (e.g., NMO) was relatively small, which might have reduced the power to detect shared genetic signals for these less common conditions. Finally, while we identified promising drug repurposing candidates, these are computational predictions primarily reflecting genetic susceptibility to VTE and do not distinguish between prophylactic and therapeutic use. Their translational relevance requires validation in experimental and clinical studies, considering potential bleeding risks.

## CONCLUSIONS

In conclusion, our study provides a robust genetic foundation for the clinical co-occurrence of ADs and VTE, establishing the Autoimmune-Thrombotic Axis as a central framework. We establish a specific causal role for SLE in driving VTE risk and reveal a broader shared genetic architecture across multiple ADs characterized by ISLs and Immunothrombosis Gene Modules bridging immune and coagulation systems. More importantly, rs4129267 within the 1q21.3 ISL is a potential predictor for VTE in RA patients. These findings advocate for integrated clinical management and open a therapeutic repositioning landscape for drugs that simultaneously target autoimmune and thrombotic processes.

## Data Availability

This study is a secondary analysis of publicly available data. All genome-wide association study (GWAS) summary statistics used in this work were obtained from public databases and consortia (e.g., FinnGen, GWAS Catalog, eQTLGen, GTEx). The specific sources are detailed in the Methods section and Table S1. No new primary data were generated.

## SUMMARY OF ABBREVIATIONS

AD: Autoimmune Disease
APS: Antiphospholipid Syndrome
AS: Ankylosing Spondylitis
CD: Crohn’s Disease
CPASSOC: Cross-Phenotype Association
DMPM: Dermatomyositis/Polymyositis
eQTL: Expression Quantitative Trait Loci
FDR: False Discovery Rate
FLS: Fibroblast-like Synoviocytes
FUMA: Functional Mapping and Annotation of Genetic Associations
GBS: Guillain-Barré Syndrome
GD: Graves’ Disease
GO: Gene Ontology
GTEx: Genotype-Tissue Expression project
GWAS: Genome-Wide Association Study
HDL: High-Definition Likelihood
HEIDI: Heterogeneity in Dependent Instruments
HT: Hashimoto’s Thyroiditis
IBD: Inflammatory Bowel Disease
IVW: Inverse Variance Weighted
KEGG: Kyoto Encyclopedia of Genes and Genomes
LAVA: Local Analysis of [co]Variant Association
LD: Linkage Disequilibrium
LDSC: Linkage Disequilibrium Score Regression
MAGMA: Multi-marker Analysis of GenoMic Annotation
MACE: Major Adverse Cardiovascular Events
MG: Myasthenia Gravis
MR: Mendelian Randomization
MS: Multiple Sclerosis
MSigDB: Molecular Signatures Database
NETs: Neutrophil Extracellular Traps
NMO: Neuromyelitis Optica
PLACO: Pleiotropic Analysis under Composite Null Hypothesis
PP.H4: Posterior Probability for Hypothesis 4
RA: Rheumatoid Arthritis
SMR: Summary-based Mendelian Randomization
SLE: Systemic Lupus Erythematosus
SNP: Single-Nucleotide Polymorphism
SS: Sjögren’s Syndrome
SSC: Systemic Sclerosis
T1D: Type 1 Diabetes
tfQTL: Transcription Factor Quantitative Trait Locus
UC: Ulcerative Colitis
VTE: Venous Thromboembolism
1KG: 1000 Genomes Project

## Ethics approval and consent to participate

Not applicable.

## Consent for publication

Not applicable.

## Availability of data and materials

The datasets used and analysed during the current study are all sourced from public databases.

## Statement

During the preparation of this work the authors used ChatGPT 4.5 in order to refine the English language. After using this tool, the authors reviewed and edited the content as needed and take full responsibility for the content of the published article.

## Authors’ contributions

Y.L. and Y.O. contributed equally as co-first authors. Y.O. conceptualized the study and designed the methodology. Y.L. conducted the formal analysis, performed the genetic analyses, and led the data curation process. G.H. also contributed equally as a co-first author and was responsible for validation, data interpretation, and visualization.

X.T. and S.Z. assisted with data collection, investigation, and provided critical technical support for the analyses. L.M. and C.S. contributed to resource provisioning, project administration, and preliminary data processing. Z.L. and H.P. served as co-corresponding authors, jointly supervising the research project. They acquired funding, provided overall supervision, and were responsible for manuscript review, editing, and final approval. All authors reviewed the manuscript drafts and approved the final version for submission.

## Acknowledgements

This project was supported by the National Natural Science Foundation of China (No. 82504843 and 82570245). We would like to express our gratitude to the researchers and consortia that made their GWAS summary statistics publicly available, enabling this work.

## Supplementary Figure Legends

Supplementary Figure 1. Manhattan plots from the PLACO analysis across the Autoimmune-Thrombotic Axis. The blue line indicates the suggestive significance threshold (1 × 10⁻⁵), and the red line indicates the genome-wide significance threshold (5 × 10⁻⁸). (A) VTE & RA; (B) VTE & SLE; (C) VTE & SS; (D) VTE & psoriasis; (E) VTE & HT. Abbreviations: HT, Hashimoto’s thyroiditis; PLACO, pleiotropy analysis and cross-phenotype analysis; RA, rheumatoid arthritis; SLE, systemic lupus erythematosus; SS, Sjögren’s syndrome; VTE, venous thromboembolism.

Supplementary Figure 2. Manhattan plots from the CPASSOC analysis across the Autoimmune-Thrombotic Axis. The blue line indicates the suggestive significance threshold (1 × 10⁻⁵), and the red line indicates the genome-wide significance threshold (5 × 10⁻⁸). (A) VTE & RA; (B) VTE & SLE; (C) VTE & SS; (D) VTE & psoriasis; (E) VTE & HT. Abbreviations: CPASSOC, Cross-Phenotype Association Analysis; HT, Hashimoto’s thyroiditis; RA, rheumatoid arthritis; SLE, systemic lupus erythematosus; SS, Sjögren’s syndrome; VTE, venous thromboembolism.

Supplementary Figure 3. Regional association plots of ISLs between AD and VTE. (A-E) ISLs for RA and VTE located at 1q21.3, 2q11.2, 4p14, 7p15.1, and 7p21.2, respectively. (F) An ISL for HT and VTE at chr12q24.12-q24.13. All loci shown satisfy PP.H4 > 0.9, indicating strong evidence for a shared causal variant. Abbreviations: HT, Hashimoto’s thyroiditis; ISLs, Immunothrombotic Shared Loci; PP.H4, posterior probability for hypothesis 4 (colocalization); RA, rheumatoid arthritis; VTE, venous thromboembolism.

Supplementary Figure 4. Colocalization analysis across the Autoimmune-Thrombotic Axis: AD or VTE and eQTL of pleiotropic genes in whole blood tissue (GTEx). Abbreviations: AD, autoimmune disease; eQTL, expression quantitative trait locus; VTE, venous thromboembolism.

Supplementary Figure 5. Colocalization analysis across the Autoimmune-Thrombotic Axis: AD or VTE and eQTL of pleiotropic genes in whole blood tissue (eQTLgen). Abbreviations: AD, autoimmune disease; eQTL, expression quantitative trait locus; VTE, venous thromboembolism.

Supplementary Figure 6. Colocalization analysis across the Autoimmune-Thrombotic Axis: AD or VTE and eQTL of pleiotropic genes in thyroid tissue. Abbreviations: AD, autoimmune disease; eQTL, expression quantitative trait locus; VTE, venous thromboembolism.

Supplementary Figure 7. Colocalization analysis across the Autoimmune-Thrombotic Axis: AD or VTE and eQTL of pleiotropic genes in spleen tissue. Abbreviations: AD, autoimmune disease; eQTL, expression quantitative trait locus; VTE, venous thromboembolism.

Supplementary Figure 8. Colocalization analysis across the Autoimmune-Thrombotic Axis: AD or VTE and eQTL of pleiotropic genes in sun exposed (lower leg) skin. Abbreviations: AD, autoimmune disease; eQTL, expression quantitative trait locus; VTE, venous thromboembolism.

Supplementary Figure 9. Colocalization analysis across the Autoimmune-Thrombotic Axis: AD or VTE and eQTL of pleiotropic genes in sun unexposed (suprapubic) skin. Abbreviations: AD, autoimmune disease; eQTL, expression quantitative trait locus; VTE, venous thromboembolism.

Supplementary Figure 10. Colocalization analysis across the Autoimmune-Thrombotic Axis: AD or VTE and eQTL of pleiotropic genes in skeletal muscle. Abbreviations: AD, autoimmune disease; eQTL, expression quantitative trait locus; VTE, venous thromboembolism.

Supplementary Figure 11. Colocalization analysis across the Autoimmune-Thrombotic Axis: AD or VTE and eQTL of pleiotropic genes in EBV-transformed lymphocytes. Abbreviations: AD, autoimmune disease; eQTL, expression quantitative trait locus; VTE, venous thromboembolism.

Supplementary Figure 12. Colocalization analysis across the Autoimmune-Thrombotic Axis: AD or VTE and eQTL of pleiotropic genes in cultured fibroblasts. Abbreviations: AD, autoimmune disease; eQTL, expression quantitative trait locus; VTE, venous thromboembolism.

Supplementary Figure 13. Colocalization analysis across the Autoimmune-Thrombotic Axis: AD or VTE and eQTL of pleiotropic genes in artery (aorta). Abbreviations: AD, autoimmune disease; eQTL, expression quantitative trait locus; VTE, venous thromboembolism.

Supplementary Figure 14. Colocalization analysis across the Autoimmune-Thrombotic Axis: AD or VTE and eQTL of pleiotropic genes in artery (coronary). Abbreviations: AD, autoimmune disease; eQTL, expression quantitative trait locus; VTE, venous thromboembolism.

Supplementary Figure 15. Colocalization analysis across the Autoimmune-Thrombotic Axis: AD or VTE and eQTL of pleiotropic genes in artery (tibial). Abbreviations: AD, autoimmune disease; eQTL, expression quantitative trait locus; VTE, venous thromboembolism.

Supplementary Figure 16. Colocalization analysis of *IL6R* multi-tissue eQTL with traits across the Autoimmune-Thrombotic Axis. (A) Cells (EBV-transformed lymphocytes) eQTL with RA; (B) Artery-Aorta eQTL with RA; (C) Artery-Tibial eQTL with RA; (D) Artery-Coronary eQTL with RA; (E) Artery-Aorta eQTL with VTE. Abbreviations: eQTL, expression quantitative trait locus; RA, rheumatoid arthritis; VTE, venous thromboembolism.

Supplementary Figure 17. Colocalization analysis of *PLCL1* multi-tissue eQTL with traits across the Autoimmune-Thrombotic Axis. (A) Spleen eQTL with RA; (B) Artery-Coronary eQTL with RA; (C) Whole Blood (eQTLGen) eQTL with RA; (D) Artery-Tibial eQTL with VTE; (E) Thyroid eQTL with VTE. Abbreviations: eQTL, expression quantitative trait locus; RA, rheumatoid arthritis; VTE, venous thromboembolism.

Supplementary Figure 18. Linkage Disequilibrium Heatmaps of the *IL6R* (A) and *PLCL1* (B) Loci from Multi-Tissue eQTL Colocalization Analysis. The color scale represents the degree of linkage disequilibrium (r²) between SNPs, with red indicating high r² (strong LD) and blue indicating low r² (weak LD). Abbreviations: LD, linkage disequilibrium.

Supplementary Figure 19. SMR analysis across the Autoimmune-Thrombotic Axis: VTE and eQTL from multiple tissues. (A) Aorta Artery eQTL from GTEx and VTE; (B) Coronary Artery eQTL from GTEx and VTE; (C) Tibial Artery eQTL from GTEx and VTE; (D) Skeletal Muscle eQTL from GTEx and VTE; (E) EBV-transformed lymphocytes eQTL from GTEx and VTE; (F) Spleen eQTL from GTEx and VTE; (G) Whole Blood eQTL from GTEx and VTE; (H) Whole Blood eQTL from eQTLGen and VTE. Abbreviations: eQTL, expression quantitative trait locus; GTEx, Genotype-Tissue Expression project; SMR, summary-based Mendelian randomization; VTE, venous thromboembolism.

Supplementary Figure 20. SMR analysis across the Autoimmune-Thrombotic Axis: RA and eQTL from multiple tissues. (A) Aorta Artery eQTL from GTEx and RA; (B) Coronary Artery eQTL from GTEx and RA; (C) Tibial Artery eQTL from GTEx and RA; (D) Skeletal Muscle eQTL from GTEx and RA; (E) EBV-transformed lymphocytes eQTL from GTEx and RA; (F) Spleen eQTL from GTEx and RA; (G) Whole Blood eQTL from GTEx and RA; (H) Whole Blood eQTL from eQTLGen and RA. Abbreviations: eQTL, expression quantitative trait locus; GTEx, Genotype-Tissue Expression project; RA, rheumatoid arthritis; SMR, summary-based Mendelian randomization.

Supplementary Figure 21. SMR analysis across the Autoimmune-Thrombotic Axis: SLE and eQTL from multiple tissues. (A) Aorta Artery eQTL from GTEx and SLE; (B) Coronary Artery eQTL from GTEx and SLE; (C) Tibial Artery eQTL from GTEx and SLE; (D) Skeletal Muscle eQTL from GTEx and SLE; (E) EBV-transformed lymphocytes eQTL from GTEx and SLE; (F) Spleen eQTL from GTEx and SLE; (G) Whole Blood eQTL from GTEx and SLE; (H) Whole Blood eQTL from eQTLGen and SLE. Abbreviations: eQTL, expression quantitative trait locus; GTEx, Genotype-Tissue Expression project; SLE, systemic lupus erythematosus; SMR, summary-based Mendelian randomization.

Supplementary Figure 22. SMR analysis across the Autoimmune-Thrombotic Axis: SS and eQTL from multiple tissues. (A) Aorta Artery eQTL from GTEx and SS; (B) Coronary Artery eQTL from GTEx and SS; (C) Tibial Artery eQTL from GTEx and SS; (D) Skeletal Muscle eQTL from GTEx and SS; (E) EBV-transformed lymphocytes eQTL from GTEx and SS; (F) Spleen eQTL from GTEx and SS; (G) Whole Blood eQTL from GTEx and SS; (H) Whole Blood eQTL from eQTLGen and SS. Abbreviations: eQTL, expression quantitative trait locus; GTEx, Genotype-Tissue Expression project; SMR, summary-based Mendelian randomization; SS, Sjögren’s syndrome.

Supplementary Figure 23. SMR analysis across the Autoimmune-Thrombotic Axis: psoriasis and eQTL from multiple tissues. (A) Aorta Artery eQTL from GTEx and psoriasis; (B) Coronary Artery eQTL from GTEx and psoriasis; (C) Tibial Artery eQTL from GTEx and psoriasis; (D) Skeletal Muscle eQTL from GTEx and psoriasis; (E) EBV-transformed lymphocytes eQTL from GTEx and psoriasis; (F) Spleen eQTL from GTEx and psoriasis; (G) Whole Blood eQTL from GTEx and psoriasis; (H) Whole Blood eQTL from eQTLGen and psoriasis. Abbreviations: eQTL, expression quantitative trait locus; GTEx, Genotype-Tissue Expression project; SMR, summary-based Mendelian randomization.

Supplementary Figure 24. SMR analysis across the Autoimmune-Thrombotic Axis: HT and eQTL from multiple tissues. (A) Aorta Artery eQTL from GTEx and HT; (B) Coronary Artery eQTL from GTEx and HT; (C) Tibial Artery eQTL from GTEx and HT; (D) Skeletal Muscle eQTL from GTEx and HT; (E) EBV-transformed lymphocytes eQTL from GTEx and HT; (F) Spleen eQTL from GTEx and HT; (G) Whole Blood eQTL from GTEx and HT; (H) Whole Blood eQTL from eQTLGen and HT. Abbreviations: eQTL, expression quantitative trait locus; GTEx, Genotype-Tissue Expression project; HT, Hashimoto’s thyroiditis; SMR, summary-based Mendelian randomization.

