## supplementary files for "Genetic insights into immunothrombosis: from shared loci to repurposed drugs for autoimmune and thrombotic diseases"

Table S1 Sources and description of summary-level data

| Trait | abbrev | case | ctl | all | Discovery ancestry |
|---|---|---|---|---|---|
| Crohn’s disease | CD | 20,873 | 346,719 | 367,592 | EUR, EAS |
| Dermatopolymyositis | DMPM | 500 | 374,317 | 37,817 | EUR |
| Guillain-Barre syndrome | GBS | 551 | 491,583 | 492,134 | EUR |
| Graves' disease | GD | 4,487 | 629,598 | 634,085 | EUR, EAS |
| Hashimoto’s thyroiditis | HT | 16,191 | 552,642 | 568,833 | EUR, EAS |
| Myasthenia gravis | MG | 560 | 495,667 | 495,667 | EUR |
| Multiple sclerosis | MS | 4,888 | 10,395 | 15,283 | EUR |
| Neuromyelitis optica | NMO | 215 | 1,244 | 1,459 | EUR |
| Psoriasis | - | 5,278 | 650,391 | 655,669 | EUR, EAS |
| Rheumatoid arthritis | RA | 31,313 | 995,377 | 1,026,690 | EUR |
| Systemic lupus erythematosus | SLE | 8,798 | 16,470 | 25,268 | EUR, EAS |
| Systemic sclerosis | SSC | 313 | 499,508 | 499,821 | EUR |
| Sjögren syndrome | SS | 3,309 | 484,260 | 487,569 | EUR |

| | | | | | |
|---|---|---|---|---|---|
| Type 1 diabetes | T1D | 18,942 | 501,638 | 520,580 | EUR |
| Ulcerative colitis | UC | 22,318 | 353,190 | 375,508 | EUR, EAS |
| Ankylosing spondylitis | AS | 1,705 | 496,755 | 498,460 | EUR |
| Venous thromboembolism | VTE | 81,190 | 1,419,671 | 1,500,861 | EUR |

Table S2 Shared pleiotropic genes identified by MAGMA between VTE and ADs

| GENE | CHR | START | STOP | NSNPS | N | ZSTAT | P | SYMBOL | Disease Pair |
|---|---|---|---|---|---|---|---|---|---|
| ENSG00000081026 | 1 | 113933371 | 114228545 | 1 | 1500860 | 5.6767 | 6.8657E-09 | MAGI3 | vte_hashimoto |
| ENSG00000143153 | 1 | 169074935 | 169101960 | 2 | 1500860 | 5.3885 | 3.5518E-08 | ATP1B1 | vte_hashimoto |
| ENSG00000143156 | 1 | 169101769 | 169337205 | 2 | 1500860 | 6.1094 | 5E-10 | NME7 | vte_hashimoto |
| ENSG00000117477 | 1 | 169364108 | 169429907 | 3 | 1500860 | 6.2602 | 1.9225E-10 | CCDC181 | vte_hashimoto |
| ENSG00000117479 | 1 | 169433147 | 169455241 | 1 | 1500860 | 5.8344 | 2.6986E-09 | SLC19A2 | vte_hashimoto |
| ENSG00000198734 | 1 | 169483404 | 169555826 | 15 | 1500860 | 5.546 | 1.4618E-08 | F5 | vte_hashimoto |
| ENSG00000174175 | 1 | 169558087 | 169599431 | 1 | 1500860 | 5.6648 | 7.3602E-09 | SELP | vte_hashimoto |
| ENSG00000000460 | 1 | 169631245 | 169823221 | 1 | 1500860 | 6.7168 | 9.2856E-12 | C1orf112 | vte_hashimoto |
| ENSG00000000457 | 1 | 169818772 | 169863408 | 1 | 1500860 | 5.3665 | 4.0144E-08 | SCYL3 | vte_hashimoto |
| ENSG00000115718 | 2 | 128176003 | 128186822 | 1 | 1500860 | 6.0087 | 9.3504E-10 | PROC | vte_hashimoto |
| ENSG00000164344 | 4 | 187130133 | 187179625 | 9 | 1500860 | 5.1592 | 1.2397E-07 | KLKB1 | vte_hashimoto |
| ENSG00000088926 | 4 | 187187099 | 187210835 | 7 | 1500860 | 7.8659 | 1.8319E-15 | F11 | vte_hashimoto |
| ENSG00000272297 | 4 | 187347700 | 187476464 | 10 | 1500860 | 6.7838 | 5.8538E-12 | RP11-215A1 | vte_hashimoto |
| ENSG00000204344 | 6 | 31938868 | 31950598 | 1 | 1500860 | 5.5966 | 1.0927E-08 | STK19 | vte_hashimoto |
| ENSG00000196735 | 6 | 32595956 | 32614839 | 1 | 1500860 | 5.3783 | 3.7601E-08 | HLA-DQA1 | vte_hashimoto |
| ENSG00000179344 | 6 | 32627244 | 32636160 | 1 | 1500860 | 6.0068 | 9.4634E-10 | HLA-DQB1 | vte_hashimoto |
| ENSG00000148297 | 9 | 136205160 | 136214986 | 1 | 1500860 | 6.3673 | 9.6209E-11 | MED22 | vte_hashimoto |
| ENSG00000198870 | 9 | 136243117 | 136271220 | 1 | 1500860 | 5.449 | 2.5322E-08 | C9orf96 | vte_hashimoto |
| ENSG00000160323 | 9 | 136279478 | 136324508 | 2 | 1500860 | 6.1094 | 5E-10 | ADAMTS13 | vte_hashimoto |
| ENSG00000160326 | 9 | 136336217 | 136344259 | 2 | 1500860 | 6.1094 | 5E-10 | SLC2A6 | vte_hashimoto |
| ENSG00000180210 | 11 | 46740730 | 46761056 | 1 | 1500860 | 5.9226 | 1.584E-09 | F2 | vte_hashimoto |
| ENSG00000175216 | 11 | 46764598 | 46867847 | 6 | 1500860 | 5.1173 | 1.55E-07 | CKAP5 | vte_hashimoto |
| ENSG00000134569 | 11 | 46878419 | 46940193 | 6 | 1500860 | 5.9607 | 1.2557E-09 | LRP4 | vte_hashimoto |
| ENSG00000149179 | 11 | 46958240 | 47185936 | 3 | 1500860 | 5.7603 | 4.1976E-09 | C11orf49 | vte_hashimoto |
| ENSG00000198324 | 12 | 111798455 | 111806925 | 1 | 1500860 | 6.4013 | 7.7019E-11 | FAM109A | vte_hashimoto |
| ENSG00000111252 | 12 | 111843752 | 111889427 | 1 | 1500860 | 10.686 | 5.9014E-27 | SH2B3 | vte_hashimoto |
| ENSG00000204842 | 12 | 111890018 | 112037480 | 2 | 1500860 | 6.1094 | 5E-10 | ATXN2 | vte_hashimoto |
| ENSG00000111300 | 12 | 112464500 | 112546826 | 1 | 1500860 | 8.7899 | 7.4868E-19 | NAA25 | vte_hashimoto |
| ENSG00000173064 | 12 | 112597992 | 112819896 | 1 | 1500860 | 9.0156 | 9.7908E-20 | HECTD4 | vte_hashimoto |
| ENSG00000179295 | 12 | 112856155 | 112947717 | 2 | 1500860 | 6.1094 | 5E-10 | PTPN11 | vte_hashimoto |

| | | | | | | | | | |
|---|---|---|---|---|---|---|---|---|---|
| ENSG00000197943 | 16 | 81772702 | 81991899 | 1 | 1500860 | 5.4653 | 2.3105E-08 | PLCG2 | vte_hashimoto |
| ENSG00000070366 | 17 | 1963133 | 2207065 | 14 | 1500860 | 6.9416 | 1.9386E-12 | SMG6 | vte_hashimoto |
| ENSG00000129353 | 19 | 10713133 | 10755235 | 7 | 1500860 | 4.9358 | 3.9905E-07 | SLC44A2 | vte_hashimoto |
| ENSG00000078747 | 20 | 32951041 | 33099198 | 1 | 1500860 | 5.4087 | 3.175E-08 | ITCH | vte_hashimoto |
| ENSG00000088298 | 20 | 33703167 | 33865928 | 5 | 1500860 | 4.6771 | 1.4551E-06 | EDEM2 | vte_hashimoto |
| ENSG00000101000 | 20 | 33759876 | 33765165 | 1 | 1500860 | 5.5278 | 1.6217E-08 | PROCR | vte_hashimoto |
| ENSG00000143153 | 1 | 169074935 | 169101960 | 2 | 1500860 | 6.1094 | 5E-10 | ATP1B1 | vte_psoriasis |
| ENSG00000143156 | 1 | 169101769 | 169337205 | 2 | 1500860 | 6.1094 | 5E-10 | NME7 | vte_psoriasis |
| ENSG00000117477 | 1 | 169364108 | 169429907 | 1 | 1500860 | 9.5425 | 6.9688E-22 | CCDC181 | vte_psoriasis |
| ENSG00000198734 | 1 | 169483404 | 169555826 | 22 | 1500860 | 6.1094 | 5E-10 | F5 | vte_psoriasis |
| ENSG00000088926 | 4 | 187187099 | 187210835 | 1 | 1500860 | 5.9647 | 1.2257E-09 | F11 | vte_psoriasis |
| ENSG00000204540 | 6 | 31082527 | 31107869 | 24 | 1500860 | 7.7806 | 3.6082E-15 | PSORS1C1 | vte_psoriasis |
| ENSG00000204539 | 6 | 31082867 | 31088223 | 8 | 1500860 | 6.2656 | 1.8565E-10 | CDSN | vte_psoriasis |
| ENSG00000204538 | 6 | 31105313 | 31107127 | 3 | 1500860 | 6.202 | 2.7876E-10 | PSORS1C2 | vte_psoriasis |
| ENSG00000234745 | 6 | 31321649 | 31324965 | 1 | 1500860 | 6.5205 | 3.5033E-11 | HLA-B | vte_psoriasis |
| ENSG00000204516 | 6 | 31462658 | 31478901 | 2 | 1500860 | 6.1094 | 5E-10 | MICB | vte_psoriasis |
| ENSG00000254870 | 6 | 31497996 | 31514385 | 2 | 1500860 | 6.1094 | 5E-10 | ATP6V1G2-I | vte_psoriasis |
| ENSG00000232810 | 6 | 31543344 | 31546113 | 2 | 1500860 | 6.1094 | 5E-10 | TNF | vte_psoriasis |
| ENSG00000204482 | 6 | 31553901 | 31556686 | 1 | 1500860 | 9.3307 | 5.258E-21 | LST1 | vte_psoriasis |
| ENSG00000204390 | 6 | 31777396 | 31783437 | 1 | 1500860 | 5.9745 | 1.1542E-09 | HSPA1L | vte_psoriasis |
| ENSG00000204385 | 6 | 31830969 | 31846823 | 1 | 1500860 | 6.2348 | 2.2621E-10 | SLC44A4 | vte_psoriasis |
| ENSG00000244255 | 6 | 31895475 | 31919825 | 1 | 1500860 | 5.6811 | 6.6912E-09 | CFB | vte_psoriasis |
| ENSG00000243649 | 6 | 31895475 | 31919861 | 1 | 1500860 | 5.6811 | 6.6912E-09 | CFB | vte_psoriasis |
| ENSG00000204356 | 6 | 31919864 | 31926887 | 1 | 1500860 | 6.219 | 2.5011E-10 | NELFE | vte_psoriasis |
| ENSG00000204344 | 6 | 31938868 | 31950598 | 1 | 1500860 | 5.7 | 5.9887E-09 | STK19 | vte_psoriasis |
| ENSG00000168477 | 6 | 32008931 | 32083111 | 6 | 1500860 | 7.1819 | 3.4373E-13 | TNXB | vte_psoriasis |
| ENSG00000213676 | 6 | 32065953 | 32096030 | 3 | 1500860 | 6.2997 | 1.4914E-10 | ATF6B | vte_psoriasis |
| ENSG00000198870 | 9 | 136243117 | 136271220 | 1 | 1500860 | 6.3393 | 1.1537E-10 | C9orf96 | vte_psoriasis |
| ENSG00000160323 | 9 | 136279478 | 136324508 | 1 | 1500860 | 5.5697 | 1.2761E-08 | ADAMTS13 | vte_psoriasis |
| ENSG00000134824 | 11 | 61560452 | 61634826 | 1 | 1500860 | 5.8376 | 2.6486E-09 | FADS2 | vte_psoriasis |
| ENSG00000111252 | 12 | 111843752 | 111889427 | 1 | 1500860 | 6.8858 | 2.8739E-12 | SH2B3 | vte_psoriasis |
| ENSG00000204842 | 12 | 111890018 | 112037480 | 2 | 1500860 | 5.2094 | 9.4738E-08 | ATXN2 | vte_psoriasis |

| | | | | | | | | | |
|---|---|---|---|---|---|---|---|---|---|
| ENSG00000099364 | 16 | 30934376 | 30960104 | 10 | 1500860 | 5.5872 | 1.1537E-08 | FBXL19 | vte_psoriasis |
| ENSG00000099381 | 16 | 30968615 | 30996437 | 9 | 1500860 | 4.1151 | 1.9348E-05 | SETD1A | vte_psoriasis |
| ENSG00000099377 | 16 | 30996519 | 31000473 | 2 | 1500860 | 5.5819 | 1.1893E-08 | HSD3B7 | vte_psoriasis |
| ENSG00000099365 | 16 | 31000577 | 31021949 | 3 | 1500860 | 5.6837 | 6.591E-09 | STX1B | vte_psoriasis |
| ENSG00000197943 | 16 | 81772702 | 81991899 | 1 | 1500860 | 5.7503 | 4.4543E-09 | PLCG2 | vte_psoriasis |
| ENSG00000070366 | 17 | 1963133 | 2207065 | 8 | 1500860 | 5.6819 | 6.6587E-09 | SMG6 | vte_psoriasis |
| ENSG00000129353 | 19 | 10713133 | 10755235 | 10 | 1500860 | 6.5488 | 2.8993E-11 | SLC44A2 | vte_psoriasis |
| ENSG00000129351 | 19 | 10764937 | 10803093 | 7 | 1500860 | 4.5347 | 2.8847E-06 | ILF3 | vte_psoriasis |
| ENSG00000213339 | 19 | 10812106 | 10824113 | 2 | 1500860 | 6.1094 | 5E-10 | QTRT1 | vte_psoriasis |
| ENSG00000079805 | 19 | 10828755 | 10944164 | 4 | 1500860 | 6.9188 | 2.277E-12 | DNM2 | vte_psoriasis |
| ENSG00000131069 | 20 | 33459949 | 33515769 | 1 | 1500860 | 5.6011 | 1.065E-08 | ACSS2 | vte_psoriasis |
| ENSG00000078814 | 20 | 33563206 | 33590240 | 2 | 1500860 | 5.5905 | 1.1323E-08 | MYH7B | vte_psoriasis |
| ENSG00000100991 | 20 | 33590207 | 33680674 | 3 | 1500860 | 6.7649 | 6.6706E-12 | TRPC4AP | vte_psoriasis |
| ENSG00000088298 | 20 | 33703167 | 33865928 | 8 | 1500860 | 6.7917 | 5.5413E-12 | EDEM2 | vte_psoriasis |
| ENSG00000101000 | 20 | 33759876 | 33765165 | 1 | 1500860 | 6.4613 | 5.1896E-11 | PROCR | vte_psoriasis |
| ENSG00000159363 | 1 | 17312453 | 17338423 | 1 | 1500860 | 5.3636 | 4.0787E-08 | ATP13A2 | vte_ra |
| ENSG00000117118 | 1 | 17345217 | 17380665 | 1 | 1500860 | 6.0252 | 8.4445E-10 | SDHB | vte_ra |
| ENSG00000159339 | 1 | 17634690 | 17690499 | 3 | 1500860 | 3.8075 | 7.0188E-05 | PADI4 | vte_ra |
| ENSG00000269468 | 1 | 17655507 | 17676070 | 1 | 1500860 | 5.4561 | 2.4332E-08 | AC004824.2 | vte_ra |
| ENSG00000081026 | 1 | 113933371 | 114228545 | 11 | 1500860 | 5.487 | 2.0446E-08 | MAGI3 | vte_ra |
| ENSG00000116793 | 1 | 114239453 | 114302111 | 4 | 1500860 | 5.6904 | 6.3379E-09 | PHTF1 | vte_ra |
| ENSG00000081019 | 1 | 114304454 | 114355098 | 6 | 1500860 | 5.9511 | 1.3318E-09 | RSBN1 | vte_ra |
| ENSG00000134242 | 1 | 114356433 | 114414381 | 8 | 1500860 | 6.3592 | 1.0142E-10 | PTPN22 | vte_ra |
| ENSG00000118292 | 1 | 150240600 | 150253327 | 1 | 1500860 | 5.6443 | 8.2922E-09 | C1orf54 | vte_ra |
| ENSG00000159208 | 1 | 150254953 | 150259505 | 1 | 1500860 | 5.6157 | 9.7862E-09 | C1orf51 | vte_ra |
| ENSG00000163125 | 1 | 150335567 | 150449042 | 2 | 1500860 | 4.2949 | 8.7396E-06 | RPRD2 | vte_ra |
| ENSG00000143369 | 1 | 150480538 | 150486265 | 1 | 1500860 | 5.3697 | 3.9427E-08 | ECM1 | vte_ra |
| ENSG00000160712 | 1 | 154377669 | 154441926 | 6 | 1500860 | 5.8408 | 2.5975E-09 | IL6R | vte_ra |
| ENSG00000143226 | 1 | 161475220 | 161493803 | 2 | 1500860 | 6.1094 | 5E-10 | FCGR2A | vte_ra |
| ENSG00000143156 | 1 | 169101769 | 169337205 | 1 | 1500860 | 8.4108 | 2.0363E-17 | NME7 | vte_ra |
| ENSG00000117477 | 1 | 169364108 | 169429907 | 1 | 1500860 | 8.5434 | 6.5183E-18 | CCDC181 | vte_ra |
| ENSG00000198734 | 1 | 169483404 | 169555826 | 5 | 1500860 | 7.0032 | 1.2513E-12 | F5 | vte_ra |

| | | | | | | | | | |
|---|---|---|---|---|---|---|---|---|---|
| ENSG00000198369 | 2 | 65537985 | 65659771 | 14 | 1500860 | 6.4421 | 5.8932E-11 | SPRED2 | vte_ra |
| ENSG00000144218 | 2 | 100162323 | 100759201 | 18 | 1500860 | 7.428 | 5.5123E-14 | AFF3 | vte_ra |
| ENSG00000138378 | 2 | 191894302 | 192016322 | 2 | 1500860 | 5.2817 | 6.3998E-08 | STAT4 | vte_ra |
| ENSG00000162944 | 2 | 198432948 | 198540769 | 8 | 1500860 | 5.6246 | 9.2982E-09 | RFTN2 | vte_ra |
| ENSG00000222017 | 2 | 198557830 | 198639547 | 9 | 1500860 | 4.7285 | 1.1307E-06 | AC011997.1 | vte_ra |
| ENSG00000152430 | 2 | 198591603 | 198651486 | 6 | 1500860 | 5.7045 | 5.8329E-09 | BOLL | vte_ra |
| ENSG00000115896 | 2 | 198669426 | 199437305 | 24 | 1500860 | 5.6391 | 8.5471E-09 | PLCL1 | vte_ra |
| ENSG00000163599 | 2 | 204732509 | 204738683 | 1 | 1500860 | 5.3422 | 4.591E-08 | CTLA4 | vte_ra |
| ENSG00000196355 | 4 | 38628029 | 38666430 | 1 | 1500860 | 5.9411 | 1.4153E-09 | AC021860.1 | vte_ra |
| ENSG00000109787 | 4 | 38665817 | 38702663 | 1 | 1500860 | 5.4455 | 2.5826E-08 | KLF3 | vte_ra |
| ENSG00000171560 | 4 | 155504278 | 155511918 | 2 | 1500860 | 6.1094 | 5E-10 | FGA | vte_ra |
| ENSG00000171557 | 4 | 155525286 | 155534119 | 2 | 1500860 | 6.1094 | 5E-10 | FGG | vte_ra |
| ENSG00000145476 | 4 | 187112674 | 187134610 | 6 | 1500860 | 5.2617 | 7.1376E-08 | CYP4V2 | vte_ra |
| ENSG00000164344 | 4 | 187130133 | 187179625 | 4 | 1500860 | 5.7733 | 3.887E-09 | KLKB1 | vte_ra |
| ENSG00000088926 | 4 | 187187099 | 187210835 | 7 | 1500860 | 7.6431 | 1.0603E-14 | F11 | vte_ra |
| ENSG00000043143 | 5 | 133860003 | 133918918 | 3 | 1500860 | 5.7038 | 5.8599E-09 | JADE2 | vte_ra |
| ENSG00000187626 | 6 | 28212401 | 28227011 | 2 | 1500860 | 5.9918 | 1.0377E-09 | ZKSCAN4 | vte_ra |
| ENSG00000137338 | 6 | 28249314 | 28270326 | 3 | 1500860 | 6.1094 | 5E-10 | PGBD1 | vte_ra |
| ENSG00000204542 | 6 | 31079000 | 31080336 | 1 | 1500860 | 7.9129 | 1.2575E-15 | C6orf15 | vte_ra |
| ENSG00000204540 | 6 | 31082527 | 31107869 | 15 | 1500860 | 7.7259 | 5.5511E-15 | PSORS1C1 | vte_ra |
| ENSG00000204539 | 6 | 31082867 | 31088223 | 1 | 1500860 | 7.1442 | 4.5277E-13 | CDSN | vte_ra |
| ENSG00000204538 | 6 | 31105313 | 31107127 | 2 | 1500860 | 5.5172 | 1.7221E-08 | PSORS1C2 | vte_ra |
| ENSG00000234745 | 6 | 31321649 | 31324965 | 1 | 1500860 | 5.7997 | 3.3216E-09 | HLA-B | vte_ra |
| ENSG00000204516 | 6 | 31462658 | 31478901 | 2 | 1500860 | 6.1094 | 5E-10 | MICB | vte_ra |
| ENSG00000254870 | 6 | 31497996 | 31514385 | 2 | 1500860 | 6.1094 | 5E-10 | ATP6V1G2-I | vte_ra |
| ENSG00000232810 | 6 | 31543344 | 31546113 | 2 | 1500860 | 5.8343 | 2.7001E-09 | TNF | vte_ra |
| ENSG00000204482 | 6 | 31553901 | 31556686 | 1 | 1500860 | 5.8072 | 3.1762E-09 | LST1 | vte_ra |
| ENSG00000204475 | 6 | 31556672 | 31560762 | 2 | 1500860 | 5.6455 | 8.2366E-09 | NCR3 | vte_ra |
| ENSG00000204410 | 6 | 31707725 | 31732622 | 3 | 1500860 | 6.3122 | 1.3753E-10 | MSH5 | vte_ra |
| ENSG00000255152 | 6 | 31707797 | 31732628 | 2 | 1500860 | 6.1094 | 5E-10 | MSH5-SAPC | vte_ra |
| ENSG00000204390 | 6 | 31777396 | 31783437 | 2 | 1500860 | 6.1094 | 5E-10 | HSPA1L | vte_ra |
| ENSG00000204385 | 6 | 31830969 | 31846823 | 12 | 1500860 | 7.6733 | 8.3822E-15 | SLC44A4 | vte_ra |

| | | | | | | | | | |
|---|---|---|---|---|---|---|---|---|---|
| ENSG00000204371 | 6 | 31847536 | 31865464 | 1 | 1500860 | 7.3986 | 6.8839E-14 | EHMT2 | vte_ra |
| ENSG00000166278 | 6 | 31865562 | 31913449 | 3 | 1500860 | 6.1094 | 5E-10 | C2 | vte_ra |
| ENSG00000244255 | 6 | 31895475 | 31919825 | 6 | 1500860 | 7.6203 | 1.2657E-14 | CFB | vte_ra |
| ENSG00000243649 | 6 | 31895475 | 31919861 | 6 | 1500860 | 7.6203 | 1.2657E-14 | CFB | vte_ra |
| ENSG00000204356 | 6 | 31919864 | 31926887 | 1 | 1500860 | 5.9366 | 1.455E-09 | NELFE | vte_ra |
| ENSG00000204344 | 6 | 31938868 | 31950598 | 1 | 1500860 | 9.7825 | 6.6922E-23 | STK19 | vte_ra |
| ENSG00000168477 | 6 | 32008931 | 32083111 | 16 | 1500860 | 7.0647 | 8.0497E-13 | TNXB | vte_ra |
| ENSG00000213676 | 6 | 32065953 | 32096030 | 7 | 1500860 | 7.845 | 2.1649E-15 | ATF6B | vte_ra |
| ENSG00000196735 | 6 | 32595956 | 32614839 | 4 | 1500860 | 6.1094 | 5E-10 | HLA-DQA1 | vte_ra |
| ENSG00000179344 | 6 | 32627244 | 32636160 | 4 | 1500860 | 6.961 | 1.6898E-12 | HLA-DQB1 | vte_ra |
| ENSG00000241106 | 6 | 32780540 | 32784825 | 2 | 1500860 | 5.4364 | 2.7182E-08 | HLA-DOB | vte_ra |
| ENSG00000250264 | 6 | 32781544 | 32806599 | 6 | 1500860 | 8.0766 | 3.3307E-16 | TAP2 | vte_ra |
| ENSG00000204267 | 6 | 32789610 | 32806557 | 4 | 1500860 | 6.8323 | 4.1795E-12 | TAP2 | vte_ra |
| ENSG00000204264 | 6 | 32808494 | 32812480 | 1 | 1500860 | 5.354 | 4.3025E-08 | PSMB8 | vte_ra |
| ENSG00000240065 | 6 | 32811913 | 32827362 | 1 | 1500860 | 5.5312 | 1.5906E-08 | PSMB9 | vte_ra |
| ENSG00000168394 | 6 | 32812986 | 32821755 | 1 | 1500860 | 5.5312 | 1.5906E-08 | TAP1 | vte_ra |
| ENSG00000248993 | 6 | 32905141 | 32920899 | 1 | 1500860 | 6.6453 | 1.5132E-11 | XXbac-BPG: | vte_ra |
| ENSG00000204252 | 6 | 32971955 | 32977389 | 3 | 1500860 | 6.1094 | 5E-10 | HLA-DOA | vte_ra |
| ENSG00000204248 | 6 | 33130458 | 33160276 | 1 | 1500860 | 5.6741 | 6.9699E-09 | COL11A2 | vte_ra |
| ENSG00000204231 | 6 | 33161365 | 33168630 | 1 | 1500860 | 5.7588 | 4.2355E-09 | RXRB | vte_ra |
| ENSG00000231925 | 6 | 33267471 | 33282164 | 1 | 1500860 | 6.2152 | 2.5623E-10 | TAPBP | vte_ra |
| ENSG00000153814 | 7 | 27870192 | 28220362 | 6 | 1500860 | 6.1094 | 5E-10 | JAZF1 | vte_ra |
| ENSG00000105810 | 7 | 92234235 | 92465908 | 17 | 1500860 | 7.2122 | 2.7528E-13 | CDK6 | vte_ra |
| ENSG00000136573 | 8 | 11351510 | 11422113 | 2 | 1500860 | 5.5644 | 1.3152E-08 | BLK | vte_ra |
| ENSG00000171102 | 9 | 136080664 | 136084630 | 1 | 1500860 | 6.3409 | 1.1423E-10 | OBP2B | vte_ra |
| ENSG00000198870 | 9 | 136243117 | 136271220 | 1 | 1500860 | 7.7046 | 6.5604E-15 | C9orf96 | vte_ra |
| ENSG00000160323 | 9 | 136279478 | 136324508 | 1 | 1500860 | 9.8939 | 2.2131E-23 | ADAMTS13 | vte_ra |
| ENSG00000160325 | 9 | 136325089 | 136335970 | 1 | 1500860 | 9.9134 | 1.8208E-23 | CACFD1 | vte_ra |
| ENSG00000160326 | 9 | 136336217 | 136344259 | 1 | 1500860 | 5.8348 | 2.6927E-09 | SLC2A6 | vte_ra |
| ENSG00000078403 | 10 | 21823094 | 22032559 | 1 | 1500860 | 5.3906 | 3.5105E-08 | MLLT10 | vte_ra |
| ENSG00000150347 | 10 | 63661059 | 63856703 | 1 | 1500860 | 6.3865 | 8.4846E-11 | ARID5B | vte_ra |
| ENSG00000175224 | 11 | 46638826 | 46696368 | 1 | 1500860 | 5.3661 | 4.0219E-08 | ATG13 | vte_ra |

| | | | | | | | | | |
|---|---|---|---|---|---|---|---|---|---|
| ENSG00000175213 | 11 | 46722368 | 46727462 | 2 | 1500860 | 6.1364 | 4.2194E-10 | ZNF408 | vte_ra |
| ENSG00000175216 | 11 | 46764598 | 46867847 | 1 | 1500860 | 5.4681 | 2.2741E-08 | CKAP5 | vte_ra |
| ENSG00000134569 | 11 | 46878419 | 46940193 | 1 | 1500860 | 5.4179 | 3.0147E-08 | LRP4 | vte_ra |
| ENSG00000124920 | 11 | 61520114 | 61555990 | 4 | 1500860 | 4.6591 | 1.5876E-06 | MYRF | vte_ra |
| ENSG00000134825 | 11 | 61535973 | 61560274 | 6 | 1500860 | 6.271 | 1.794E-10 | TMEM258 | vte_ra |
| ENSG00000168496 | 11 | 61560109 | 61564716 | 1 | 1500860 | 5.8142 | 3.0456E-09 | FEN1 | vte_ra |
| ENSG00000134824 | 11 | 61560452 | 61634826 | 14 | 1500860 | 5.7641 | 4.1039E-09 | FADS2 | vte_ra |
| ENSG00000149485 | 11 | 61567099 | 61596790 | 8 | 1500860 | 6.8361 | 4.0703E-12 | FADS1 | vte_ra |
| ENSG00000221968 | 11 | 61640991 | 61659523 | 1 | 1500860 | 5.4557 | 2.4391E-08 | FADS3 | vte_ra |
| ENSG00000111252 | 12 | 111843752 | 111889427 | 2 | 1500860 | 6.1094 | 5E-10 | SH2B3 | vte_ra |
| ENSG00000204842 | 12 | 111890018 | 112037480 | 2 | 1500860 | 6.1094 | 5E-10 | ATXN2 | vte_ra |
| ENSG00000111300 | 12 | 112464500 | 112546826 | 1 | 1500860 | 6.5476 | 2.923E-11 | NAA25 | vte_ra |
| ENSG00000173064 | 12 | 112597992 | 112819896 | 1 | 1500860 | 6.7478 | 7.5048E-12 | HECTD4 | vte_ra |
| ENSG00000179295 | 12 | 112856155 | 112947717 | 2 | 1500860 | 4.8077 | 7.6354E-07 | PTPN11 | vte_ra |
| ENSG00000172575 | 15 | 38780304 | 38857776 | 9 | 1500860 | 5.7156 | 5.4645E-09 | RASGRP1 | vte_ra |
| ENSG00000166949 | 15 | 67356101 | 67487533 | 1 | 1500860 | 5.341 | 4.6208E-08 | SMAD3 | vte_ra |
| ENSG00000140575 | 15 | 90931450 | 91045475 | 6 | 1500860 | 4.4511 | 4.2725E-06 | IQGAP1 | vte_ra |
| ENSG00000156860 | 16 | 30669752 | 30682135 | 2 | 1500860 | 4.2284 | 1.1767E-05 | FBRS | vte_ra |
| ENSG00000080603 | 16 | 30709530 | 30755602 | 3 | 1500860 | 4.3136 | 8.0294E-06 | SRCAP | vte_ra |
| ENSG00000103549 | 16 | 30773066 | 30787628 | 1 | 1500860 | 5.802 | 3.2771E-09 | RNF40 | vte_ra |
| ENSG00000099385 | 16 | 30844947 | 30906281 | 7 | 1500860 | 5.9895 | 1.0522E-09 | BCL7C | vte_ra |
| ENSG00000153774 | 16 | 75327596 | 75467383 | 14 | 1500860 | 5.2731 | 6.7087E-08 | CFDP1 | vte_ra |
| ENSG00000261717 | 16 | 75446582 | 75498604 | 6 | 1500860 | 6.4245 | 6.6153E-11 | RP11-77K12 | vte_ra |
| ENSG00000166822 | 16 | 75476952 | 75499395 | 4 | 1500860 | 6.1929 | 2.9538E-10 | TMEM170A | vte_ra |
| ENSG00000183196 | 16 | 75510949 | 75529282 | 1 | 1500860 | 5.4131 | 3.0963E-08 | CHST6 | vte_ra |
| ENSG00000129347 | 19 | 10663761 | 10676713 | 1 | 1500860 | 5.9515 | 1.3285E-09 | KRI1 | vte_ra |
| ENSG00000129353 | 19 | 10713133 | 10755235 | 14 | 1500860 | 7.845 | 2.1649E-15 | SLC44A2 | vte_ra |
| ENSG00000129351 | 19 | 10764937 | 10803093 | 9 | 1500860 | 6.9684 | 1.6024E-12 | ILF3 | vte_ra |
| ENSG00000079805 | 19 | 10828755 | 10944164 | 1 | 1500860 | 6.9606 | 1.6937E-12 | DNM2 | vte_ra |
| ENSG00000198646 | 20 | 33284722 | 33413452 | 6 | 1500860 | 5.9031 | 1.784E-09 | NCOA6 | vte_ra |
| ENSG00000131067 | 20 | 33432523 | 33460663 | 1 | 1500860 | 7.5703 | 1.8617E-14 | GGT7 | vte_ra |
| ENSG00000131069 | 20 | 33459949 | 33515769 | 18 | 1500860 | 6.1094 | 5E-10 | ACSS2 | vte_ra |

| | | | | | | | | | |
|---|---|---|---|---|---|---|---|---|---|
| ENSG00000100983 | 20 | 33516236 | 33543620 | 5 | 1500860 | 6.3733 | 9.2499E-11 | GSS | vte_ra |
| ENSG00000078814 | 20 | 33563206 | 33590240 | 10 | 1500860 | 7.7451 | 4.774E-15 | MYH7B | vte_ra |
| ENSG00000100991 | 20 | 33590207 | 33680674 | 12 | 1500860 | 6.8255 | 4.3821E-12 | TRPC4AP | vte_ra |
| ENSG00000088298 | 20 | 33703167 | 33865928 | 21 | 1500860 | 7.2702 | 1.7952E-13 | EDEM2 | vte_ra |
| ENSG00000101000 | 20 | 33759876 | 33765165 | 1 | 1500860 | 6.6819 | 1.179E-11 | PROCR | vte_ra |
| ENSG00000125966 | 20 | 33814457 | 33864801 | 2 | 1500860 | 5.1889 | 1.0574E-07 | MMP24 | vte_ra |
| ENSG00000143226 | 1 | 161475220 | 161493803 | 1 | 1500860 | 6.8407 | 3.9414E-12 | FCGR2A | vte_sle |
| ENSG00000143156 | 1 | 169101769 | 169337205 | 1 | 1500860 | 5.9367 | 1.4538E-09 | NME7 | vte_sle |
| ENSG00000138378 | 2 | 191894302 | 192016322 | 12 | 1500860 | 7.9284 | 1.1102E-15 | STAT4 | vte_sle |
| ENSG00000171560 | 4 | 155504278 | 155511918 | 1 | 1500860 | 7.7026 | 6.668E-15 | FGA | vte_sle |
| ENSG00000171557 | 4 | 155525286 | 155534119 | 1 | 1500860 | 7.3914 | 7.2629E-14 | FGG | vte_sle |
| ENSG00000145476 | 4 | 187112674 | 187134610 | 8 | 1500860 | 5.9115 | 1.6951E-09 | CYP4V2 | vte_sle |
| ENSG00000164344 | 4 | 187130133 | 187179625 | 9 | 1500860 | 5.1081 | 1.6271E-07 | KLKB1 | vte_sle |
| ENSG00000088926 | 4 | 187187099 | 187210835 | 4 | 1500860 | 6.7973 | 5.3298E-12 | F11 | vte_sle |
| ENSG00000204540 | 6 | 31082527 | 31107869 | 3 | 1500860 | 5.4141 | 3.0799E-08 | PSORS1C1 | vte_sle |
| ENSG00000179344 | 6 | 32627244 | 32636160 | 1 | 1500860 | 5.8737 | 2.1314E-09 | HLA-DQB1 | vte_sle |
| ENSG00000196821 | 6 | 34555065 | 34664636 | 11 | 1500860 | 6.7807 | 5.9784E-12 | C6orf106 | vte_sle |
| ENSG00000124562 | 6 | 34725183 | 34741571 | 1 | 1500860 | 6.6895 | 1.1195E-11 | SNRPC | vte_sle |
| ENSG00000065060 | 6 | 34759857 | 34850915 | 11 | 1500860 | 7.0069 | 1.2179E-12 | UHRF1BP1 | vte_sle |
| ENSG00000128604 | 7 | 128577666 | 128590089 | 1 | 1500860 | 5.6624 | 7.465E-09 | IRF5 | vte_sle |
| ENSG00000136573 | 8 | 11351510 | 11422113 | 8 | 1500860 | 6.0771 | 6.1205E-10 | BLK | vte_sle |
| ENSG00000134569 | 11 | 46878419 | 46940193 | 2 | 1500860 | 5.3521 | 4.3481E-08 | LRP4 | vte_sle |
| ENSG00000134825 | 11 | 61535973 | 61560274 | 1 | 1500860 | 5.6786 | 6.7911E-09 | TMEM258 | vte_sle |
| ENSG00000134824 | 11 | 61560452 | 61634826 | 3 | 1500860 | 5.5225 | 1.6714E-08 | FADS2 | vte_sle |
| ENSG00000149485 | 11 | 61567099 | 61596790 | 3 | 1500860 | 5.5225 | 1.6714E-08 | FADS1 | vte_sle |
| ENSG00000110799 | 12 | 6058040 | 6233936 | 1 | 1500860 | 5.3295 | 4.9233E-08 | VWF | vte_sle |
| ENSG00000099385 | 16 | 30844947 | 30906281 | 1 | 1500860 | 5.5376 | 1.5331E-08 | BCL7C | vte_sle |
| ENSG00000099364 | 16 | 30934376 | 30960104 | 8 | 1500860 | 4.3104 | 8.1489E-06 | FBXL19 | vte_sle |
| ENSG00000099381 | 16 | 30968615 | 30996437 | 9 | 1500860 | 4.3783 | 5.9816E-06 | SETD1A | vte_sle |
| ENSG00000099377 | 16 | 30996519 | 31000473 | 2 | 1500860 | 5.8685 | 2.1988E-09 | HSD3B7 | vte_sle |
| ENSG00000099365 | 16 | 31000577 | 31021949 | 2 | 1500860 | 5.0652 | 2.0398E-07 | STX1B | vte_sle |
| ENSG00000169896 | 16 | 31271311 | 31344213 | 1 | 1500860 | 5.3868 | 3.5854E-08 | ITGAM | vte_sle |

| | | | | | | | | | |
|---|---|---|---|---|---|---|---|---|---|
| ENSG00000129353 | 19 | 10713133 | 10755235 | 4 | 1500860 | 5.3248 | 5.0523E-08 | SLC44A2 | vte_sle |
| ENSG00000161179 | 22 | 21982378 | 21984353 | 2 | 1500860 | 5.2818 | 6.3951E-08 | YDJC | vte_sle |
| ENSG00000128274 | 22 | 43088127 | 43117304 | 1 | 1500860 | 6.0392 | 7.7428E-10 | A4GALT | vte_sle |
| ENSG00000160685 | 1 | 154975127 | 154990998 | 1 | 1500860 | 5.3774 | 3.7795E-08 | ZBTB7B | vte_ss |
| ENSG00000143153 | 1 | 169074935 | 169101960 | 8 | 1500860 | 6.4997 | 4.0249E-11 | ATP1B1 | vte_ss |
| ENSG00000143156 | 1 | 169101769 | 169337205 | 1 | 1500860 | 5.6736 | 6.9904E-09 | NME7 | vte_ss |
| ENSG00000198734 | 1 | 169483404 | 169555826 | 1 | 1500860 | 6.494 | 4.1807E-11 | F5 | vte_ss |
| ENSG00000145476 | 4 | 187112674 | 187134610 | 4 | 1500860 | 6.248 | 2.0789E-10 | CYP4V2 | vte_ss |
| ENSG00000164344 | 4 | 187130133 | 187179625 | 9 | 1500860 | 6.1094 | 5E-10 | KLKB1 | vte_ss |
| ENSG00000088926 | 4 | 187187099 | 187210835 | 5 | 1500860 | 6.3534 | 1.0531E-10 | F11 | vte_ss |
| ENSG00000079691 | 6 | 25279306 | 25620758 | 3 | 1500860 | 4.1691 | 1.5287E-05 | LRRC16A | vte_ss |
| ENSG00000146039 | 6 | 25754927 | 25781419 | 3 | 1500860 | 5.4173 | 3.0245E-08 | SLC17A4 | vte_ss |
| ENSG00000124568 | 6 | 25783125 | 25832287 | 1 | 1500860 | 5.58 | 1.2028E-08 | SLC17A1 | vte_ss |
| ENSG00000124564 | 6 | 25833294 | 25882514 | 1 | 1500860 | 5.8044 | 3.2298E-09 | SLC17A3 | vte_ss |
| ENSG00000112337 | 6 | 25912982 | 25930946 | 2 | 1500860 | 5.0146 | 2.6578E-07 | SLC17A2 | vte_ss |
| ENSG00000112343 | 6 | 25963030 | 25987384 | 3 | 1500860 | 5.8243 | 2.8668E-09 | TRIM38 | vte_ss |
| ENSG00000124610 | 6 | 26017260 | 26018040 | 1 | 1500860 | 5.6085 | 1.0205E-08 | HIST1H1A | vte_ss |
| ENSG00000010704 | 6 | 26087509 | 26098571 | 1 | 1500860 | 5.3322 | 4.8515E-08 | HFE | vte_ss |
| ENSG00000198315 | 6 | 28109688 | 28127250 | 1 | 1500860 | 5.3862 | 3.599E-08 | ZKSCAN8 | vte_ss |
| ENSG00000137338 | 6 | 28249314 | 28270326 | 1 | 1500860 | 5.5703 | 1.2718E-08 | PGBD1 | vte_ss |
| ENSG00000235109 | 6 | 28292470 | 28324048 | 2 | 1500860 | 6.1094 | 5E-10 | ZSCAN31 | vte_ss |
| ENSG00000189298 | 6 | 28317691 | 28336947 | 2 | 1500860 | 6.1094 | 5E-10 | ZKSCAN3 | vte_ss |
| ENSG00000158691 | 6 | 28346732 | 28367511 | 2 | 1500860 | 5.6135 | 9.9149E-09 | ZSCAN12 | vte_ss |
| ENSG00000187987 | 6 | 28399707 | 28411279 | 1 | 1500860 | 5.7705 | 3.9509E-09 | ZSCAN23 | vte_ss |
| ENSG00000204644 | 6 | 29640169 | 29648887 | 4 | 1500860 | 4.5016 | 3.3726E-06 | ZFP57 | vte_ss |
| ENSG00000204540 | 6 | 31082527 | 31107869 | 4 | 1500860 | 6.6704 | 1.2755E-11 | PSORS1C1 | vte_ss |
| ENSG00000204539 | 6 | 31082867 | 31088223 | 2 | 1500860 | 6.1094 | 5E-10 | CDSN | vte_ss |
| ENSG00000213760 | 6 | 31512239 | 31516204 | 1 | 1500860 | 5.5423 | 1.4929E-08 | ATP6V1G2 | vte_ss |
| ENSG00000204498 | 6 | 31514647 | 31526606 | 1 | 1500860 | 5.5423 | 1.4929E-08 | NFKBIL1 | vte_ss |
| ENSG00000226979 | 6 | 31539831 | 31542101 | 2 | 1500860 | 5.6197 | 9.5664E-09 | LTA | vte_ss |
| ENSG00000204385 | 6 | 31830969 | 31846823 | 8 | 1500860 | 4.7924 | 8.2387E-07 | SLC44A4 | vte_ss |
| ENSG00000204371 | 6 | 31847536 | 31865464 | 1 | 1500860 | 7.2879 | 1.5744E-13 | EHMT2 | vte_ss |

| | | | | | | | | | |
|---|---|---|---|---|---|---|---|---|---|
| ENSG00000244255 | 6 | 31895475 | 31919825 | 2 | 1500860 | 5.6178 | 9.67E-09 | CFB | vte_ss |
| ENSG00000243649 | 6 | 31895475 | 31919861 | 2 | 1500860 | 5.6178 | 9.67E-09 | CFB | vte_ss |
| ENSG00000204344 | 6 | 31938868 | 31950598 | 1 | 1500860 | 10.095 | 2.893E-24 | STK19 | vte_ss |
| ENSG00000196735 | 6 | 32595956 | 32614839 | 2 | 1500860 | 6.1094 | 5E-10 | HLA-DQA1 | vte_ss |
| ENSG00000179344 | 6 | 32627244 | 32636160 | 1 | 1500860 | 7.4734 | 3.9073E-14 | HLA-DQB1 | vte_ss |
| ENSG00000240065 | 6 | 32811913 | 32827362 | 2 | 1500860 | 5.2365 | 8.1846E-08 | PSMB9 | vte_ss |
| ENSG00000168394 | 6 | 32812986 | 32821755 | 2 | 1500860 | 5.2365 | 8.1846E-08 | TAP1 | vte_ss |
| ENSG00000160271 | 9 | 135973107 | 136039301 | 2 | 1500860 | 4.6309 | 1.8204E-06 | RALGDS | vte_ss |
| ENSG00000148288 | 9 | 136028340 | 136039332 | 2 | 1500860 | 4.6309 | 1.8204E-06 | GBGT1 | vte_ss |
| ENSG00000148296 | 9 | 136197552 | 136203235 | 1 | 1500860 | 5.7122 | 5.5757E-09 | SURF6 | vte_ss |
| ENSG00000148290 | 9 | 136218610 | 136223552 | 1 | 1500860 | 5.3438 | 4.5514E-08 | SURF1 | vte_ss |
| ENSG00000198870 | 9 | 136243117 | 136271220 | 1 | 1500860 | 7.9362 | 1.0421E-15 | C9orf96 | vte_ss |
| ENSG00000160323 | 9 | 136279478 | 136324508 | 1 | 1500860 | 7.2055 | 2.8908E-13 | ADAMTS13 | vte_ss |
| ENSG00000160325 | 9 | 136325089 | 136335970 | 1 | 1500860 | 7.1158 | 5.5631E-13 | CACFD1 | vte_ss |
| ENSG00000160326 | 9 | 136336217 | 136344259 | 1 | 1500860 | 6.2404 | 2.1818E-10 | SLC2A6 | vte_ss |
| ENSG00000149179 | 11 | 46958240 | 47185936 | 6 | 1500860 | 4.2126 | 1.2624E-05 | C11orf49 | vte_ss |
| ENSG00000099385 | 16 | 30844947 | 30906281 | 4 | 1500860 | 5.6205 | 9.5216E-09 | BCL7C | vte_ss |
| ENSG00000225190 | 17 | 43513266 | 43568115 | 2 | 1500860 | 5.4975 | 1.9264E-08 | PLEKHM1 | vte_ss |
| ENSG00000127616 | 19 | 11071598 | 11176071 | 3 | 1500860 | 4.3083 | 8.224E-06 | SMARCA4 | vte_ss |
| ENSG00000088298 | 20 | 33703167 | 33865928 | 2 | 1500860 | 6.0312 | 8.1383E-10 | EDEM2 | vte_ss |

Table S3: Heterogeneity Tests

| Exposure trait | MR-Egger | | | IVW | | |
|---|---|---|---|---|---|---|
| | Cochran's Q | $I^2$(%) | p-value | Cochran's Q | $I^2$(%) | p-value |
| SLE | 1.93E+02 | 4.56E+01 | 3.69E-07 | 1.95E+02 | 4.55E+01 | 3.43E-07 |
| RA | 5.26E+02 | 6.20E+01 | 3.70E-31 | 5.69E+02 | 6.47E+01 | 6.69E-37 |
| HT | 1.51E+02 | 5.50E+01 | 2.92×10-8 | 1.51E+02 | 5.43E+01 | 4.41×10-8 |
| SS | 2.60E+01 | 1.86E+01 | 2.14E-01 | 3.20E+01 | 3.10E+01 | 7.92E-02 |
| psoriasis | 1.63E+02 | 6.63E+01 | 1.20E-12 | 1.74E+02 | 6.79E+01 | 4.93E-14 |

Abbreviations: IVW, inverse-variance weighted; MR, Mendelian randomization; VTE, venous thromboembolism; SLE, systemic lupus erythematosus; RA, rheumatoid arthritis; HT, Hashimoto's thyroiditis; SS, Sjögren's syndrome

### Supplementary Figure Legends

Supplementary Figure 1. Manhattan plots from the PLACO analysis across the Autoimmune–Thrombotic Axis. The blue line indicates the suggestive significance threshold ($1 \times 10^{-5}$), and the red line indicates the genome-wide significance threshold ($5 \times 10^{-8}$). (A) VTE & RA; (B) VTE & SLE; (C) VTE & SS; (D) VTE & psoriasis; (E) VTE & HT. Abbreviations: HT, Hashimoto's thyroiditis; PLACO, pleiotropy analysis and cross-phenotype analysis; RA, rheumatoid arthritis; SLE, systemic lupus erythematosus; SS, Sjögren's syndrome; VTE, venous thromboembolism.

Supplementary Figure 2. Manhattan plots from the CPASSOC analysis across the Autoimmune–Thrombotic Axis. The blue line indicates the suggestive significance threshold ($1 \times 10^{-5}$), and the red line indicates the genome-wide significance threshold ($5 \times 10^{-8}$). (A) VTE & RA; (B) VTE & SLE; (C) VTE & SS; (D) VTE & psoriasis; (E) VTE & HT. Abbreviations: CPASSOC, Cross-Phenotype Association Analysis; HT, Hashimoto's thyroiditis; RA, rheumatoid arthritis; SLE, systemic lupus erythematosus; SS, Sjögren's syndrome; VTE, venous thromboembolism.

Supplementary Figure 3. Regional association plots of ISLs between AD and VTE. (A–E) ISLs for RA and VTE located at 1q21.3, 2q11.2, 4p14, 7p15.1, and 7p21.2, respectively. (F) An ISL for HT and VTE at chr12q24.12–q24.13. All loci shown satisfy PP.H4 > 0.9, indicating strong evidence for a shared causal variant. Abbreviations: HT, Hashimoto’s thyroiditis; ISLs, Immunothrombotic Shared Loci; PP.H4, posterior probability for hypothesis 4 (colocalization); RA, rheumatoid arthritis; VTE, venous thromboembolism.

Supplementary Figure 4. Colocalization analysis across the Autoimmune–Thrombotic Axis: AD or VTE and eQTL of pleiotropic genes in whole blood tissue (GTEx). Abbreviations: AD, autoimmune disease; eQTL, expression quantitative trait locus; VTE, venous thromboembolism.

Supplementary Figure 5. Colocalization analysis across the Autoimmune–Thrombotic Axis: AD or VTE and eQTL of pleiotropic genes in whole blood tissue (eQTLgen). Abbreviations: AD, autoimmune disease; eQTL, expression quantitative trait locus; VTE, venous thromboembolism.

Supplementary Figure 6. Colocalization analysis across the Autoimmune–Thrombotic Axis: AD or VTE and eQTL of pleiotropic genes in thyroid tissue. Abbreviations: AD, autoimmune disease; eQTL, expression quantitative trait locus; VTE, venous thromboembolism.

Supplementary Figure 7. Colocalization analysis across the Autoimmune–Thrombotic Axis: AD or VTE and eQTL of pleiotropic genes in spleen tissue. Abbreviations: AD, autoimmune disease; eQTL, expression quantitative trait locus; VTE, venous thromboembolism.

Supplementary Figure 8. Colocalization analysis across the Autoimmune–Thrombotic Axis: AD or VTE and eQTL of pleiotropic genes in sun exposed (lower leg) skin. Abbreviations: AD, autoimmune disease; eQTL, expression quantitative trait locus; VTE, venous thromboembolism.

Supplementary Figure 9. Colocalization analysis across the Autoimmune–Thrombotic Axis: AD or VTE and eQTL of pleiotropic genes in sun unexposed (suprapubic) skin. Abbreviations: AD, autoimmune disease; eQTL, expression quantitative trait locus; VTE, venous thromboembolism.

Supplementary Figure 10. Colocalization analysis across the Autoimmune–Thrombotic Axis: AD or VTE and eQTL of pleiotropic genes in skeletal muscle. Abbreviations: AD, autoimmune disease; eQTL, expression quantitative trait locus; VTE, venous thromboembolism.

Supplementary Figure 11. Colocalization analysis across the Autoimmune–Thrombotic Axis: AD or VTE and eQTL of pleiotropic genes in EBV-transformed lymphocytes. Abbreviations: AD, autoimmune disease; eQTL, expression quantitative trait locus; VTE, venous thromboembolism.

Supplementary Figure 12. Colocalization analysis across the Autoimmune–Thrombotic Axis: AD or VTE and eQTL of pleiotropic genes in cultured fibroblasts. Abbreviations: AD, autoimmune disease; eQTL, expression quantitative trait locus; VTE, venous thromboembolism.

Supplementary Figure 13. Colocalization analysis across the Autoimmune–Thrombotic Axis: AD or VTE and eQTL of pleiotropic genes in artery (aorta). Abbreviations: AD, autoimmune disease; eQTL, expression quantitative trait locus; VTE, venous thromboembolism.

Supplementary Figure 14. Colocalization analysis across the Autoimmune–Thrombotic Axis: AD or VTE and eQTL of pleiotropic genes in artery (coronary). Abbreviations: AD, autoimmune disease; eQTL, expression quantitative trait locus; VTE, venous thromboembolism.

Supplementary Figure 15. Colocalization analysis across the Autoimmune–Thrombotic Axis: AD or VTE and eQTL of pleiotropic genes in artery (tibial). Abbreviations: AD, autoimmune disease; eQTL, expression quantitative trait locus; VTE, venous thromboembolism.

Supplementary Figure 16. Colocalization analysis of IL6R multi-tissue eQTL with traits across the Autoimmune–Thrombotic Axis. (A) Cells (EBV-transformed lymphocytes) eQTL with RA; (B) Artery–Aorta eQTL with RA; (C) Artery–Tibial eQTL with RA; (D) Artery–Coronary eQTL with RA; (E) Artery–Aorta eQTL with VTE. Abbreviations: eQTL, expression quantitative trait locus; RA, rheumatoid arthritis; VTE, venous thromboembolism.

Supplementary Figure 17. Colocalization analysis of PLCL1 multi-tissue eQTL with traits across the Autoimmune–Thrombotic Axis. (A) Spleen eQTL with RA; (B) Artery–Coronary eQTL with RA; (C) Whole Blood (eQTLGen) eQTL with RA; (D) Artery–Tibial eQTL with VTE; (E) Thyroid eQTL with VTE. Abbreviations: eQTL, expression quantitative trait locus; RA, rheumatoid arthritis; VTE, venous thromboembolism.

Supplementary Figure 18. Linkage Disequilibrium Heatmaps of the IL6R (A) and PLCL1 (B) Loci from Multi-Tissue eQTL Colocalization Analysis. The color scale represents the degree of linkage disequilibrium ($r^2$) between SNPs, with red indicating high $r^2$ (strong LD) and blue indicating low $r^2$ (weak LD). Abbreviations: LD, linkage disequilibrium.

Supplementary Figure 19. SMR analysis across the Autoimmune–Thrombotic Axis: VTE and eQTL from multiple tissues. (A) Aorta Artery eQTL from GTEx and VTE; (B) Coronary Artery eQTL from GTEx and VTE; (C) Tibial Artery eQTL from GTEx and VTE; (D) Skeletal Muscle eQTL from GTEx and VTE; (E) EBV-transformed lymphocytes eQTL from GTEx and VTE; (F) Spleen eQTL from GTEx and VTE; (G) Whole Blood eQTL from GTEx and VTE; (H) Whole Blood eQTL from eQTLGen and VTE. Abbreviations: eQTL, expression quantitative trait locus; GTEx, Genotype-Tissue Expression project; SMR, summary-based Mendelian randomization; VTE, venous thromboembolism.

Supplementary Figure 20. SMR analysis across the Autoimmune–Thrombotic Axis: RA and eQTL from multiple tissues. (A) Aorta Artery eQTL from GTEx and RA; (B) Coronary Artery eQTL from GTEx and RA; (C) Tibial Artery eQTL from GTEx and RA; (D) Skeletal Muscle eQTL from GTEx and RA; (E) EBV-transformed lymphocytes eQTL from GTEx and RA; (F) Spleen eQTL from GTEx and RA; (G) Whole Blood eQTL from GTEx and RA; (H) Whole Blood eQTL from eQTLGen and RA. Abbreviations: eQTL, expression quantitative trait locus; GTEx, Genotype-Tissue Expression project; RA, rheumatoid arthritis; SMR, summary-based Mendelian randomization.

Supplementary Figure 21. SMR analysis across the Autoimmune–Thrombotic Axis: SLE and eQTL from multiple tissues. (A) Aorta Artery eQTL from GTEx and SLE; (B) Coronary Artery eQTL from GTEx and SLE; (C) Tibial Artery eQTL from GTEx and SLE; (D) Skeletal Muscle eQTL from GTEx and SLE; (E) EBV-transformed lymphocytes eQTL from GTEx and SLE; (F) Spleen eQTL from GTEx and SLE; (G) Whole Blood eQTL from GTEx and SLE; (H) Whole Blood eQTL from eQTLGen and SLE. Abbreviations: eQTL, expression quantitative trait locus; GTEx, Genotype-Tissue Expression project; SLE, systemic lupus erythematosus; SMR, summary-based Mendelian randomization.

Supplementary Figure 22. SMR analysis across the Autoimmune–Thrombotic Axis: SS and eQTL from multiple tissues. (A) Aorta Artery eQTL from GTEx and SS; (B) Coronary Artery eQTL from GTEx and SS; (C) Tibial Artery eQTL from GTEx and SS; (D) Skeletal Muscle eQTL from GTEx and SS; (E) EBV-transformed lymphocytes eQTL from GTEx and SS; (F) Spleen eQTL from GTEx and SS; (G) Whole Blood eQTL from GTEx and SS; (H) Whole Blood eQTL from eQTLGen and SS. Abbreviations: eQTL, expression quantitative trait locus; GTEx, Genotype-Tissue Expression project; SMR, summary-based Mendelian randomization; SS, Sjögren's syndrome.

Supplementary Figure 23. SMR analysis across the Autoimmune–Thrombotic Axis: psoriasis and eQTL from multiple tissues. (A) Aorta Artery eQTL from GTEx and psoriasis; (B) Coronary Artery eQTL from GTEx and psoriasis; (C) Tibial Artery eQTL from GTEx and psoriasis; (D) Skeletal Muscle eQTL from GTEx and psoriasis; (E) EBV-transformed lymphocytes eQTL from GTEx and psoriasis; (F) Spleen eQTL from GTEx and psoriasis; (G) Whole Blood eQTL from GTEx and psoriasis; (H) Whole Blood eQTL from eQTLGen and psoriasis. Abbreviations: eQTL, expression quantitative trait locus; GTEx, Genotype-Tissue Expression project; SMR, summary-based Mendelian randomization.

Supplementary Figure 24. SMR analysis across the Autoimmune–Thrombotic Axis: HT and eQTL from multiple tissues. (A) Aorta Artery eQTL from GTEx and HT; (B) Coronary Artery eQTL from GTEx and HT; (C) Tibial Artery eQTL from GTEx and HT; (D) Skeletal Muscle eQTL from GTEx and HT; (E) EBV-transformed lymphocytes eQTL from GTEx and HT; (F) Spleen eQTL from GTEx and HT; (G) Whole Blood eQTL from GTEx and HT; (H) Whole Blood eQTL from eQTLGen and HT. Abbreviations: eQTL, expression quantitative trait locus; GTEx, Genotype-Tissue Expression project; HT, Hashimoto's thyroiditis; SMR, summary-based Mendelian randomization.

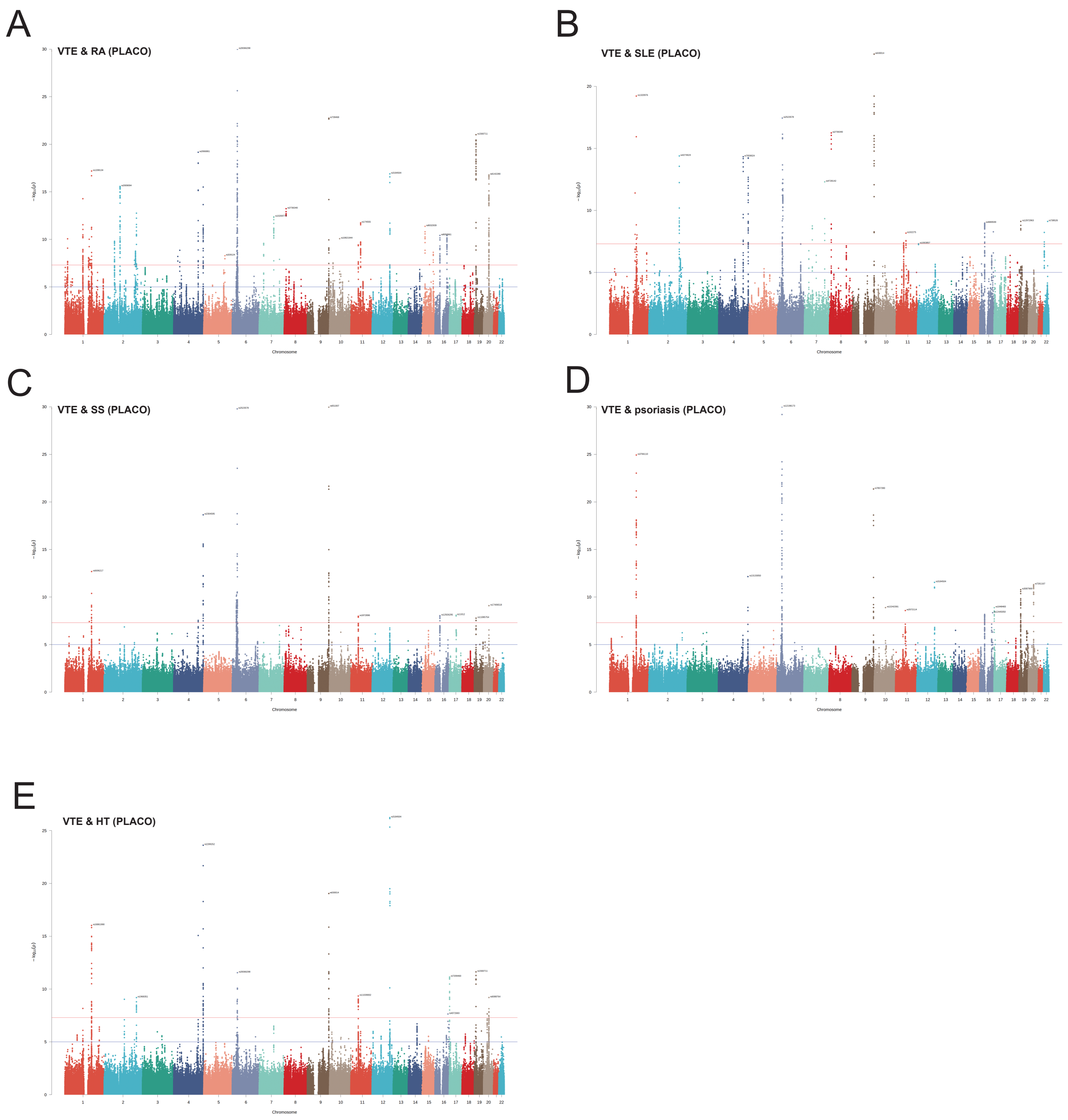


Supplementary Figure 1

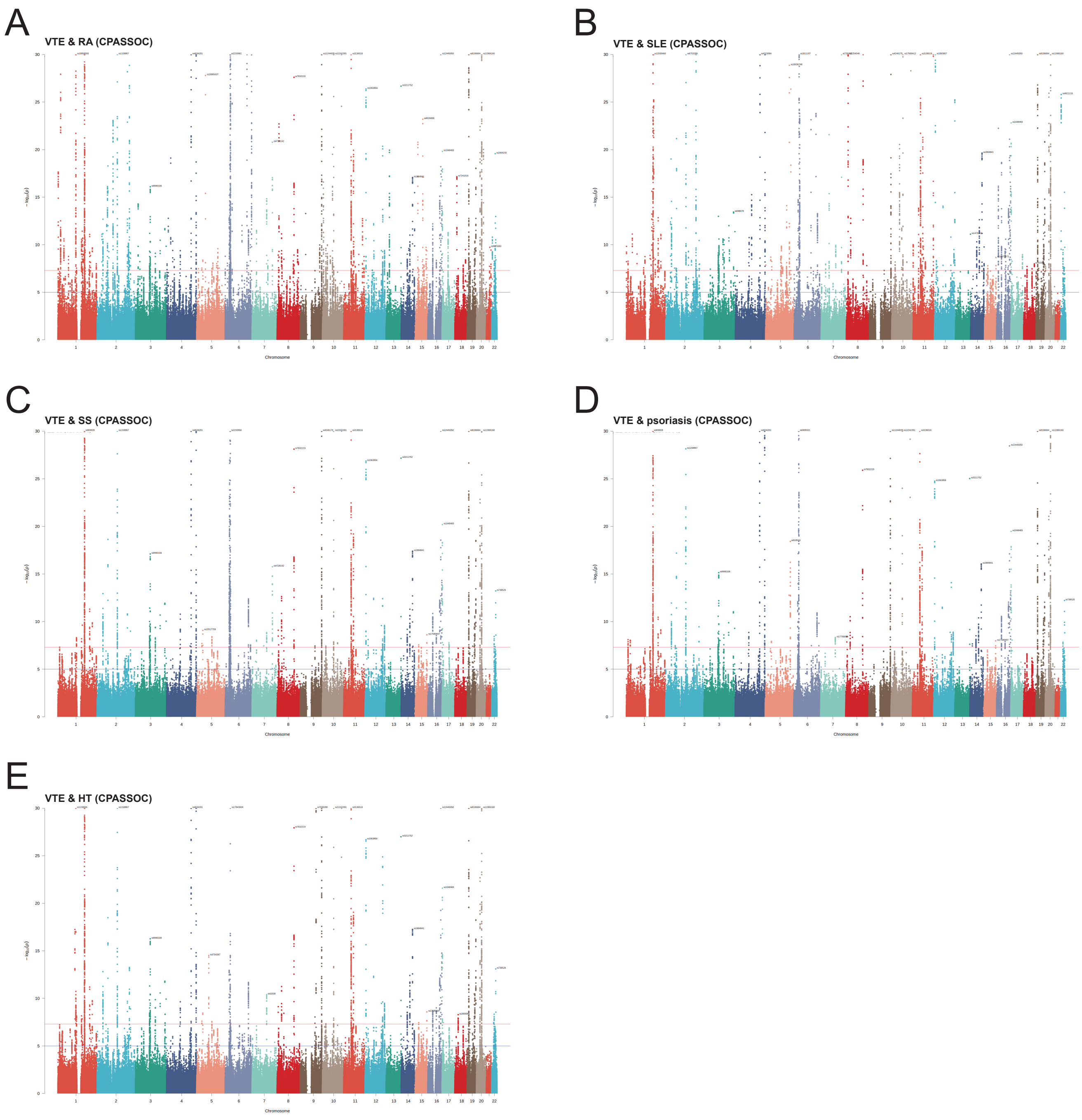


Supplementary Figure 2

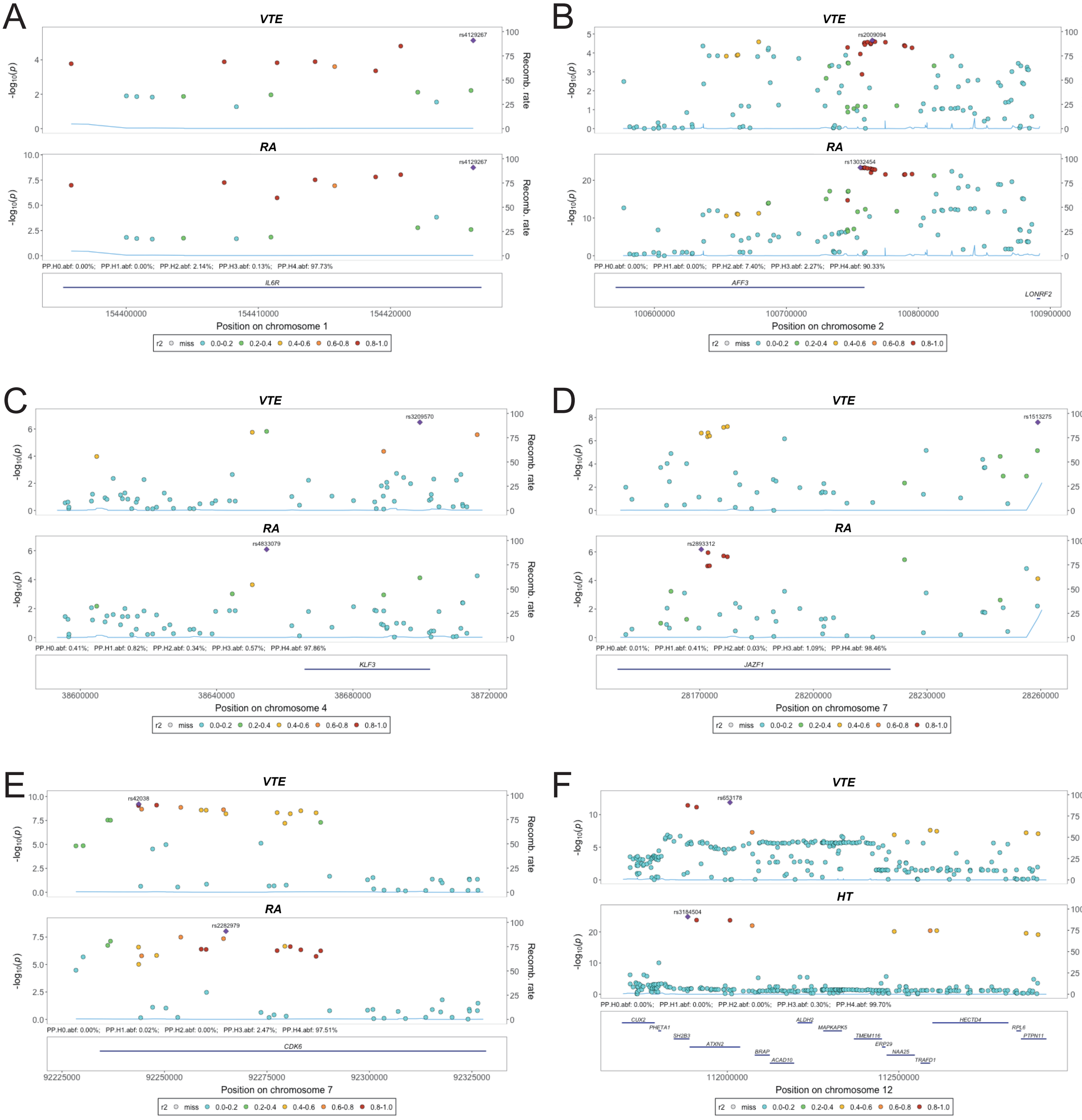


Supplementary Figure 3

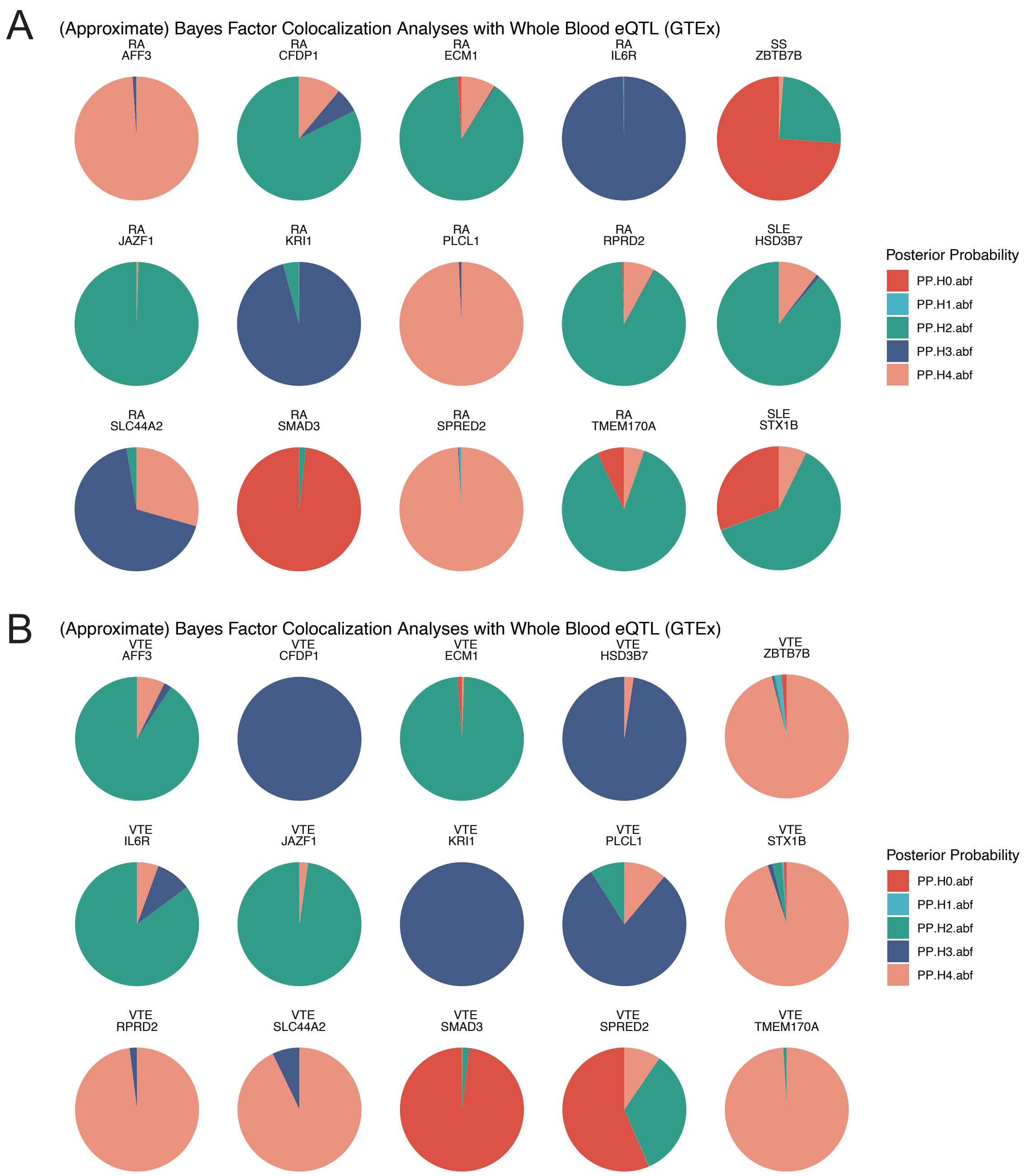


Supplementary Figure 4

A
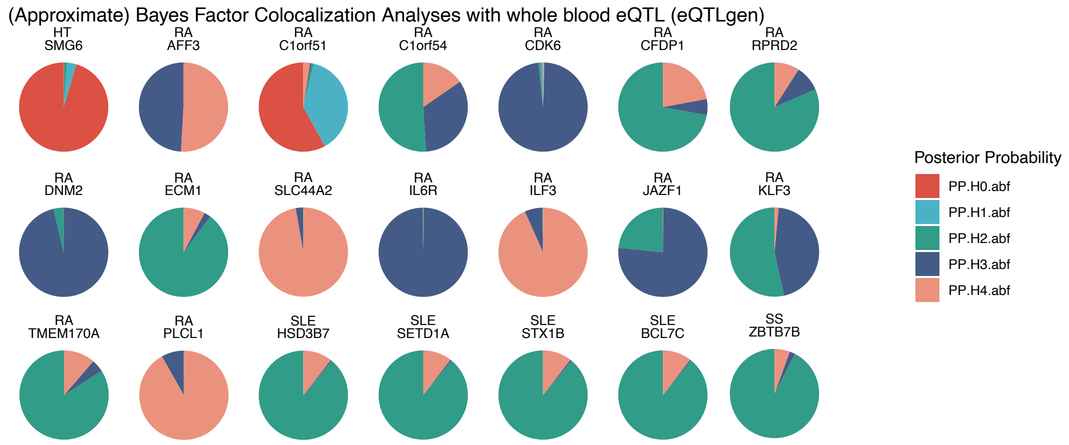


B
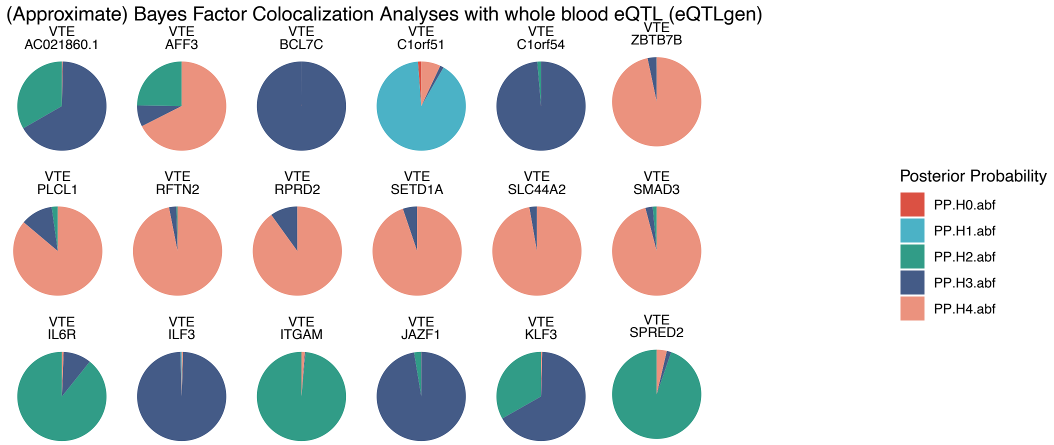


Supplementary Figure 5

A

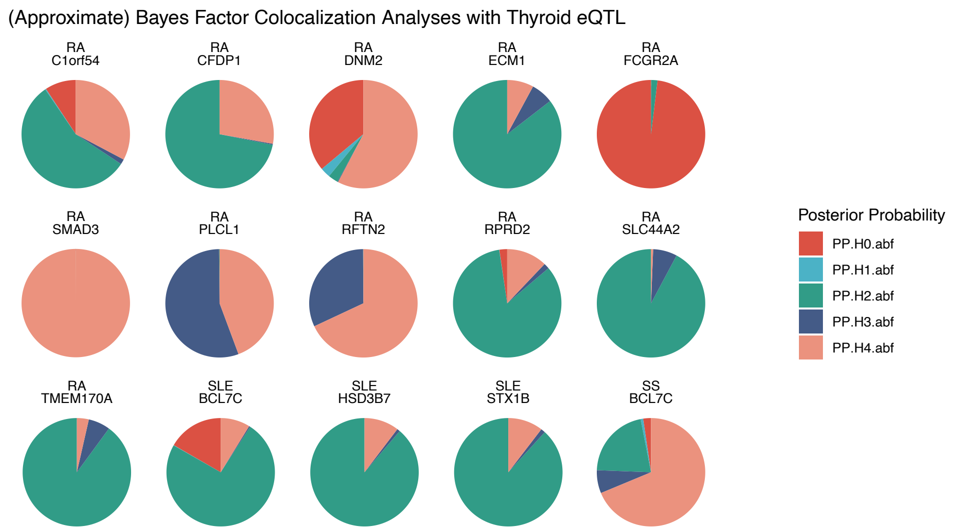


B

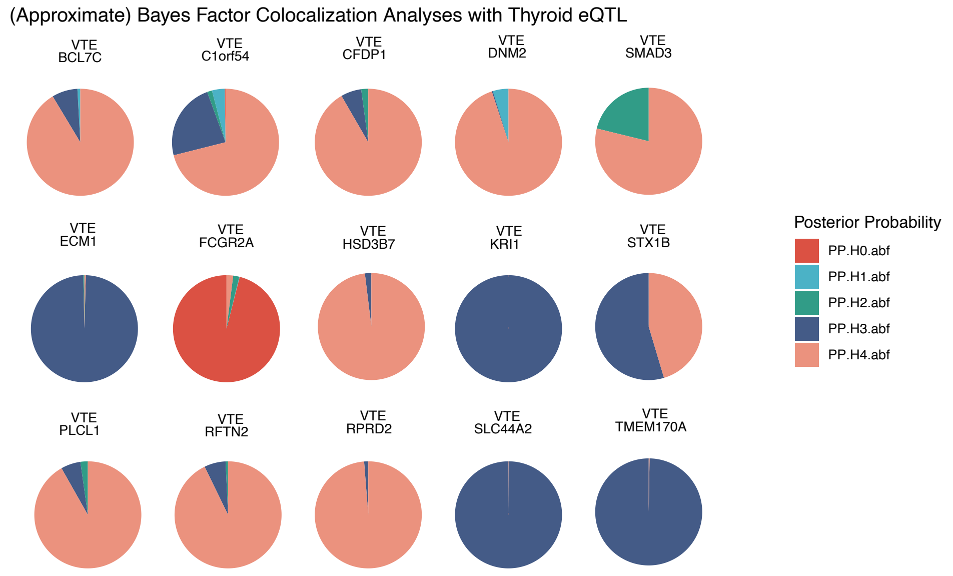


Supplementary Figure 6

A

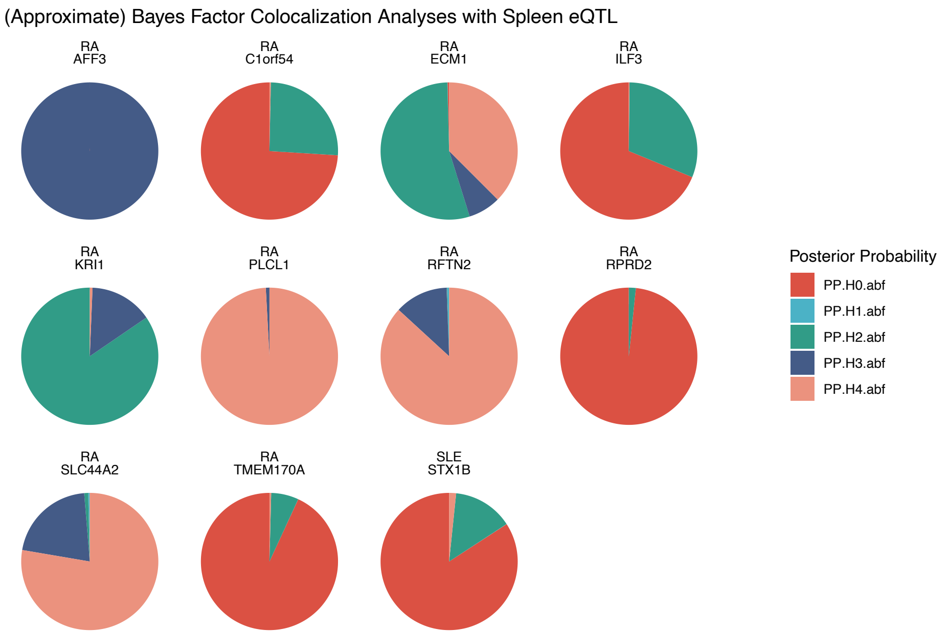


B

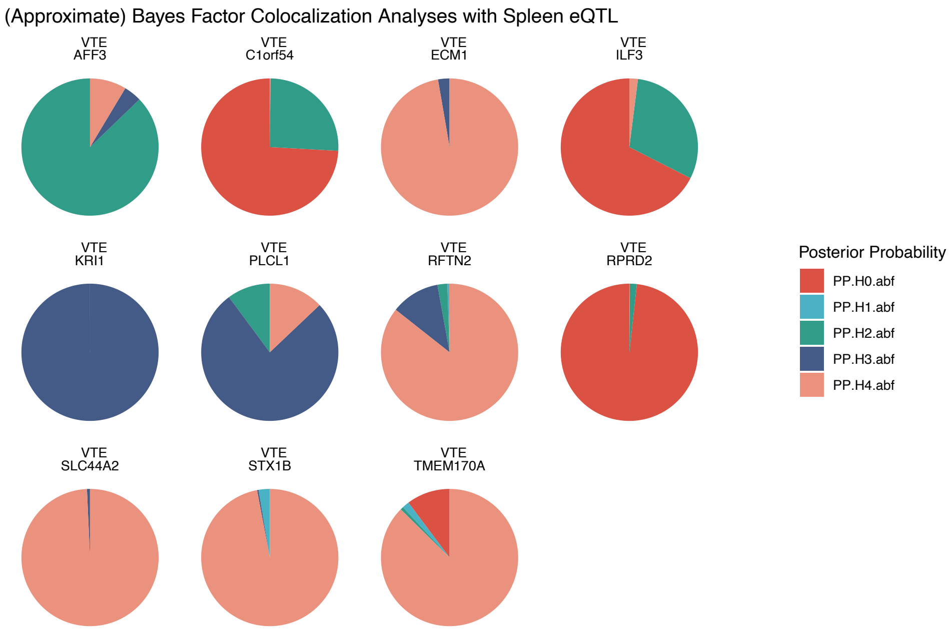


Supplementary Figure 7

A (Approximate) Bayes Factor Colocalization Analyses with Skin – Sun Exposed (Lower leg) eQTL

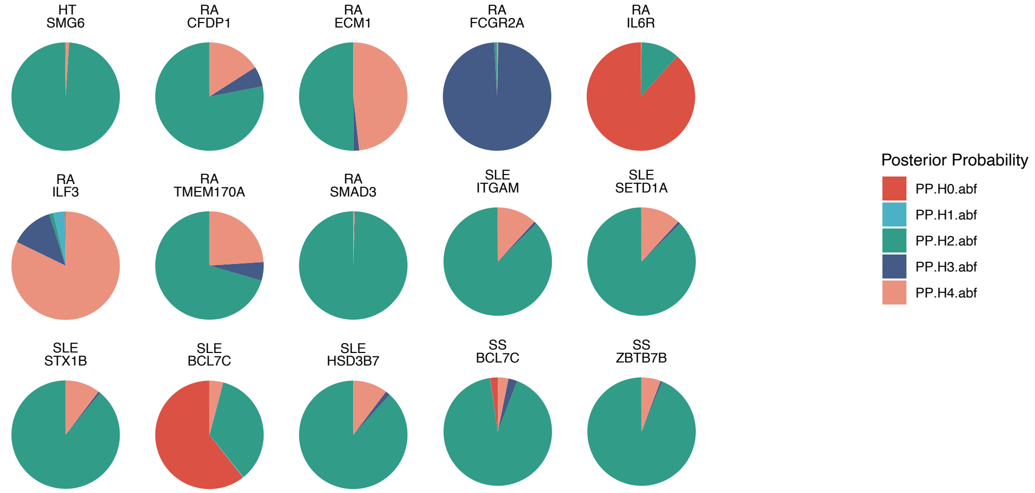


B (Approximate) Bayes Factor Colocalization Analyses with Skin – Sun Exposed (Lower leg) eQTL

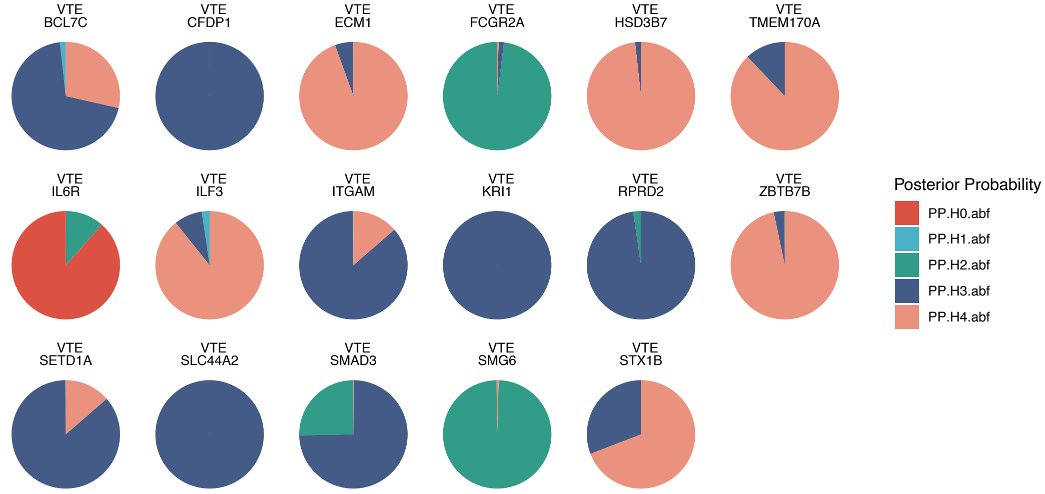


Supplementary Figure 8

A

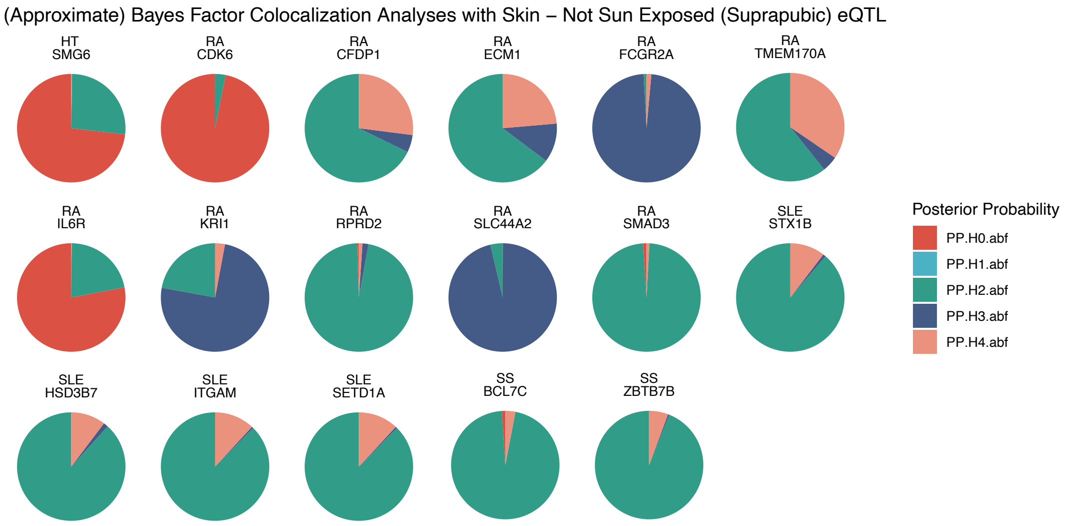


B

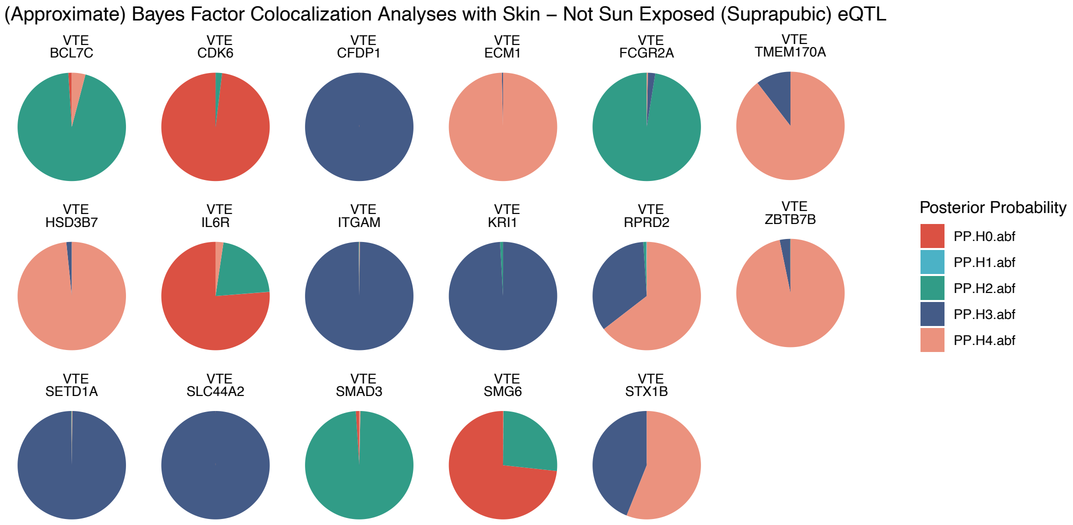


Supplementary Figure 9

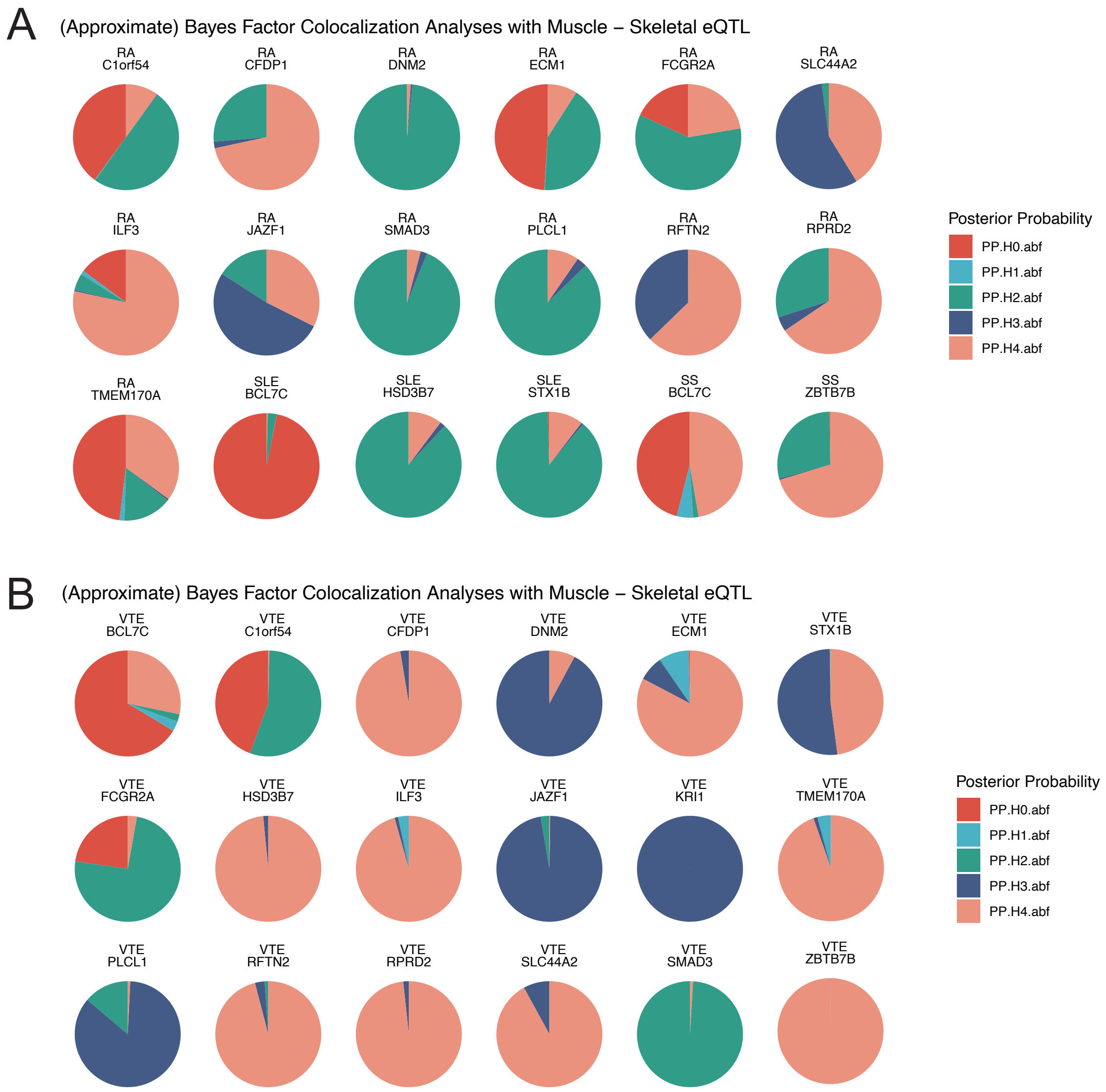


Supplementary Figure 10

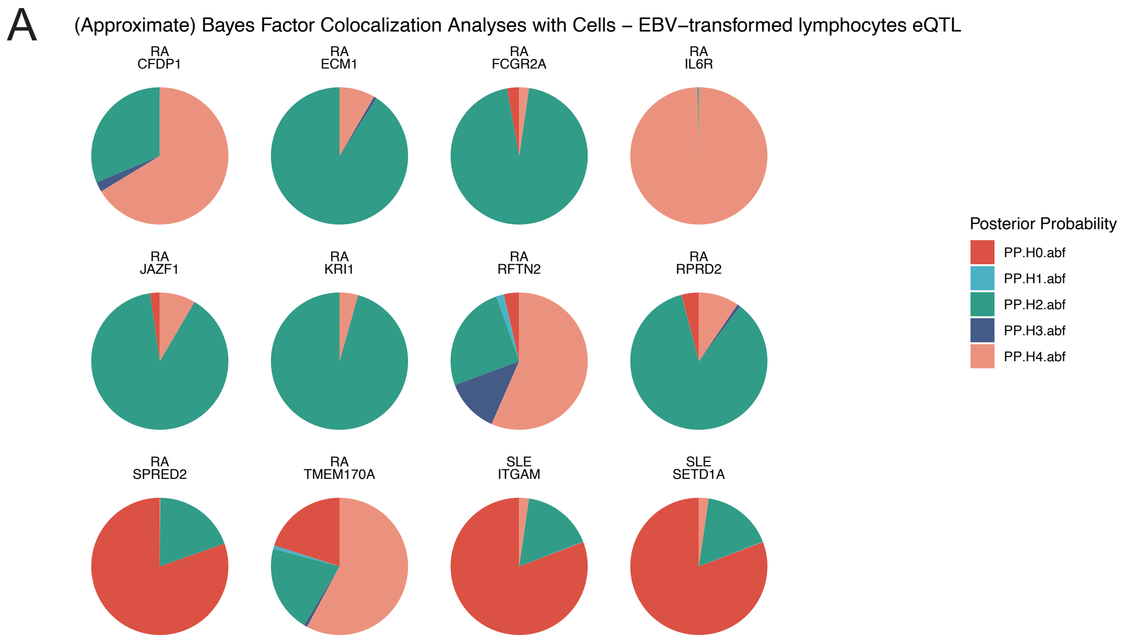


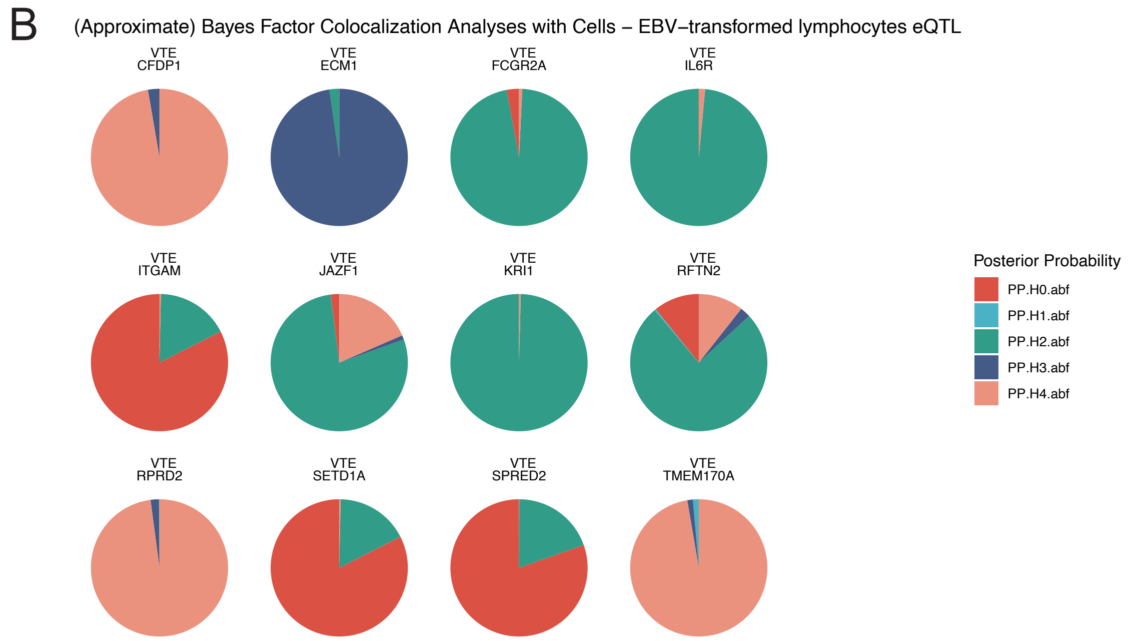


Supplementary Figure 11

A

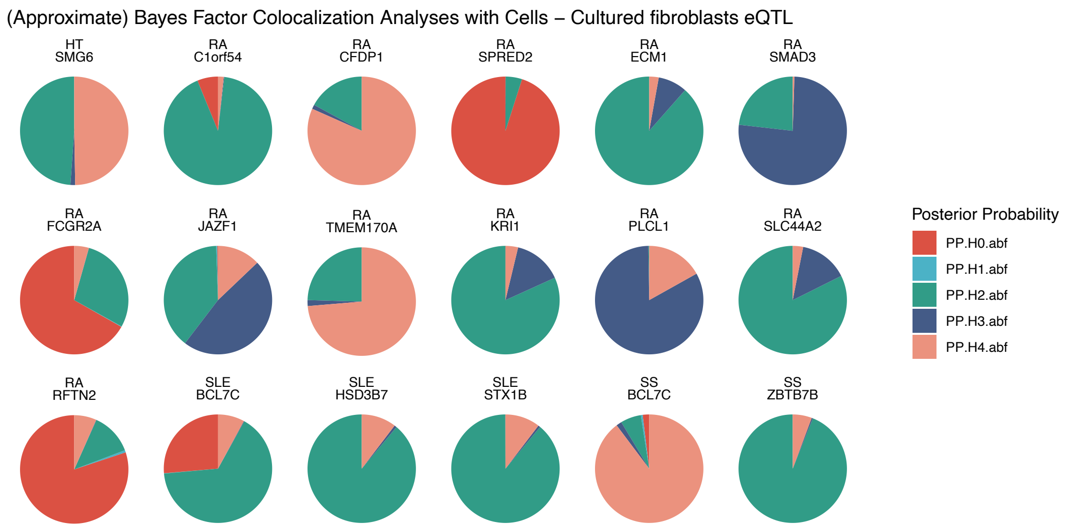


B

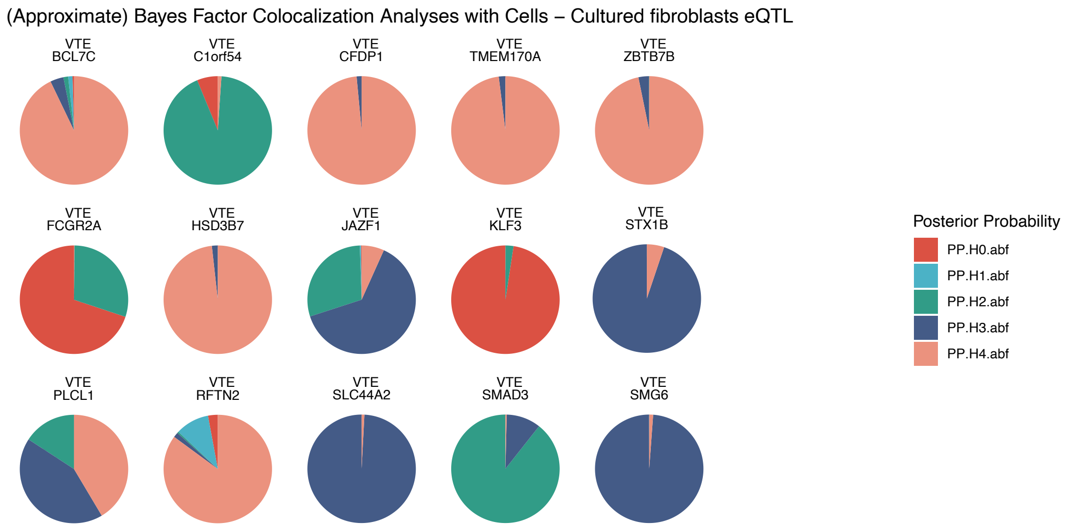


Supplementary Figure 12

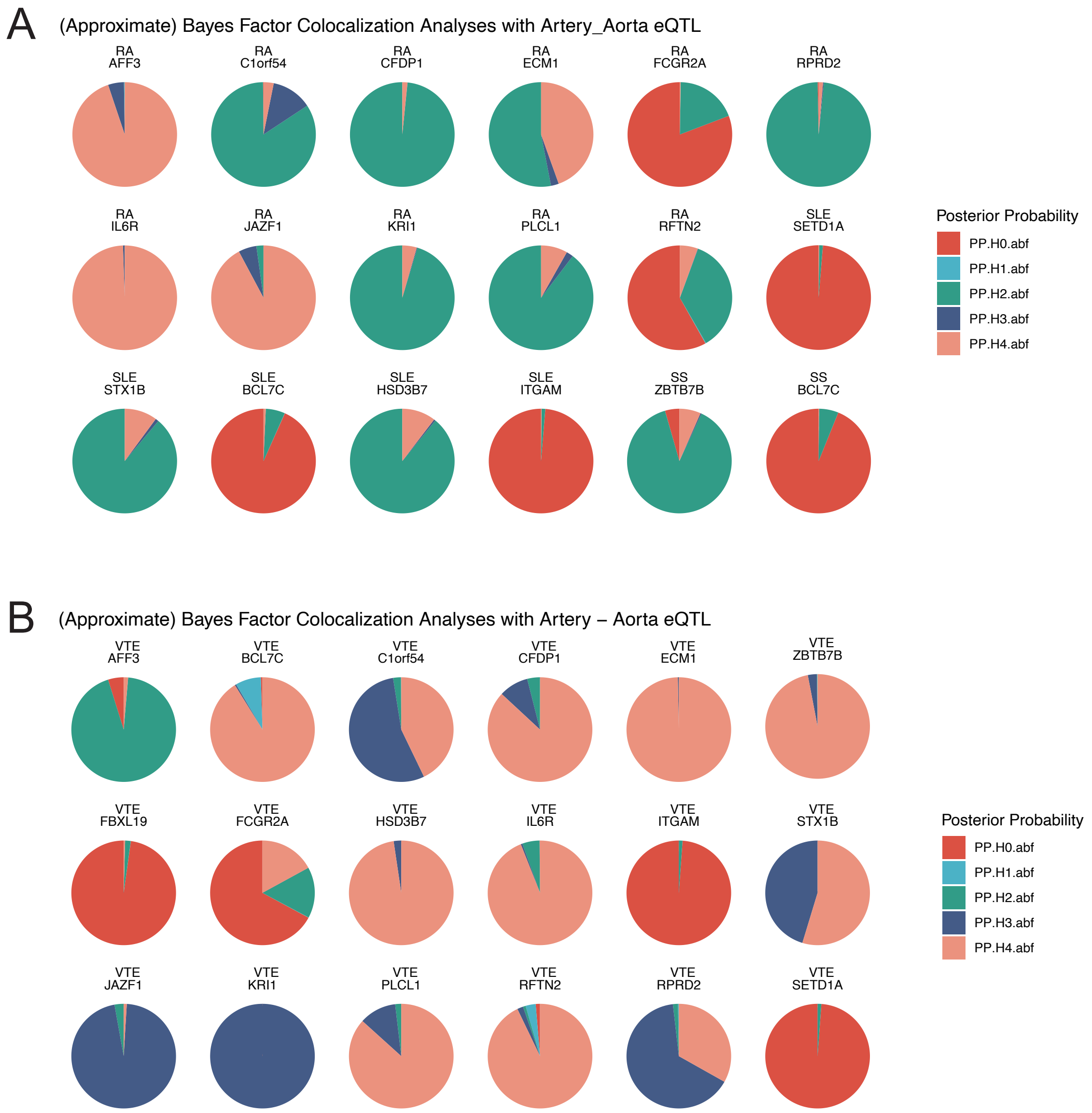


Supplementary Figure 13

A

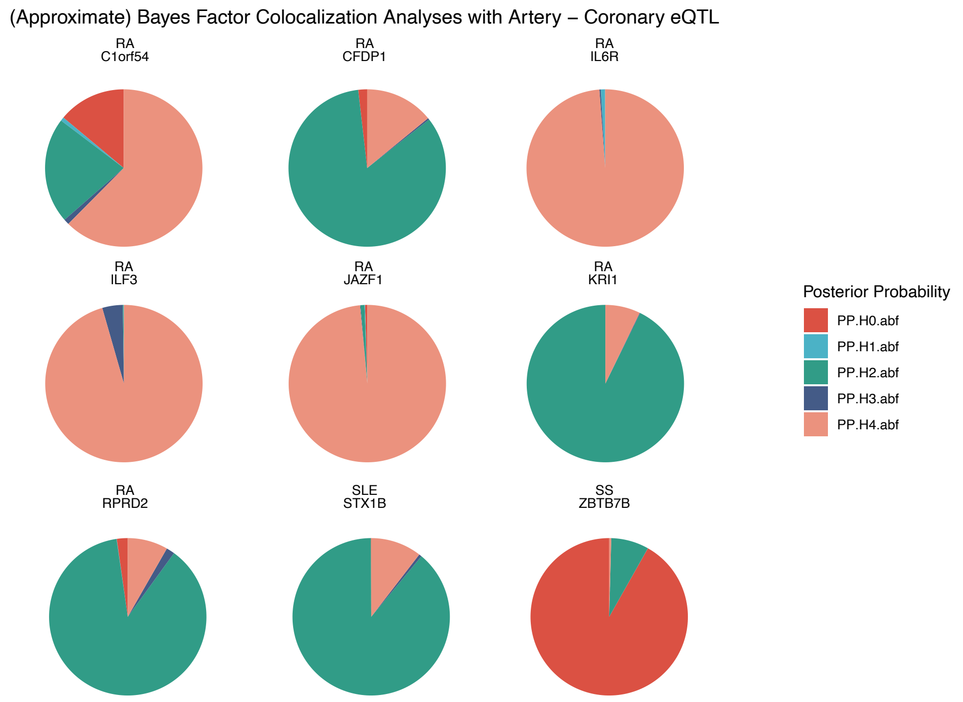


B

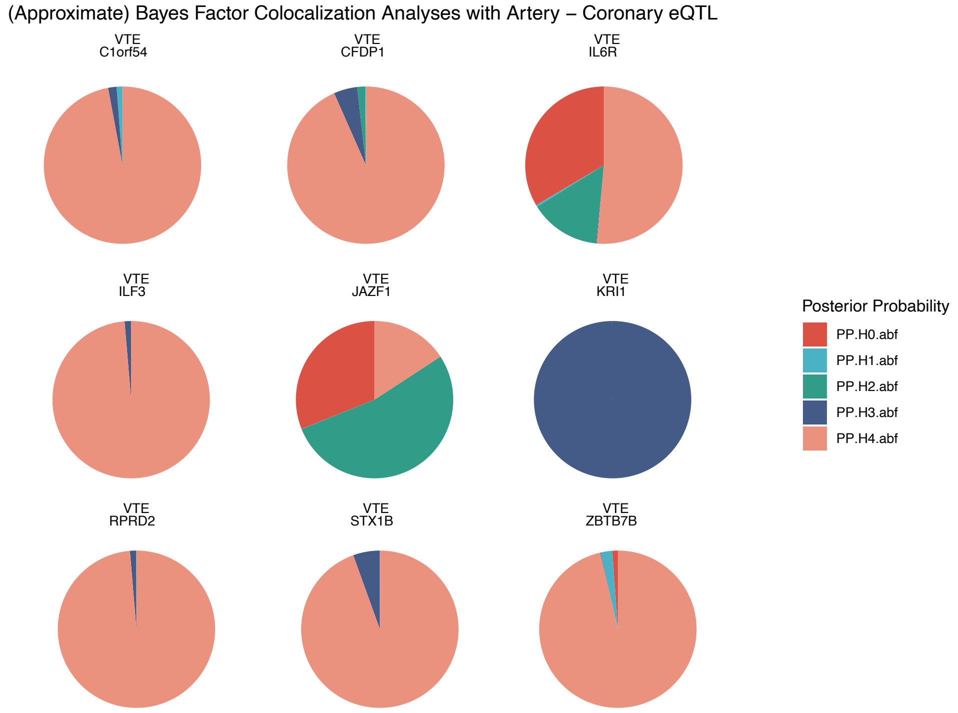


Supplementary Figure 14

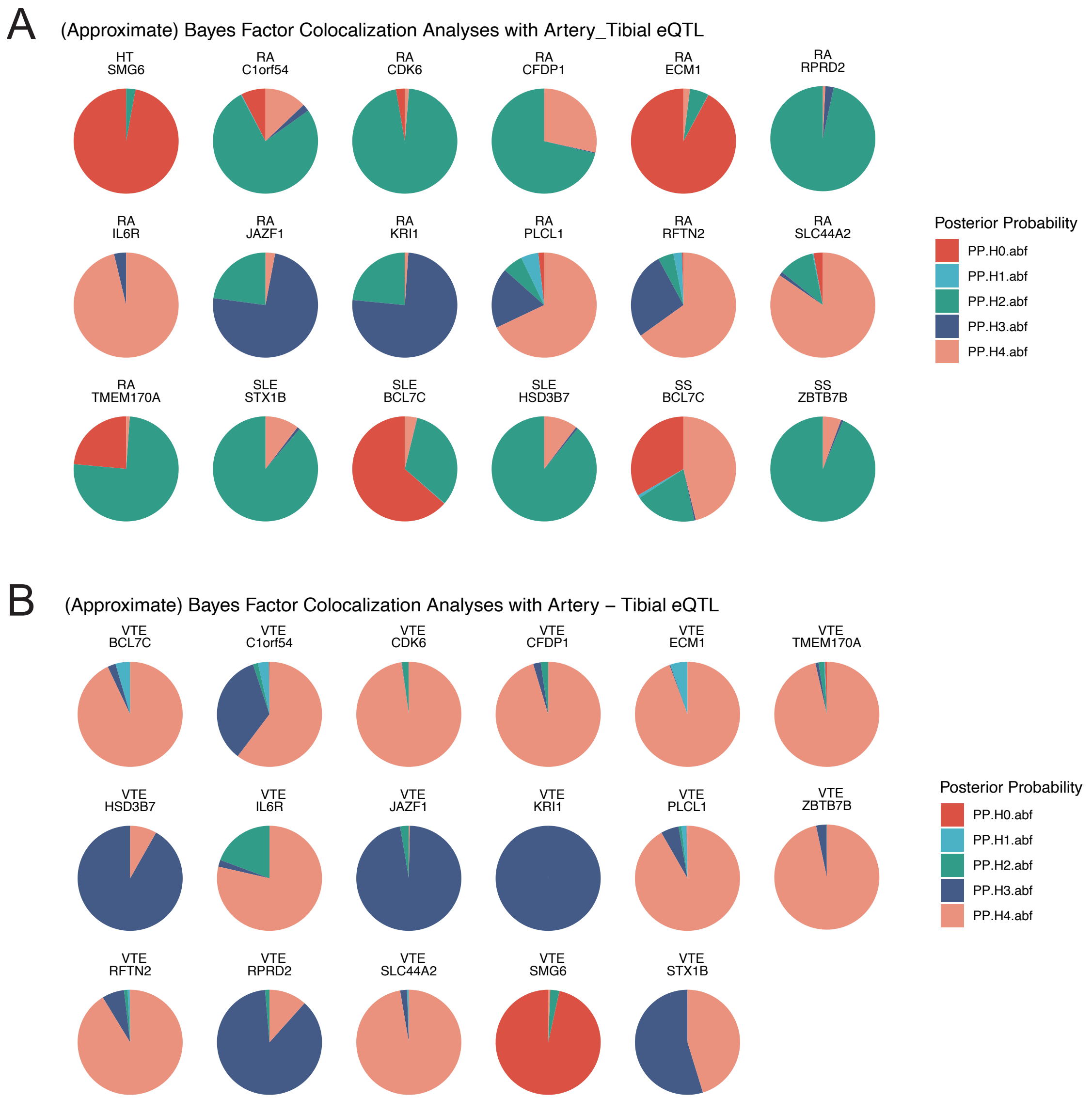


Supplementary Figure 15

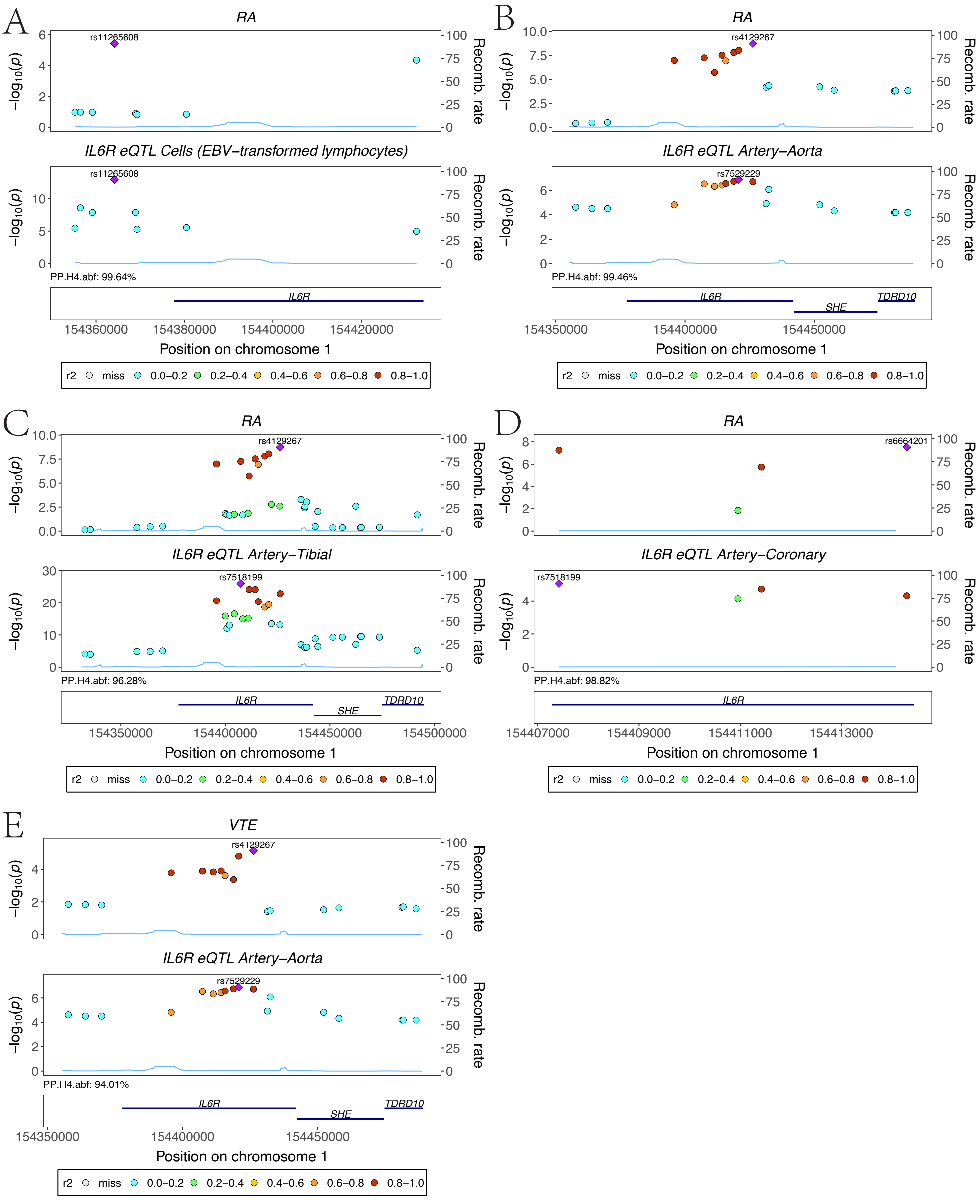


Supplementary Figure 16

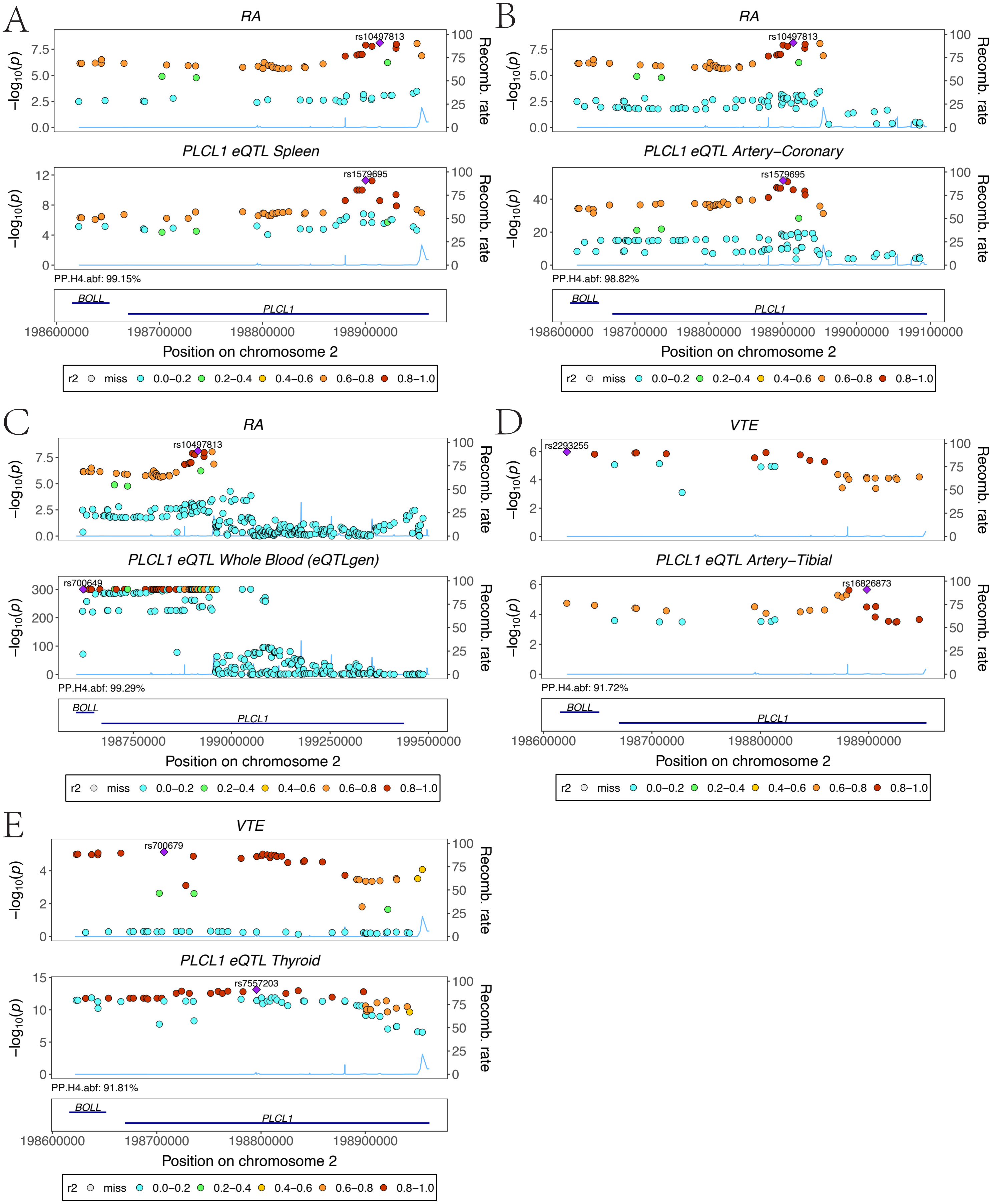


Supplementary Figure 17

A

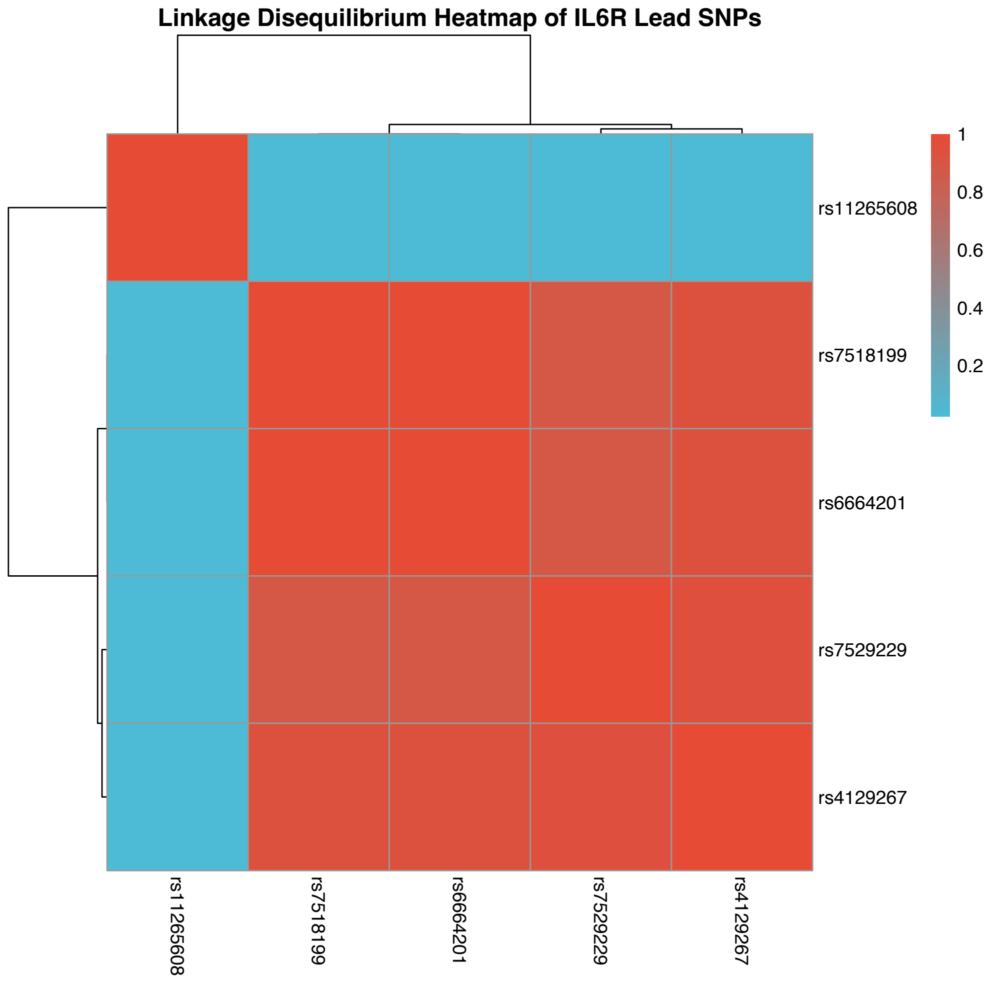


B

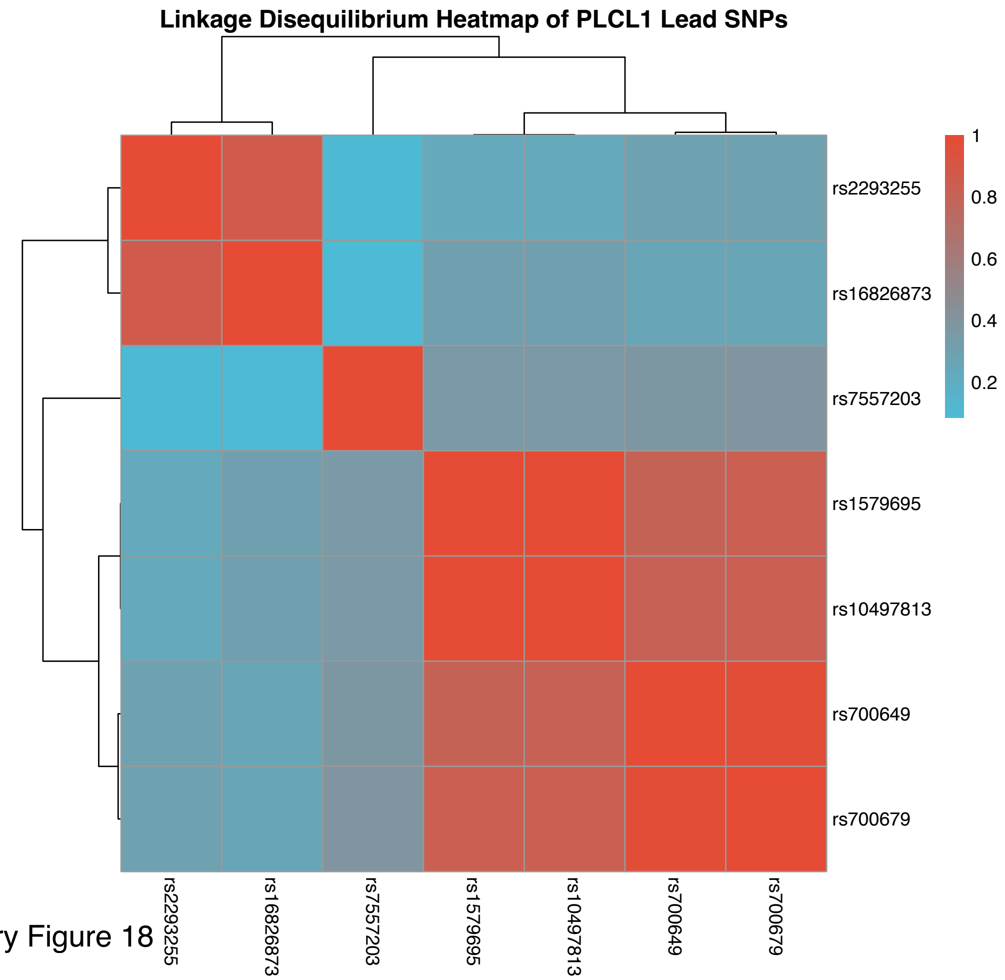


Supplementary Figure 18

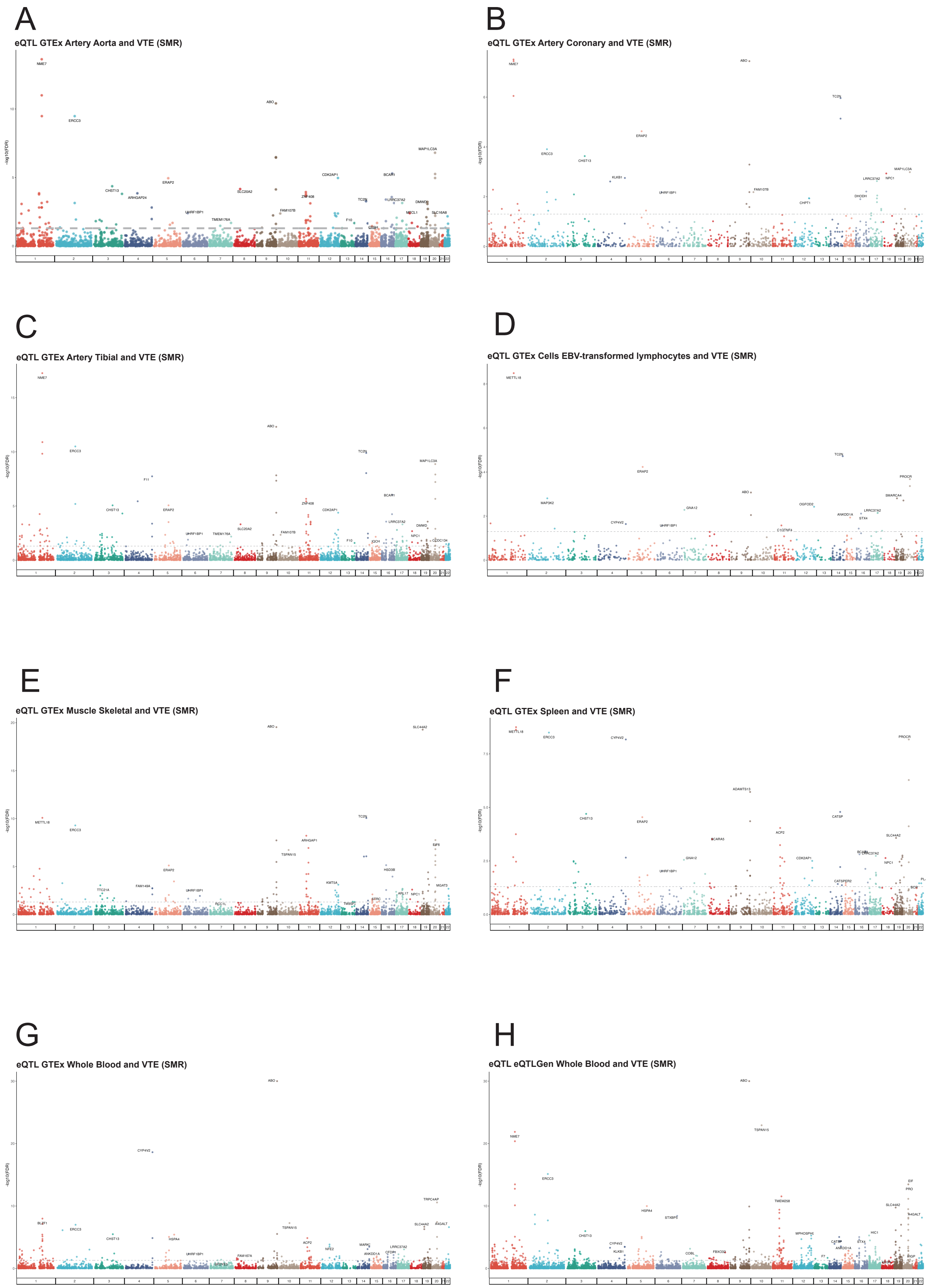


Supplementary Figure 19

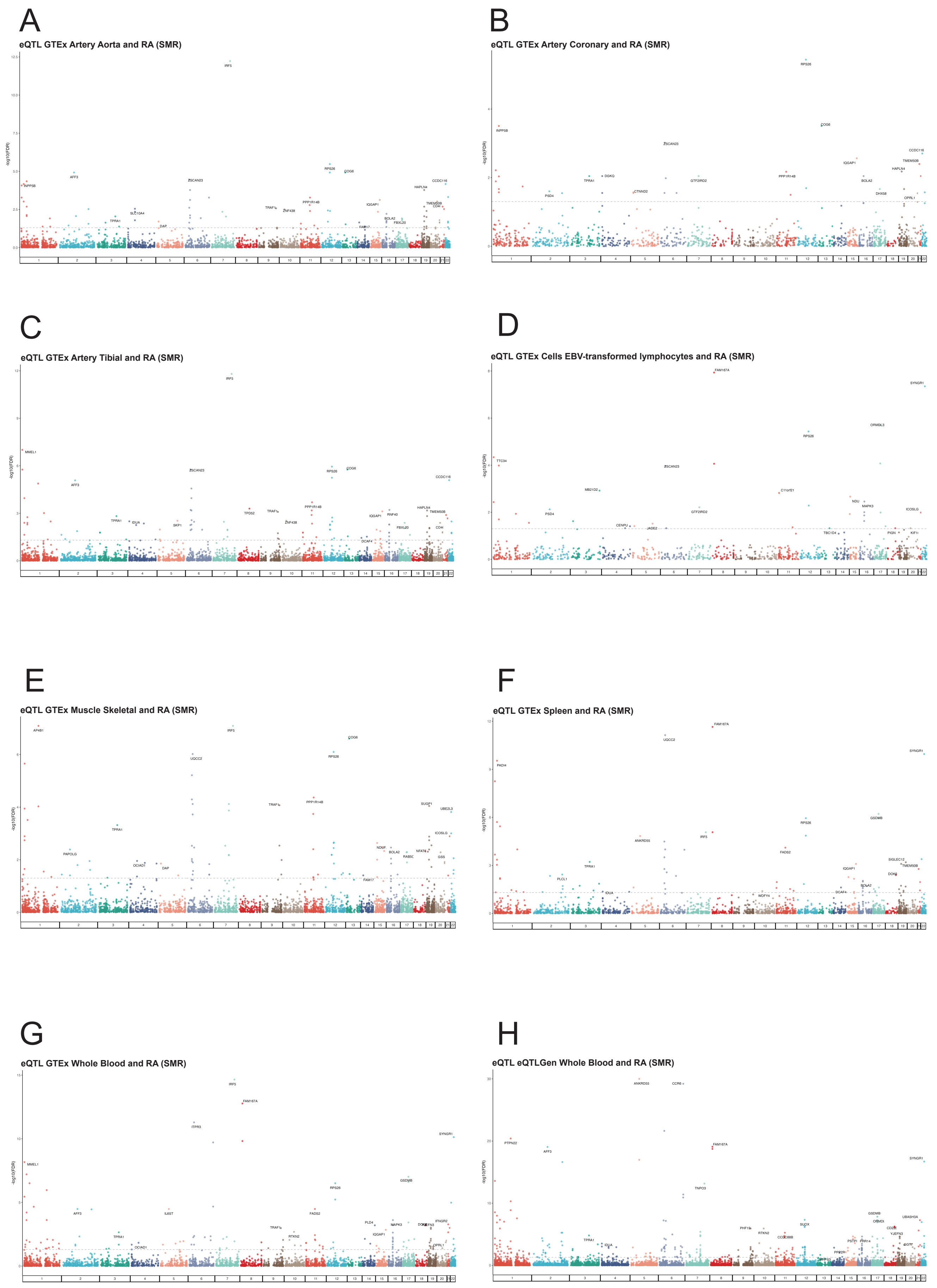


Supplementary Figure 20

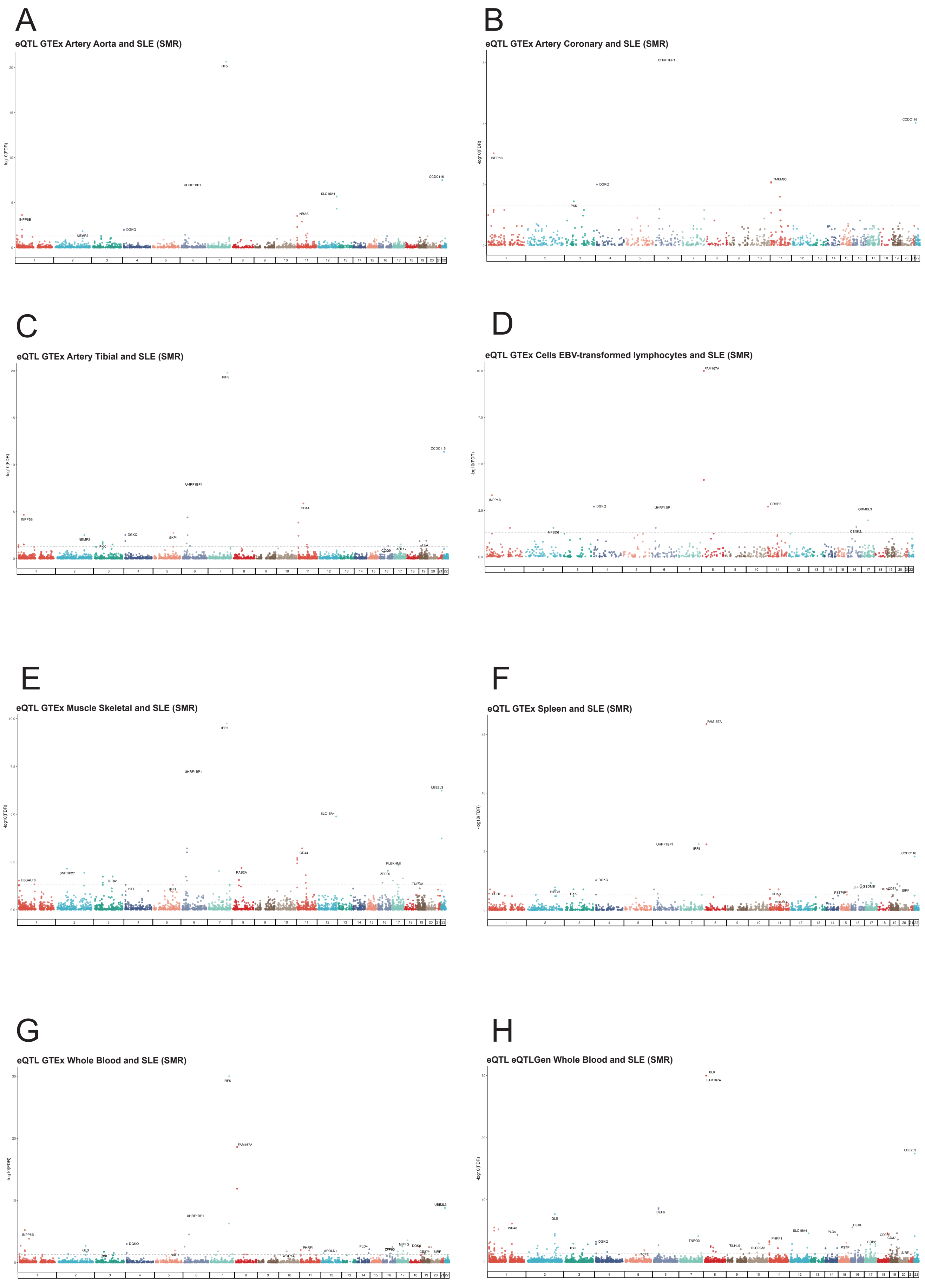


Supplementary Figure 21

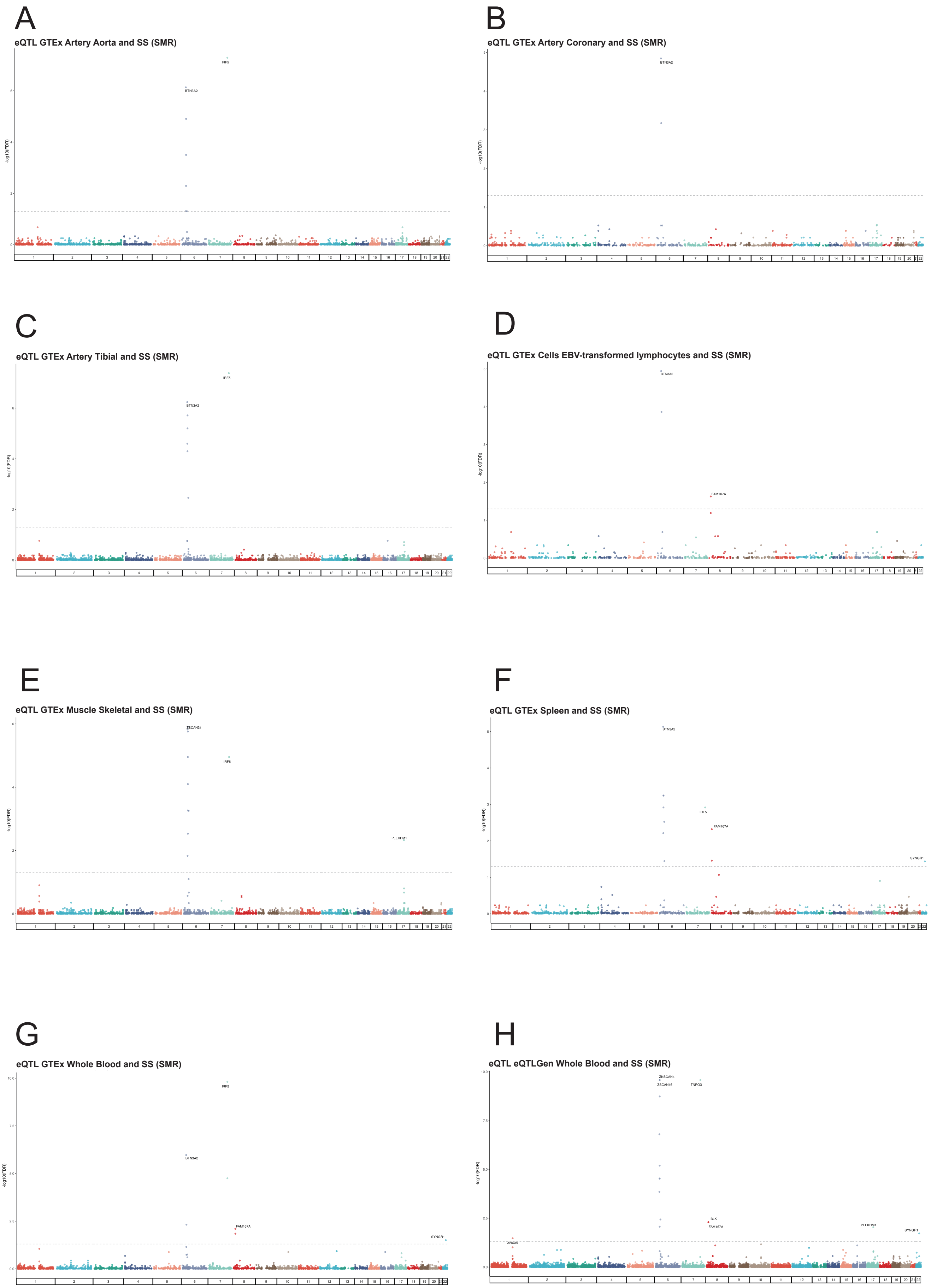


Supplementary Figure 22

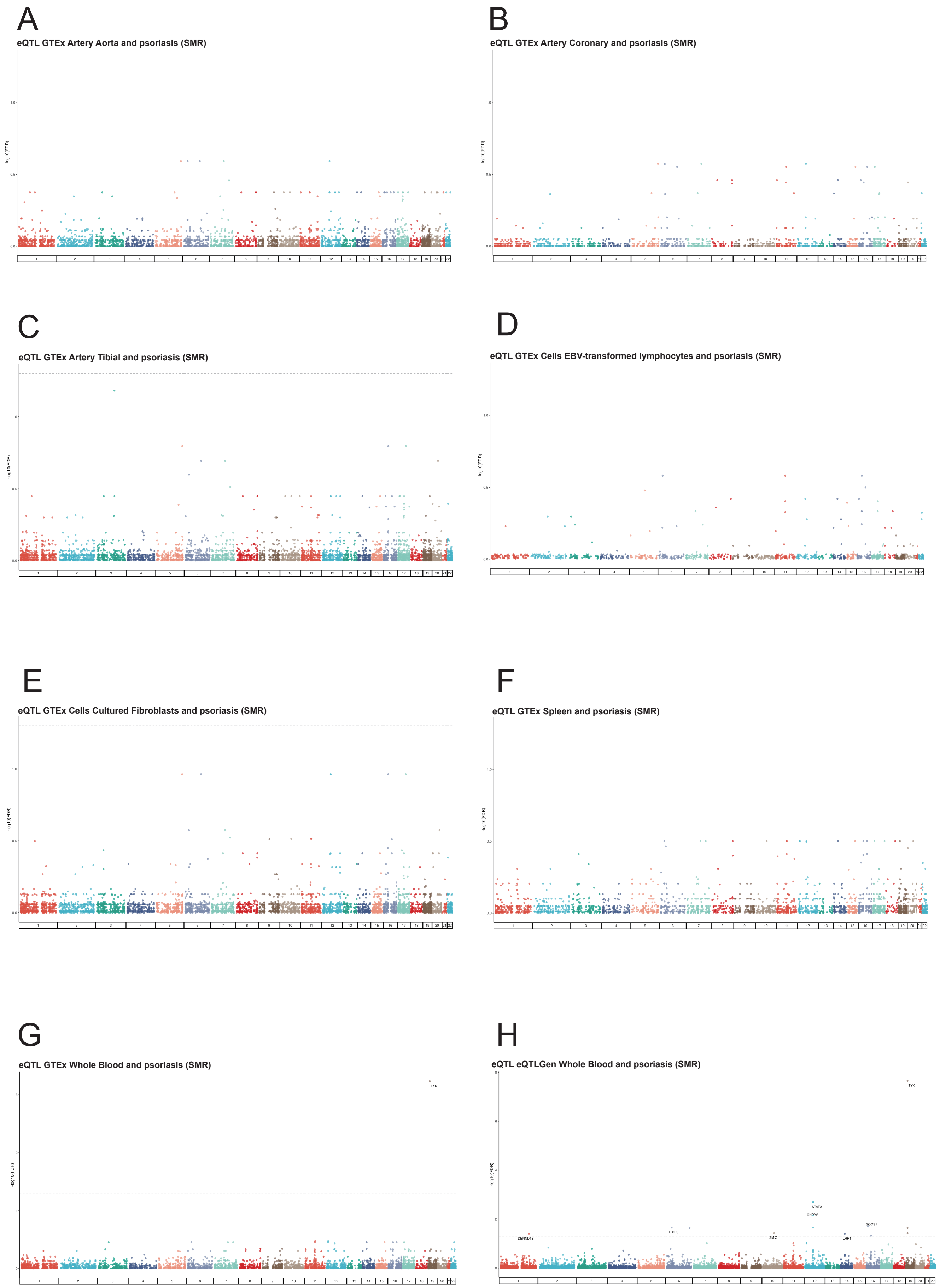


Supplementary Figure 23

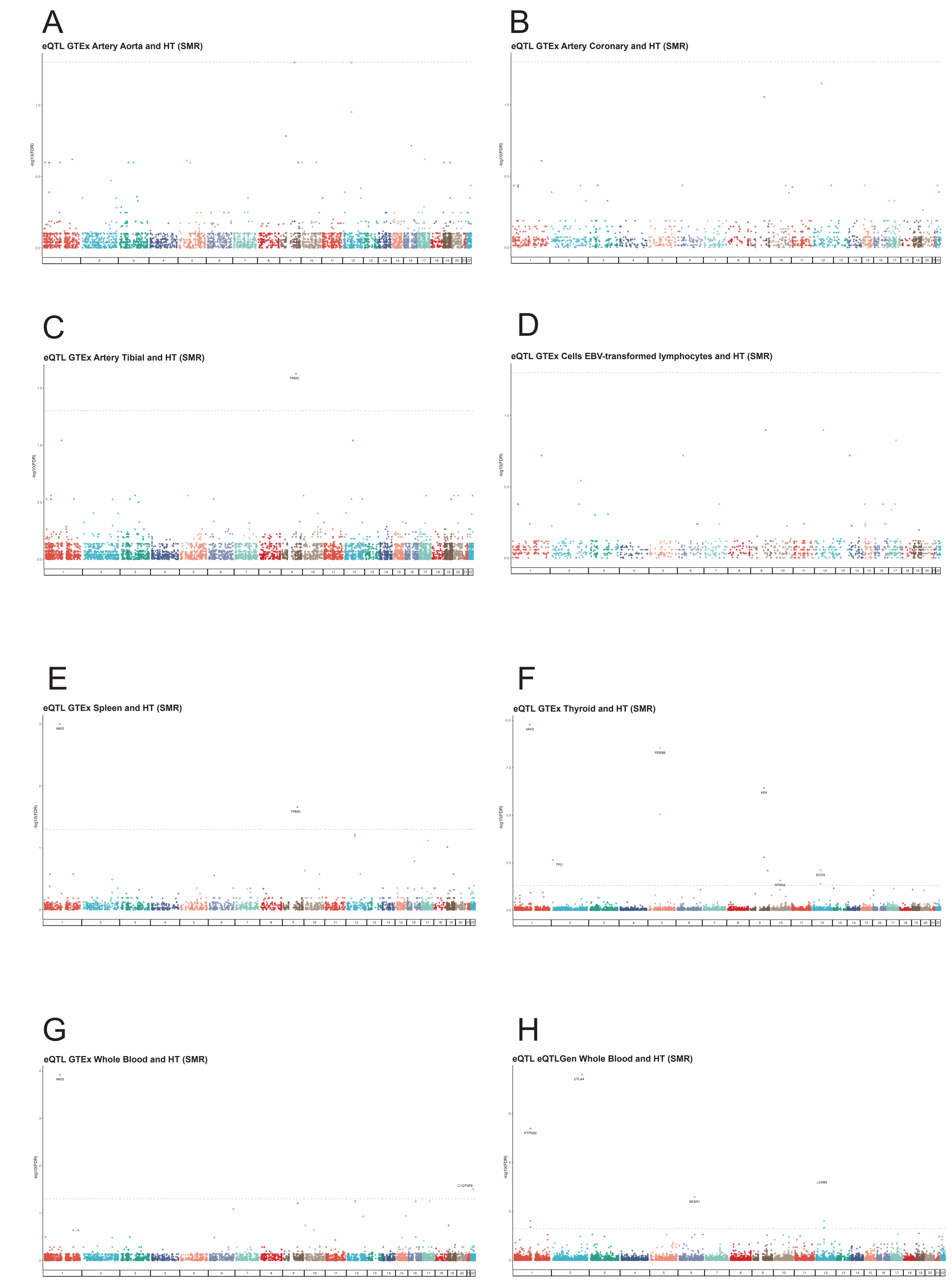


Supplementary Figure 24
